# A simplified risk model for pretreatment stratification of newly diagnosed acute myeloid leukemia patients treated with venetoclax and azacitidine

**DOI:** 10.1101/2024.12.02.24318344

**Authors:** Nazmul Islam, Jamie S. Reuben, Justin L. Dale, Jingjing Zhang, James W. Coates, Karan Sapiah, Frank R. Markson, Lezhou Wu, Ujjwal V. Kulkarni, Michael Boyiadzis, Clayton A. Smith

**Author notes:** Communicating Author: Nazmul Islam, PhD, RefinedScience, 2115 N Scranton Street, Suite 2-70 Aurora Colorado, 80045, USA.

## Abstract

Venetoclax plus azacitidine (ven/aza) is a new standard of care for adult Acute Myeloid Leukemia (AML) patients who are not candidates for intensive therapies. Risk stratification approaches have been proposed to identify patients with favorable, intermediate, and adverse therapeutic outcomes following ven/aza and other lower intensive therapies. However, most have been developed for retrospective data analyses and have limitations in their application to upfront risk stratification of newly diagnosed patients. Here, we describe an AML risk model, termed the Refined Risk Model (RRM), that is specific for ven/aza, addresses important real-world considerations and utilizes pathology features that have the potential to be available relatively quickly-and-broadly following diagnosis. The RRM was developed and internally validated using a single center cohort of 316 AML patients from the University of Colorado treated upfront with ven/aza, and then externally validated on an AML cohort from a nationwide electronic health record-derived de-identified AML database. The RRM effectively stratified patients into Adverse, Intermediate, and Favorable groups across both the internal and external cohorts; it performed well in subsets with or without allogeneic transplant recipients, demonstrated tolerance to missing data, and showed numerical performance comparable to or exceeding the existing alternatives such as the European Leukemia Network (ELN 2022) and molecular prognostic risk signature (mPRS) models. These findings suggest that the RRM may have potential application in defining the prognostic mortality risk for newly diagnosed AML patients, which may help guide clinical trial design and execution as well as other important elements of AML clinical decision support.

## Introduction

Acute myeloid leukemia (AML) is diagnosed in ∼15,000-20,000 persons each year in the United States ^1^. Treatment for young and fit patients typically involves aggressive initial intensive induction chemotherapy (IC) such as an anthracycline plus cytosine arabinoside followed by consolidative chemotherapy or allogeneic hematopoietic cell transplant (allo-HCT) ^2, 3^. For older or less fit patients, the bcl-2 directed agent venetoclax combined with a hypomethylating agent (HMA) such as azacitidine or decitabine has become a standard of care ^4–6^. For the IC type therapies, a variety of prognostic strategies have been developed to stratify patients into subgroups with varying outcomes ^7–15^. The European Leukemia Network (ELN) has developed the widely used ELN 2017 (ELN17) and more recently, the ELN 2022 (ELN22) risk categories ^16,17^. These divide patients into favorable, intermediate, and adverse-risk groups based on AML cytogenetic (CYT), fluorescence in situ hybridization (FISH), and next generation sequencing (NGS) features associated with overall survival (OS) outcomes ^16, 17^. However, it has been shown that the original ELN risk models do not effectively stratify patients treated with lower intensity regimens, including ven/aza, likely because the risk features were largely based on treatment outcomes following IC ^18–23^. Recently, several new risk stratification approaches have been proposed for AML patients treated with low intensity regimens that appear to have improved performance over the prior ELN17 and ELN22 models ^18–22^. These stratification approaches have been explored in the post-hoc analysis of clinical trial and real-world data (RWD) so their applicability and practicality in upfront patient risk assignment remains unclear. In addition, most of these methods place a heavy reliance on NGS results which may be variably available for upfront patient allocation due to expense, technical challenges, accessibility, and turn-around time ^24^. Lastly, data missingness is a common issue in both clinical trial and RWD. Most of the current AML risk stratification approaches make no provision, other than excluding patients, for dealing with missing data and this strategy may induce selection bias and confound up front risk stratification ^25, 26^.

Recently, we have described a machine learning (ML) based AML risk stratification strategy for newly diagnosed AML patients treated specifically with ven/aza that effectively stratified newly diagnosed patients and addressed a variety of commonly encountered RWD issues including data missingness, data skewing, biases based on underlying assumptions, and other considerations ^27^. In the current study, we describe the development and testing of a relatively simple risk stratification model derived from this ML based strategy, termed the Refined Risk Model (RRM), that was designed to be applicable to patient risk classification in the upfront setting.

## Methods

The RRM training dataset included 316 adult patients from the University of Colorado (CU) with newly diagnosed AML treated front line with ven/aza either as standard of care or in a clinical trial between January 2015 and March 2024. The RRM external testing set was an independent, heterogeneous dataset (termed as real-world cohort (RWC)) comprised of 971 AML patients treated with ven/aza at 87 unique sites of care obtained from the nationwide Flatiron Health electronic health record-derived, de-identified database which is a longitudinal database, comprising patient-level structured and unstructured de-identified data, curated via technology-enabled abstraction ^28, 29^. Table 1 summarizes patient features, while Figure 1 illustrates patient and cohort subset management. A pictorial representation of the AML phenotypic and genetic features of these two cohorts is illustrated in Figure 2. Kaplan-Meier (KM) analyses of OS by important features are performed (Supplemental Figure 1-5). Data definitions, standardization, and harmonization details are summarized in *Supplemental Section A.1*, *Supplemental Table 2-3*, and a previous study ^27^.

**Figure 1.**
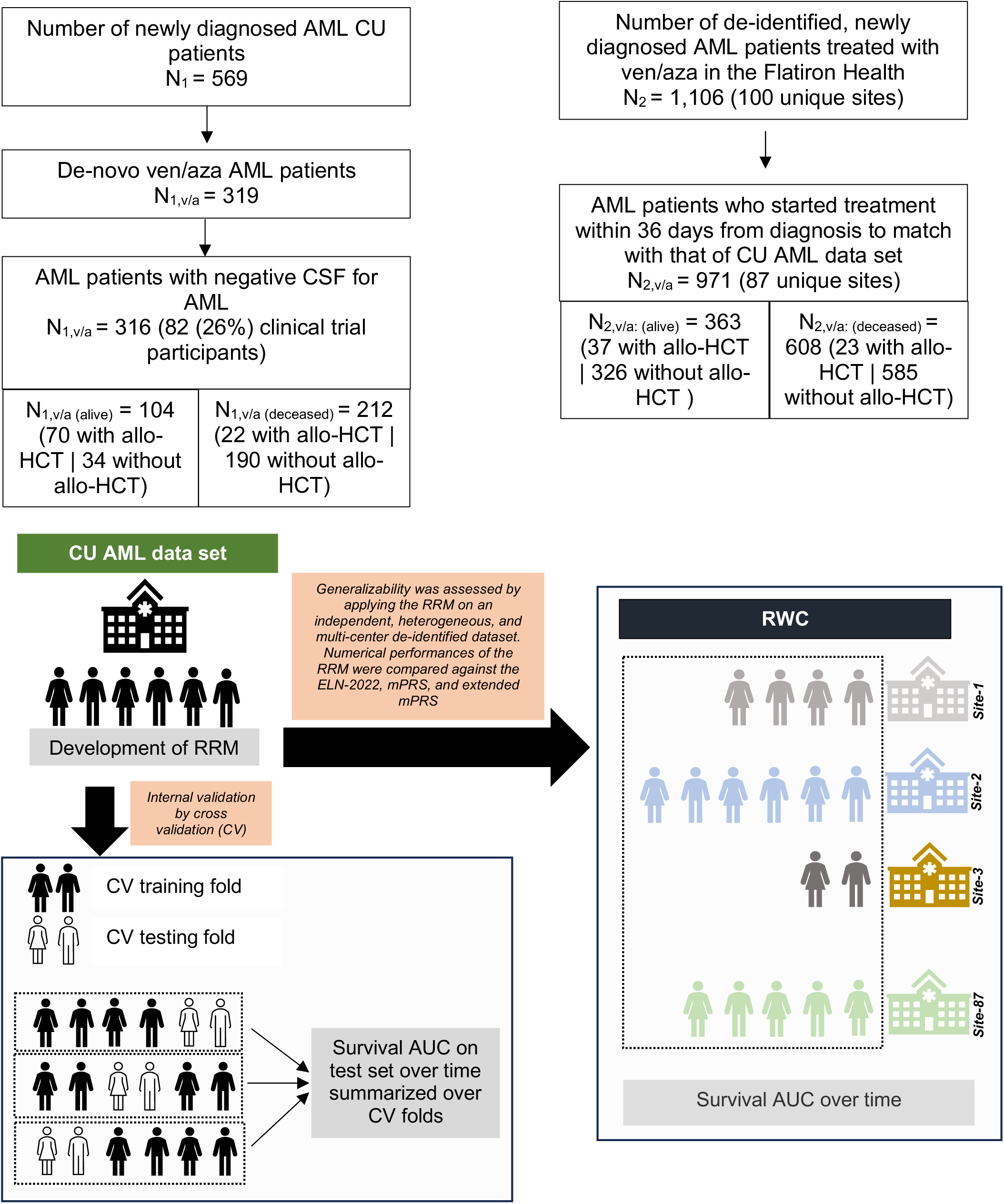
Cohort management. Summary of CU and RWC cohorts used in the refined risk model (RRM) development and testing.

**Figure 2.**
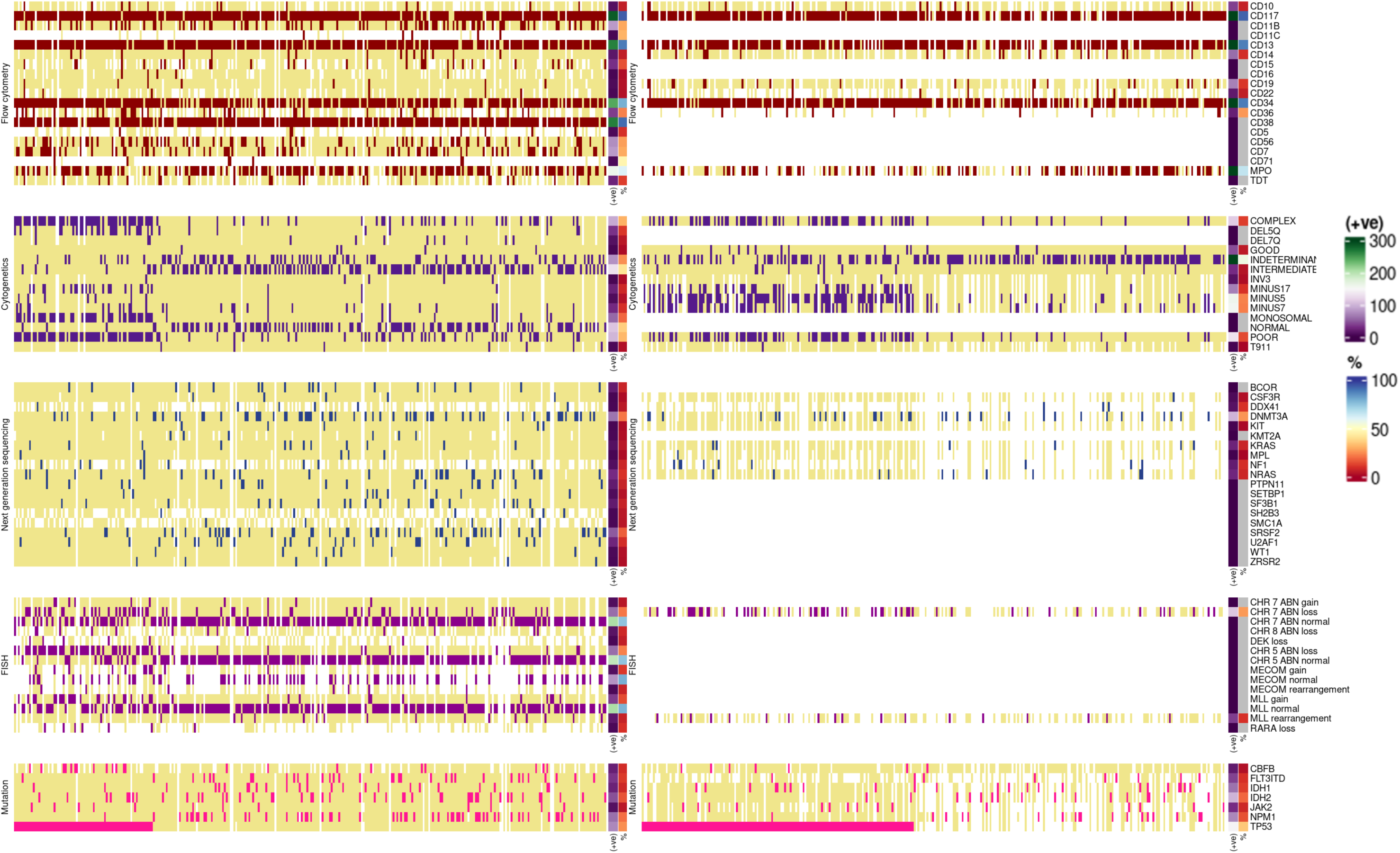
OncoPlot for CU and RWC datasets. Data represents the AML phenotypic and genotypic features on a per patient basis for the CU (left panel) and RWC (right panel) datasets. The top legend on the right (+ve) refers to the number of patients mutated or positive for each feature and the lower legend (%) refers to the proportion of patients with mutated or positive label for that feature in the observed (complete) data.

**Table 1.** Summary statistics for CU and RWC datasets. (%) is the percentage of available data and [%] is percentage of total patient counts. *Standardized mean differences (SMD) were calculated after excluding missing cases (i.e., it compares the respective proportion reported within the first parenthesis). SMD > 0.10 for each covariate refers to substantial systematic differences between CU and RWC patients in the sample.

|  |  | CU | RWC | *SMD |
| --- | --- | --- | --- | --- |
| **N** |  | 316 | 971 |  |
| **†Demographics** |  |  |  |  |
| **Age, median (IQR)** |  | 71.0 (11.0) | 76.0 (9.0) | 0.63 |
| **Male, n (%)** |  | 171 (54.1) | 563 (58.0) | 0.07 |
| **ECOG, n (%) [%]†** |  |  |  | 0.15 |
|  | 0 | 48 (23.8) [15.2] | 92 (30.3) [ 9.5] |  |
|  | 1 | 112 (55.4) [35.4] | 152 (50.0) [15.7] |  |
|  | 2 | 30 (14.9) [ 9.5] | 42 (13.8) [ 4.3] |  |
|  | ≥3 | 12 (5.9) [ 3.5] | 18 (5.9) [ 1.9] |  |
| **†ELN 2022, n (%) [%]†** |  |  |  | 0.56 |
|  | Favorable | 54 (17.3) [17.1] | 23 (3.3) [ 2.4] |  |
|  | Intermediate | 50 (16.0) [15.8] | 223 (31.7) [23.0] |  |
|  | Adverse | 208 (66.7) [65.8] | 458 (65.1) [47.2] |  |
| **Comorbidities, n (%)** |  |  |  |  |
| **Obesity** |  | 124 (39.2) | 50 (5.1) | 0.90 |
| **Prior non-AML cancer** |  | 165 (52.2) | 251 (25.8) | 0.56 |
| **Prior heart disease** |  | 140 (44.3) | 174 (17.9) | 0.60 |
| **Prior MDS** |  | 16 ( 5.1) | 182 (18.7) | 0.43 |
| **Prior CKD** |  | 95 (30.1) | 117 (12.0) | 0.45 |
| **Prior coagulopathy** |  | 35 (11.1) | 22 ( 2.3) | 0.36 |
| **Prior VTE** |  | 40 (12.7) | 29 ( 3.0) | 0.37 |
| **Prior COPD** |  | 34 (10.8) | 49 ( 5.0) | 0.21 |
| **Prior GERD** |  | 111 (35.1) | 130 (13.4) | 0.52 |
| **Prior hyperlipidemia** |  | 127 (40.2) | 191 (19.7) | 0.46 |
| **Prior hypertension** |  | 168 (53.2) | 301 (31.0) | 0.46 |
| **Prior hypothyroidism** |  | 79 (25.0) | 88 ( 9.1) | 0.43 |
| **†Genetics, n (%) [%]†** |  |  |  |  |
| **AML composite mutations** |  |  |  |  |
| **TP53** | (-ve) | 222 (75.0) [70.3] | 272 (64.9) [28.0] | 0.23 |
|  | (+ve) | 74 (25.0) [23.4] | 147 (35.1) [15.1] |  |
| **IDH1** | (-ve) | 275 (92.0) [87.0] | 461 (88.1) [47.5] | 0.13 |
|  | (+ve) | 24 (8.0) [ 7.6] | 62 (11.9) [ 6.4] |  |
| **IDH2** | (-ve) | 254 (84.7) [80.4] | 428 (81.2) [44.1] | 0.09 |
|  | (+ve) | 46 (15.3) [14.6] | 99 (18.8) [10.2] |  |
| **NPM1** | (-ve) | 236 (79.7) [74.7] | 416 (85.1) [42.8] | 0.14 |
|  | (+ve) | 60 (20.3) [19.0] | 73 (14.9) [ 7.5] |  |
| **FLT3-ITD** | (-ve) | 267 (88.7) [84.5] | 367 (90.2) [37.8] | 0.05 |
|  | (+ve) | 34 (11.3) [10.8] | 40 (9.8) [ 4.1] |  |
| **Cytogenetics** |  |  |  |  |
| **Good risk cytogenetics** | (-ve) | 311 (98.4) [98.4] | 926 (95.4) [95.4] | 0.18 |
|  | (+ve) | 5 (1.6) [1.6] | 45 (4.6) [ 4.6] |  |
| **Inv3** | (-ve) | 297 (98.3) [94.0] | 675 (98.1) [69.5] | 0.02 |
|  | (+ve) | 5 (1.7) [ 1.6] | 13 (1.9) [ 1.3] |  |
| **Minus5** | (-ve) | 293 (97.0) [92.7] | 530 (76.9) [54.6] | 0.63 |
|  | (+ve) | 9 (3.0) [ 2.8] | 159 (23.1) [16.4] |  |
| **Del7q** | (-ve) | 290 (96.0) [91.8] | NA | NA |
|  | (+ve) | 12 (4.0) [3.8] | NA |  |
| **Minus7** | (-ve) | 273 (90.4) [86.4] | 533 (77.5) [54.9] | 0.36 |
|  | (+ve) | 29 (9.6) [9.2] | 155 (22.5) [16.0] |  |
| **Minus17** | (-ve) | 278 (92.1) [88.0] | 619 (90.0) [63.7] | 0.07 |
|  | (+ve) | 24 (7.9) [ 7.6] | 69 (10.0) [ 7.1] |  |
| **Complex cytogenetics** | (-ve) | 209 (69.2) [66.1] | 856 (88.2) [88.2] | 0.48 |
|  | (+ve) | 93 (30.8) [29.4] | 115 (11.8) [11.8] |  |
| **\*t(9;11)** | (-ve) | 295 (97.7) [93.4] | 684 (~98.0) [~70.0] | 0.15 |
|  | (+ve) | 7 (2.3) [ 2.2] | <5 (<1.0) [<1.0] |  |
| **Next generation sequencing** |  |  |  |  |
| **KIT** | (-ve) | 289 (98.3) [91.5] | 382 (98.5) [39.3] | 0.01 |
|  | (+ve) | 5 (1.7) [1.6] | 6 (1.5) [ 0.6] |  |
| **JAK2** | (-ve) | 284 (95.9) [89.9] | 353 (92.2) [36.4] | 0.16 |
|  | (+ve) | 12 (4.1) [ 3.8] | 30 (7.8) [ 3.1] |  |
| **CSF3R** | (-ve) | 286 (98.3) [90.5] | 333 (97.7) [34.3] | 0.05 |
|  | (+ve) | 5 (1.7) [ 1.6] | 8 (2.3) [ 0.8] |  |
| **KRAS** | (-ve) | 282 (95.9) [89.2] | 335 (93.1) [34.5] | 0.13 |
|  | (+ve) | 12 (4.1) [ 3.8] | 25 (6.9) [ 2.6] |  |
| **\*MPL** | (-ve) | 283 (98.3) [89.6] | 293 (~98.0) [~30.0] | 0.10 |
|  | (+ve) | 5 (1.7) [ 1.6] | <5 (<1.0) [< 1.0] |  |
| **DNMT3A** | (-ve) | 230 (78.5) [72.8] | 284 (74.0) [29.2] | 0.11 |
|  | (+ve) | 63 (21.5) [19.9] | 100 (26.0) [10.3] |  |
| **DDX41** | (-ve) | 140 (95.9) [44.3] | 118 (90.1) [12.2] | 0.23 |
|  | (+ve) | 6 (4.1) [ 1.9] | 13 (9.9) [ 1.3] |  |
| **NRAS** | (-ve) | 265 (89.8) [83.9] | 325 (92.3) [33.5] | 0.09 |
|  | (+ve) | 30 (10.2) [9.5] | 27 (7.7) [ 2.8] |  |

- The counts and percentages for T9;11 and MPL (highlighted in \*) are provided in intervals and approximate percentages since the counts in the frequency table are <5 for the RWC. - Summary statistics of features with superscript <sup>¶</sup> for the CU patients' cohort are discussed previously in a separate study <sup>27</sup> with the same group of authors.

The RRM was developed based on a previously described ML specific hazard model for OS where the risk of mortality was estimated over time by counterfactual arguments ^27^. The RRM utilizes AML diagnostic genetic features that could be potentially identified currently with CYT, polymerase chain reaction (PCR), FISH, and Sanger sequencing or other tests that have rapid turn-around, are relatively inexpensive, and widely available. Though not selected empirically, *FLT3-ITD* status was added in the feature list due to its clinical relevance and consistent importance in other risk models ^20, 21^. Risk stratification classifying each patient into Adverse, Intermediate, or Favorable groups was performed as described ^27^. Numerical performance of the RRM was compared with the ELN22, a newly described ven/HMA specific **m**olecular **p**rognostic **r**isk **s**ignature (mPRS) model, and a variant of the mPRS termed the extended-mPRS (e-mPRS) ^20, 22^ using both the CU and RWC cohorts. To address data missingness, comparative analyses were performed based on the different analytical datasets summarized in *Supplemental Table 1*. Comparative analyses in both the CU and RWC were evaluated with respect to 5 key features: (a) *equitability* (extent to which risk groups distribute patients equitably) was assessed by summary statistics, (b) *separability* (extent to which OS is associated with risk strata) was performed by assessing the survival differences between-and-within strata using KM analyses and the corresponding *P*-values testing the equality of curves, (c) *conformity* (extent to which risk groups overlap between methods) was assessed by Fleiss Kappa, (d) *predictability* (extent to which risk stratification predicts OS) was compared by survival metrics characterizing area **u**nder the **c**urve of **c**umulative case dynamic control receiver operative curves (coined as cAUC), and (e) *generalizability* (extent to which risk models reproduce results in an external dataset) was assessed by applying the RMs in the test RWC set and re-evaluating (a)-(d) independently. Further details of methodologies are provided in the *Supplemental Section A.1-A.2*.

All statistical tests were two-sided, with a significance level of 5% without multiplicity adjustments. All analyses were conducted using R, version 4.2.3.

## Results

### RRM model development

The RRM was developed as described in Methods and Supplemental Methods. A summary of the risk features and subject level risk assignment to Adverse, Intermediate, and Favorable categories based on these features is depicted in Figure 3A. Favorable risk features in the RRM included *IDH1*, *IDH2*, *NPM1*, and good risk cytogenetics. Adverse risk features included *TP53*, *Inv3*, *Minus 17*, *Del7q*, *t(9;11)*, *Minus 5*, and Complex cytogenetics. Intermediate risk features included either any good risk feature plus *FLT3-ITD*, the absence of any Favorable or Adverse risk features, or any 2-factor combination of Adverse and Favorable risk features in the same patient. First, the RRM was tested on the CU cohort (Figures 3B-D). As described in Methods and Supplemental Methods, the RRM was designed to account for the impact of allo-HCT recipients on OS and so all testing was performed using subsets of the CU cohort that included and excluded allo-HCT recipients. The RRM was also designed to manage missing data, so the numerical performance of the RRM was initially tested on the CU Full Analytic Set (FAS, i.e., the dataset that included all patients in the cohort regardless of data missingness) for both the allo-HCT included and allo-HCT excluded subsets. Of note in these analyses, data was complete for 281/316 (89%) patients in the subset including allo-HCT recipients and 200/224 (89%) patients in the subset excluding CU allo-HCT recipients. Initial analyses were performed for *equitability* of patient distribution and *separability* as defined in Methods. The RRM generated relatively equal distribution of patients between the three risk categories for both cohorts other than an elevated proportion in the Intermediate category (Figure 3B). This was consistent with the strategy of directing patients with unclear and undefined risk features as well as patients with the 2-feature combinations into the Intermediate group. The RRM effectively separated OS for the CU FAS allo-HCT included and excluded subsets by multiple statistical measures (Figure 3C, left panel, LR:*P*-value <0.0001 and Figure 3D left panel, LR:*P*-value <0.0001). The RRM also separated best response (BR) for these subsets as well (Figure 3C, right panel and Figure 3D, right panel; *P*-value <0.0001). Subsequently numerical performance of the RRM were tested for the impact of data missingness. To evaluate this, the RRM was applied to a subset of the CU cohort, termed the Complete Case Analytic Set (CCAS, i.e. the subset of patients that had all available data necessary for assignment to a particular risk group) and its performance was compared with that of the RRM using the CU FAS. No obvious differences with respect to *equitability* of patient assignments or *separability* were noted between the CCAS and FAS datasets (*Supplemental Figure 6A*; ∼23%:43%:33% for three risk groups respectively; LR *P*-value 0.0001). As a further test of the impact of incomplete data on the behavior of the RRM, missing data was imputed using multivariate models from the observed data to generate an imputed CU dataset, termed the Imputed Analytic Set (IAS).

**Figure 3.**
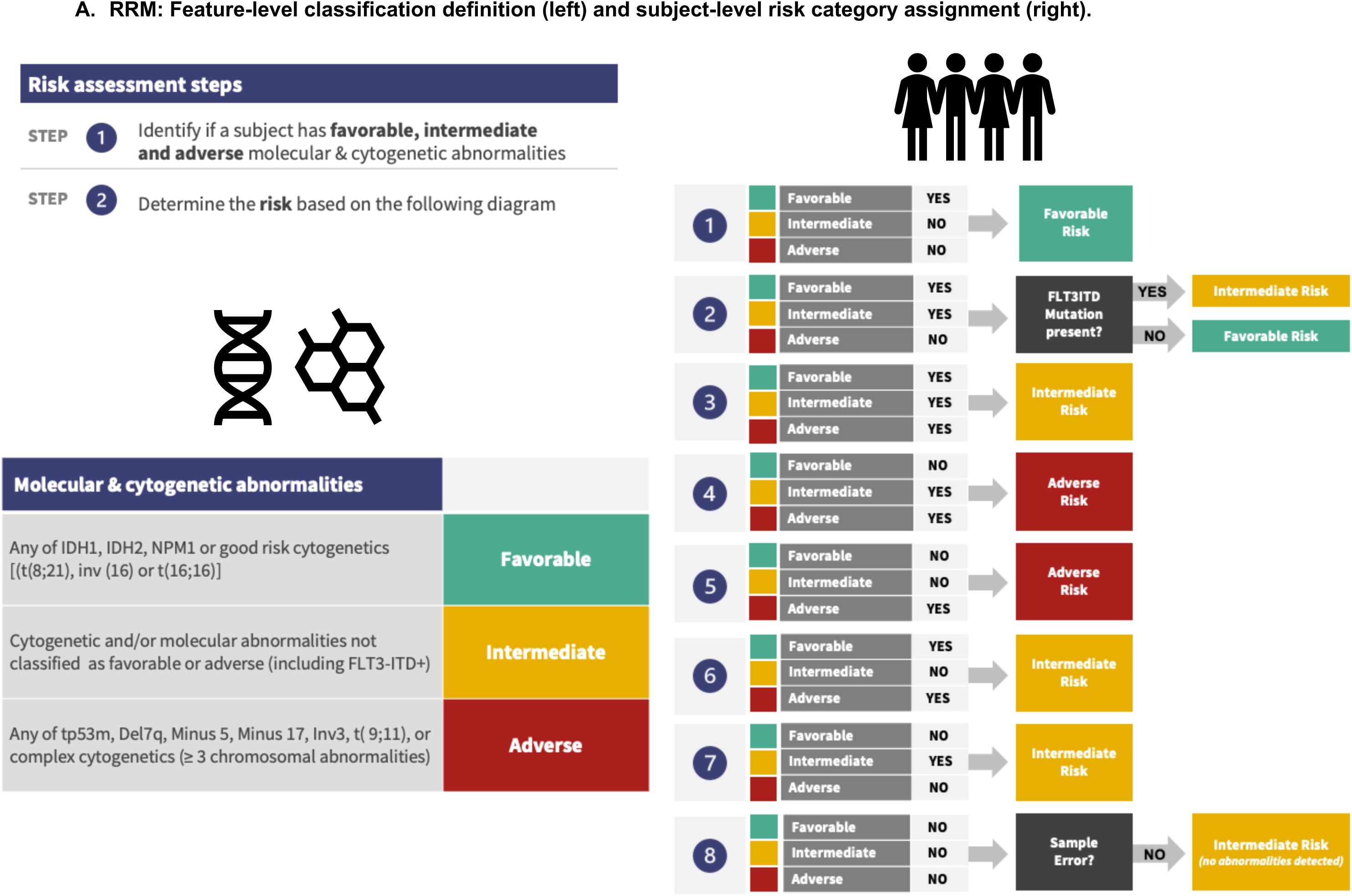
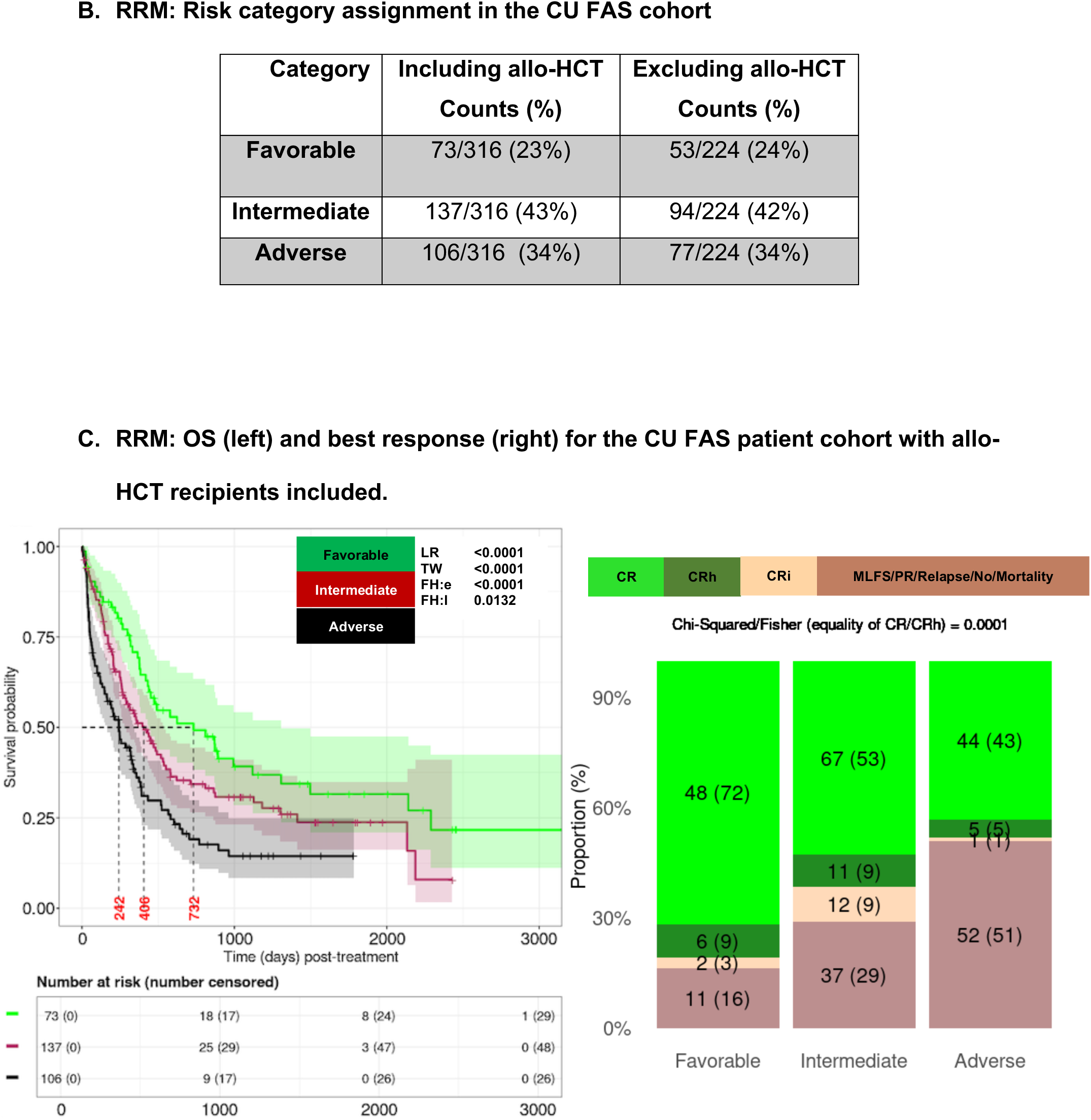
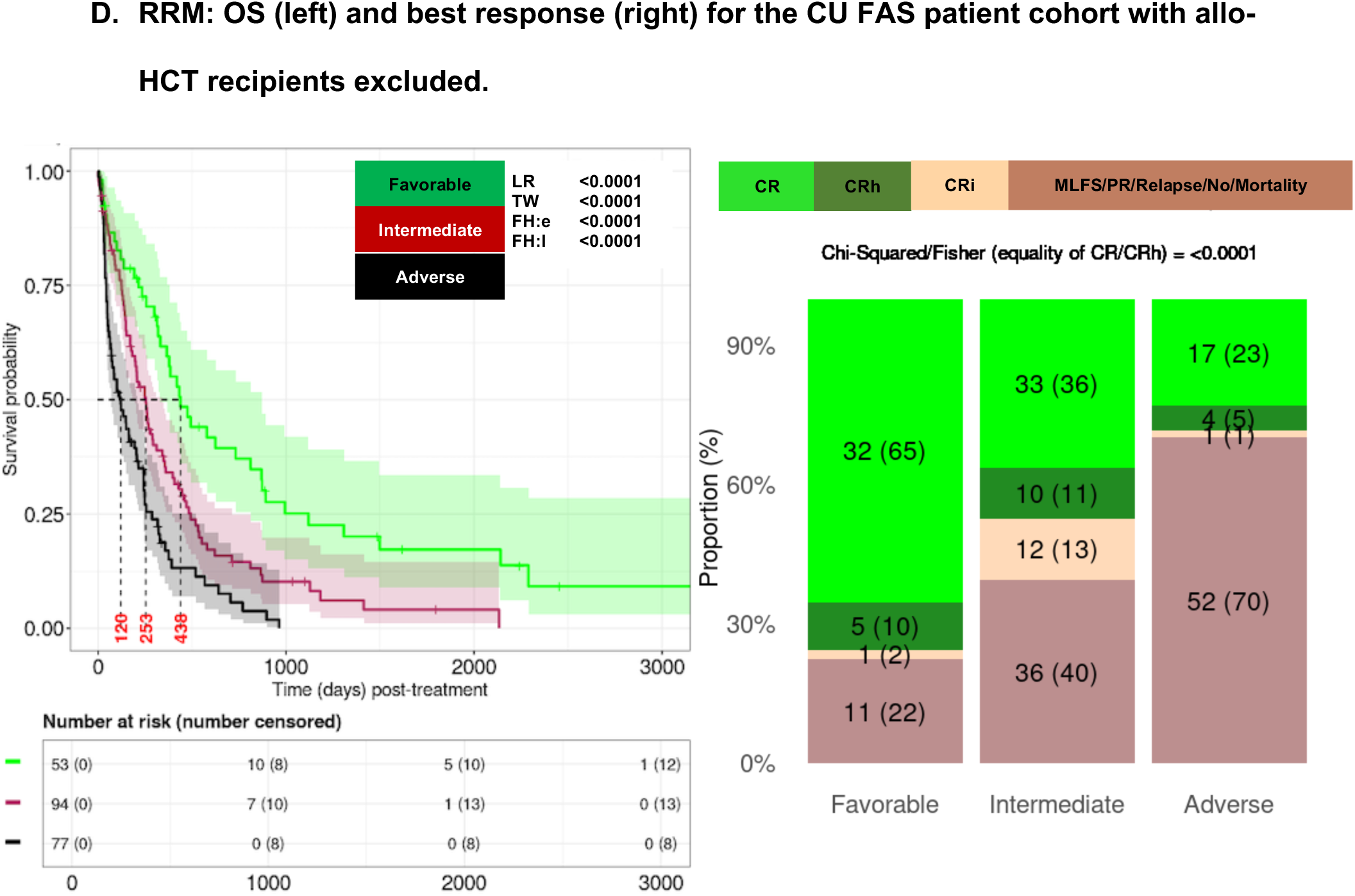
Refined Risk Model (RRM). A) Definition of the RRM features associated with AML risk groups for OS (left panels) and methods for assigning patients to Adverse, Intermediate and Favorable risk categories (right panels); B) Frequency of patients in the CU Full Analytic Set (FAS, i.e. total patients) assigned to the different risk categories; C) Application of the RRM to the ven/aza treated from the CU FAS subset with allo-HCT recipients included for OS (left panel) and best response (BR, right panel); D) Application of the RRM to the ven/aza treated CU FAS subset excluding allo-HCT patients for OS (left panel) and best response (right panel).

Again, inconsequential differences were noted between the IAS-based analyses and that of the FAS analyses (*Supplemental Figure 6B*). Together, these observations demonstrate that the RRM effectively separated Adverse, Intermediate, and Favorable risk groups for both OS and BR, performed well whether allo-HCT recipients were included or excluded, also performed well with a modest degree of data missingness, was adaptable to a version of the dataset with imputed data.

### Comparison of the RRM to ELN22 and mPRS risk stratification models

Next, numerical performance of the RRM was compared to two competing AML risk stratification approaches, the ELN22 and mPRS ^17, 22^. The application of ELN22 model to the CU dataset has been described previously ^27^. In pairwise comparison between the risk subgroups defined by the RRM and ELN22, the main differences were noted in the Adverse risk subsets with the RRM Adverse subgroup having lower median OS (120d vs 206d) and shorter OS behavior than that of the ELN22 Adverse group (Figure 4A; LR *P*-value 0.0037). Fleiss kappa analysis, a measurement of conformity between subject-level risk assignments within risk groups between the RRM and ELN22, demonstrated the least conformity (0.20; *P*-value <0.001) in the Intermediate risk groups (Figure 4B). These observations suggest that many patients classified as Adverse risk group by the ELN22 were stratified as Intermediate group by the RRM.

**Figure 4.**
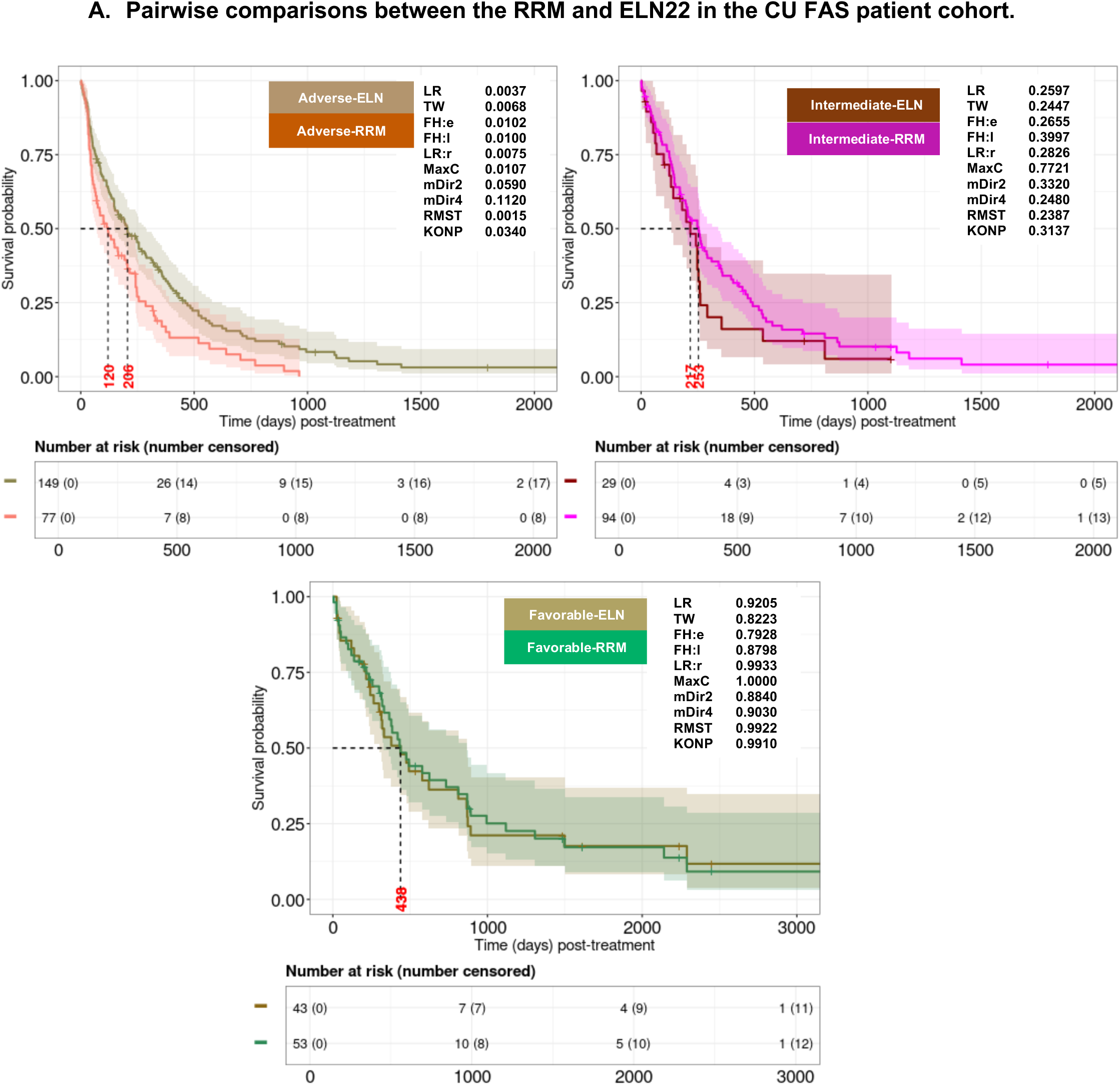
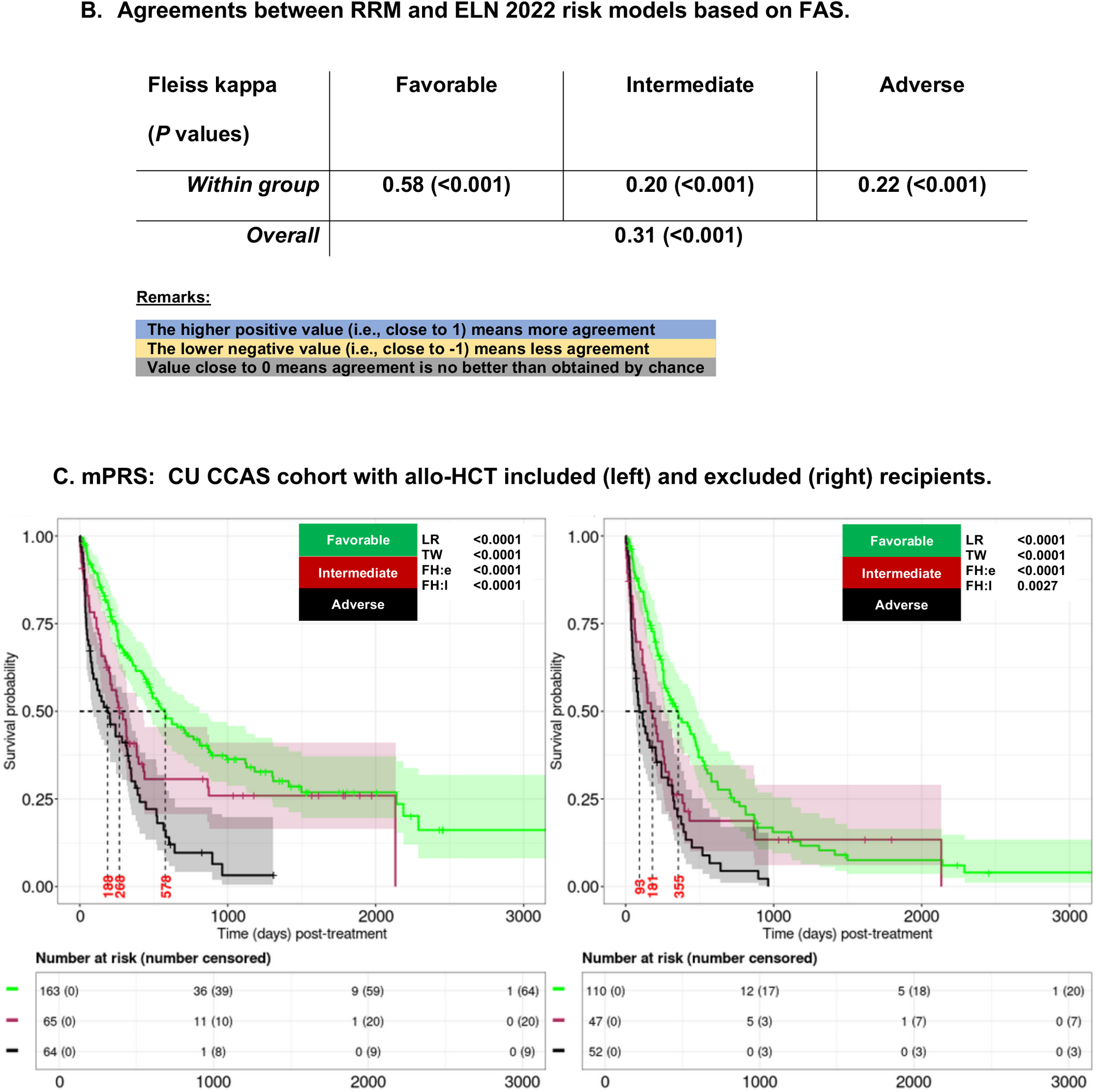
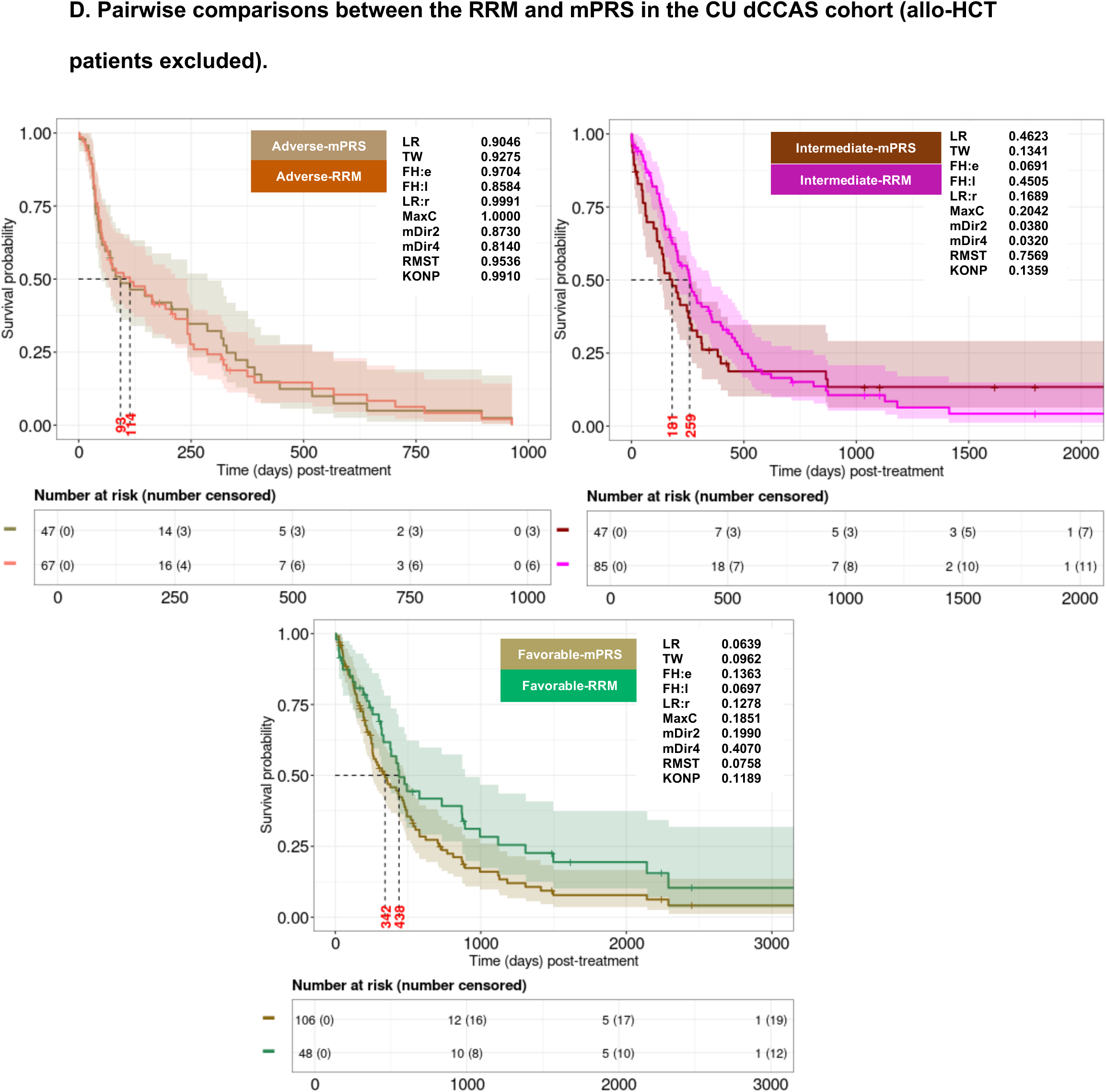
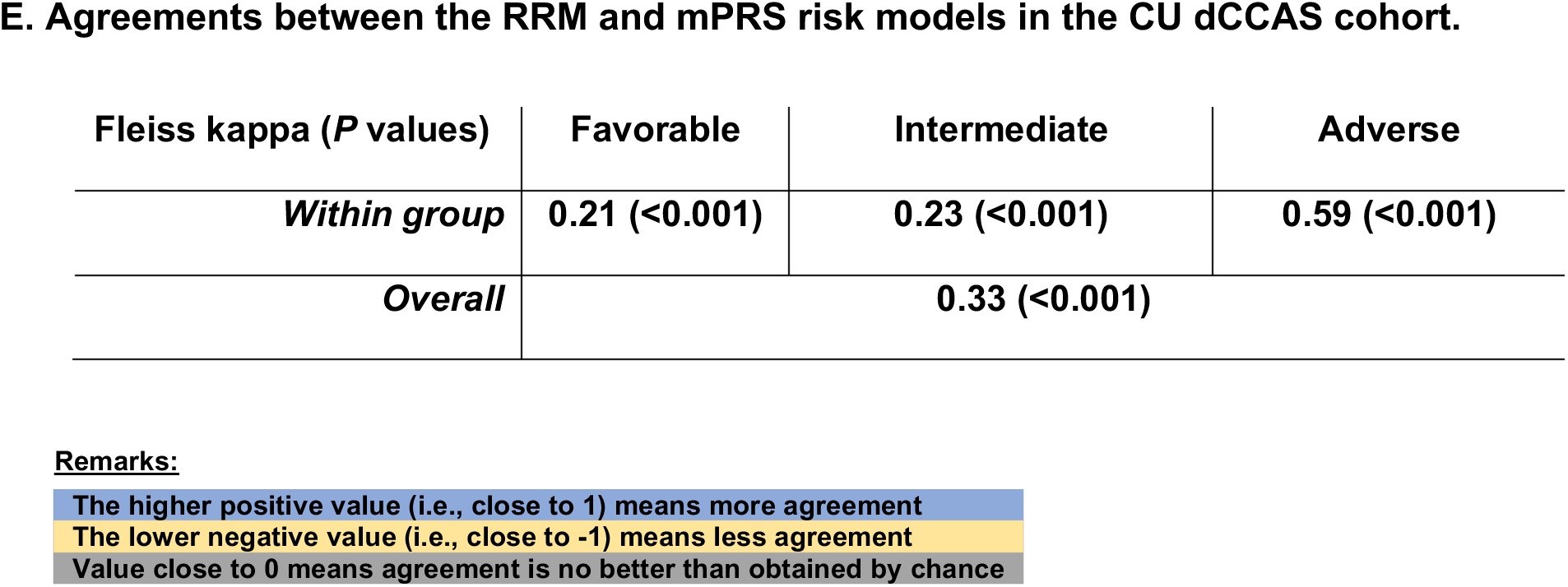
Comparison of the RRM with ELN22 and mPRS risk models in the CU cohort. A) Pairwise comparison between the RRM and the ELN22 model for Adverse, Intermediate, and Favorable risk groups using the CU FAS cohort; B) Agreements of patient assignment to the different risk groups by RRM and ELN22 in the CU FAS cohort; C) Application of the mPRS risk model to the CU Complete Case Analytical Set (CCAS, i.e. the subset of data for which complete data was available for the mPRS) for OS with allo-HCT patients included (right panel) and excluded (left panel); D) Pairwise comparison of the RRM and mPRS risk models using the CU doubly Complete Case Analytical Set (dCCAS, i.e. the subset of data for which complete data was available for both the RRM and the mPRS) for OS with allo-HCT patients excluded; E) Agreement of patient assignment to the different risk categories between the RRM and mPRS in the CU dCCAS using Fleiss kappa.

Next, the RRM was compared to the mPRS model using the CU cohort. First the mPRS was directly applied to the CU cohort and its numerical performance was evaluated. The mPRS model has no obvious provision for assigning patients to risk groups if they are missing any of the four genes used in this model, (i.e., *TP53*, *NRAS/KRAS*, or *FLT3-ITD*). To account for this, the mPRS was initially tested using the CCAS as this most closely parallels the published approach where only cases with complete data for all four genes were used ^20^. All analyses were performed on the CU subsets with allo-HCT recipients included and allo-HCT recipients excluded as above. The mPRS assigned many patients to the High Benefit group (∼56%:22%:22% for three risk groups respectively) as previously reported. The results demonstrated some compression of Intermediate (median OS: 268d) and Low (median OS: 188d) Benefit OS curves at early time points and overlap of Intermediate and High benefit OS curves at later time points after 500 days (Figure 4C) ^20, 22^. Next, pairwise comparisons of the RRM and mPRS models were performed. Note, these pairwise comparisons were performed using a doubly Complete Case Analytical Set termed dCCAS (i.e., the dataset with complete features for both RRM and mPRS) to harmonize the data as closely as possible. The largest OS differences were noted in the Favorable/High Benefit risk groups with higher median and long-term OS for RRM than that of mPRS (Figure 4D, lower panel; median OS of 438d vs 342d respectively). Fleiss kappa analysis also demonstrated the least conformity (0.21; *P*-value <0.001) in the Favorable groups (Figure 4E). To test the impact of missingness on the mPRS, it was tested using the CU FAS as well as IAS analytic datasets as described above. Note, in the CU FAS cohort tested by the mPRS, patients without data on *TP53*, *NRAS/KRAS*, or *FLT3-ITD* status were assigned to the High benefit group as this was where mPRS assigns patients without mutations in these genes. In both the FAS and IAS, the mPRS performed similar to that of the CCAS suggesting that modest degrees of data missingness did not affect numerical performance but again demonstrating possibly reduced *separability* of OS curves relative to the RRM (*Supplemental Figures 7A and 7B*) ^30^. Lastly, empirical performance was assessed for e- mPRS, which included both NGS and additional molecular testing for the same 4 feature genes as defined in Supplemental Table 3. The e-mPRS performed similarly to the mPRS model in the different analytical datasets (*Supplemental Figures 7C-F*) suggesting that additional data types other than NGS may be useful in populating the mPRS.

### External testing of the RRM

To test the generalizability of the RRM, it was applied to the RWC FAS that included and excluded allo-HCT recipients (Figure 5A, left and right panels, respectively). As with the CU cohort, the RRM assigned the largest proportion of patients to the Intermediate category and effectively stratified Adverse, Intermediate, and Favorable OS risk groups for both allo-HCT included and excluded subsets. To test for the impact of data missingness, the RRM was next tested as above on the RWC CCAS which contained 181/971 (∼19%) complete cases in the allo-HCT included and 170/911 (∼19%) in the allo-HCT excluded subsets. Again, the RRM effectively separated the Adverse, Intermediate, and Favorable risk groups in the RWC CCAS (Figure 5B; median OS of 977d, 411d, and 255d for three groups respectively in the set including allo-HCT recipients; LR:*P*-value <0.0001). Similar performance was noted with the RWC IAS as well (*Supplemental Figure 8A*; LR:*P*-value <0.0001). BR testing with the RWC was not feasible due to the absence of short-term response data. Next, comparative analyses between the ELN22 and RRM were performed using the RWC. The ELN22 risk stratification was available for 704/971 (∼73%) and 658/911 (∼72%) patients in the RWC including and excluding allo-HCT recipients, respectively. As with the CU cohort, most patients in the RWC were assigned by the ELN22 to the Adverse risk group (∼65% patients). Separation between the Favorable and Intermediate groups did not appear as robust by the ELN22 as with the RRM model, and attenuation of the Intermediate and Adverse groups at early time points was noted (*Supplemental Figure 8B*, left and right panels). Direct pairwise comparison between the RRM and ELN22 in the RWC demonstrated lower OS for both the RRM-specific Adverse (LR:*P*-value 0.0020) and Intermediate (LR:*P*-value 0.0038) groups (*Supplemental Figure 8C* top panels).

**Figure 5.**
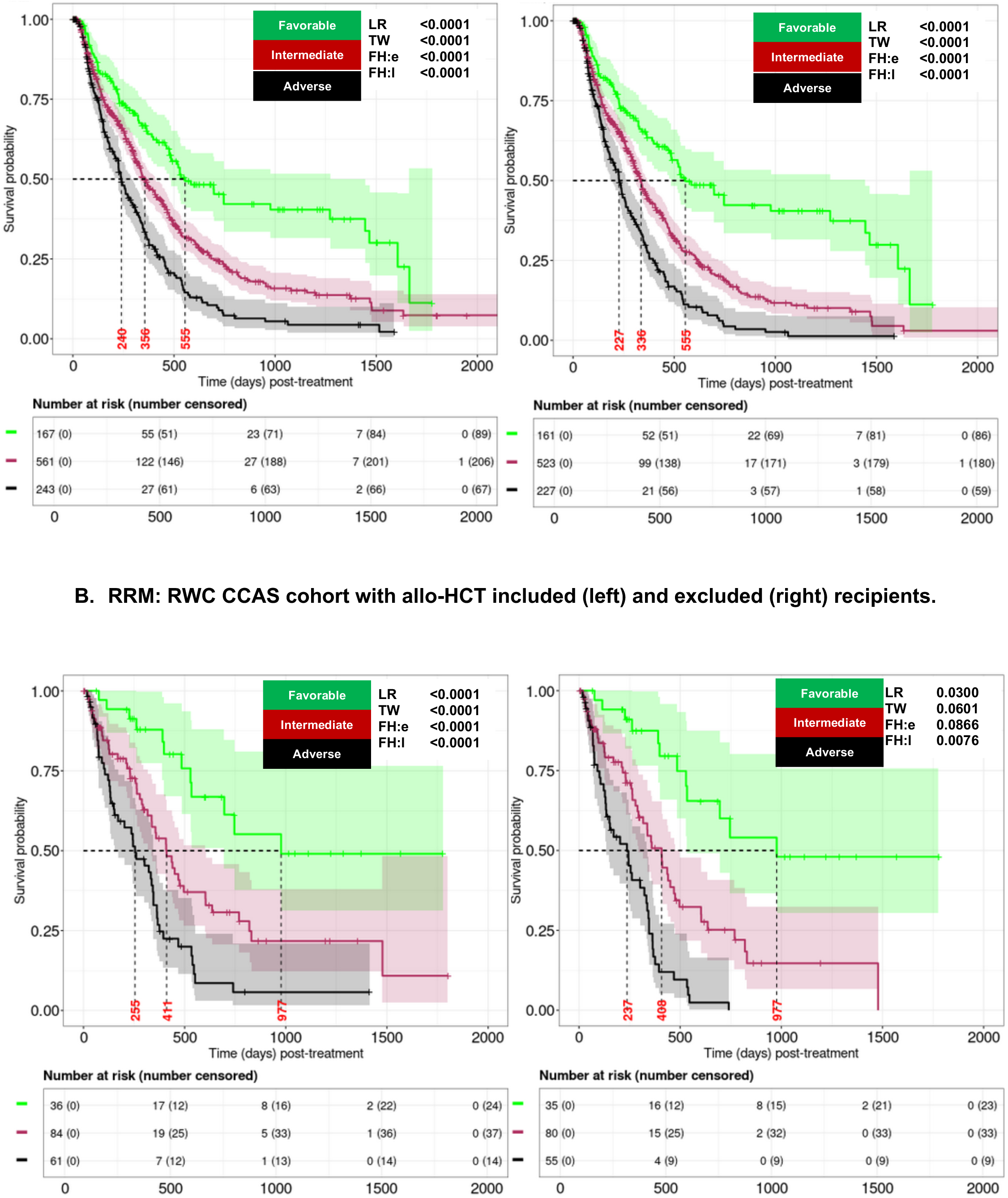
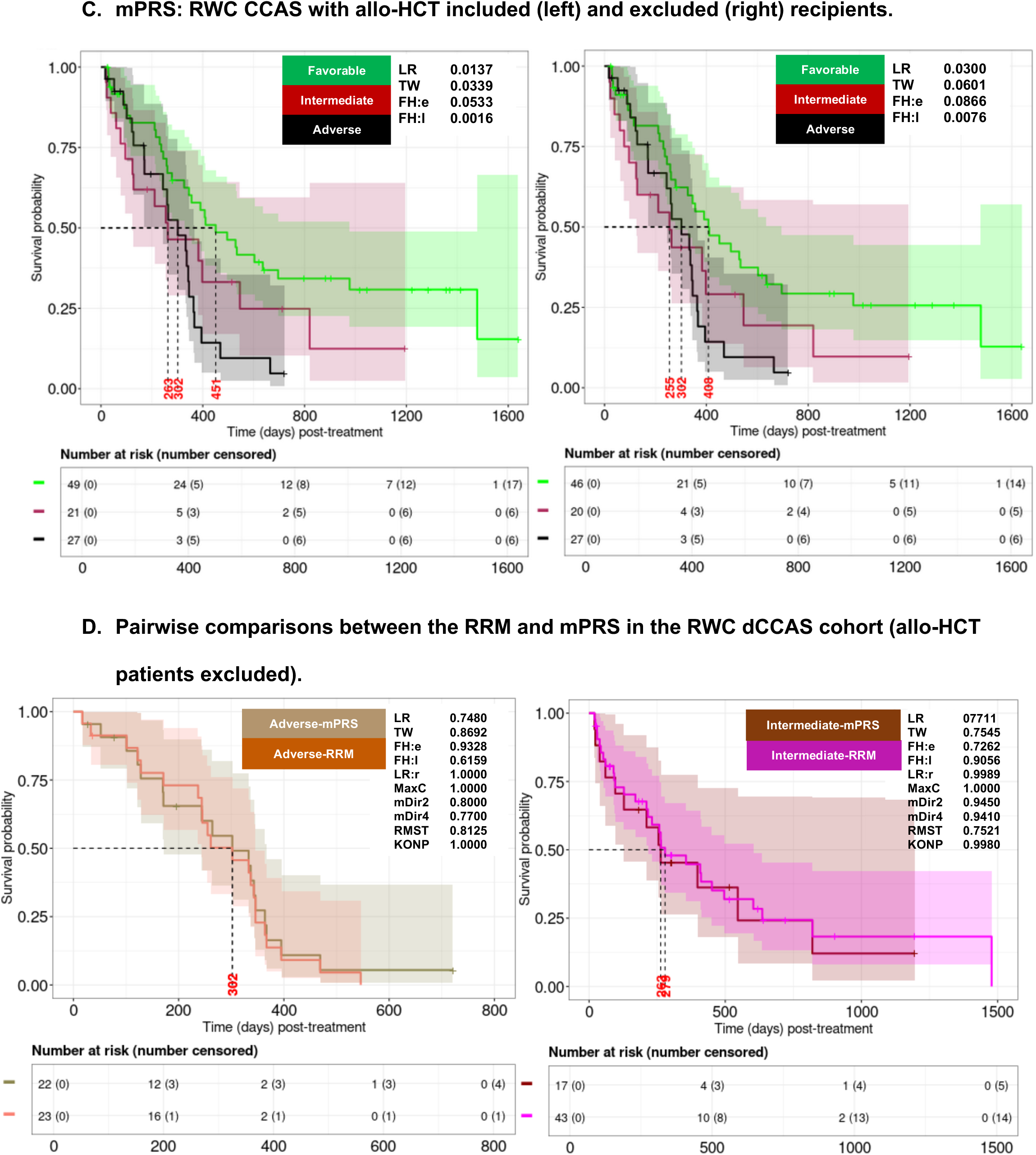
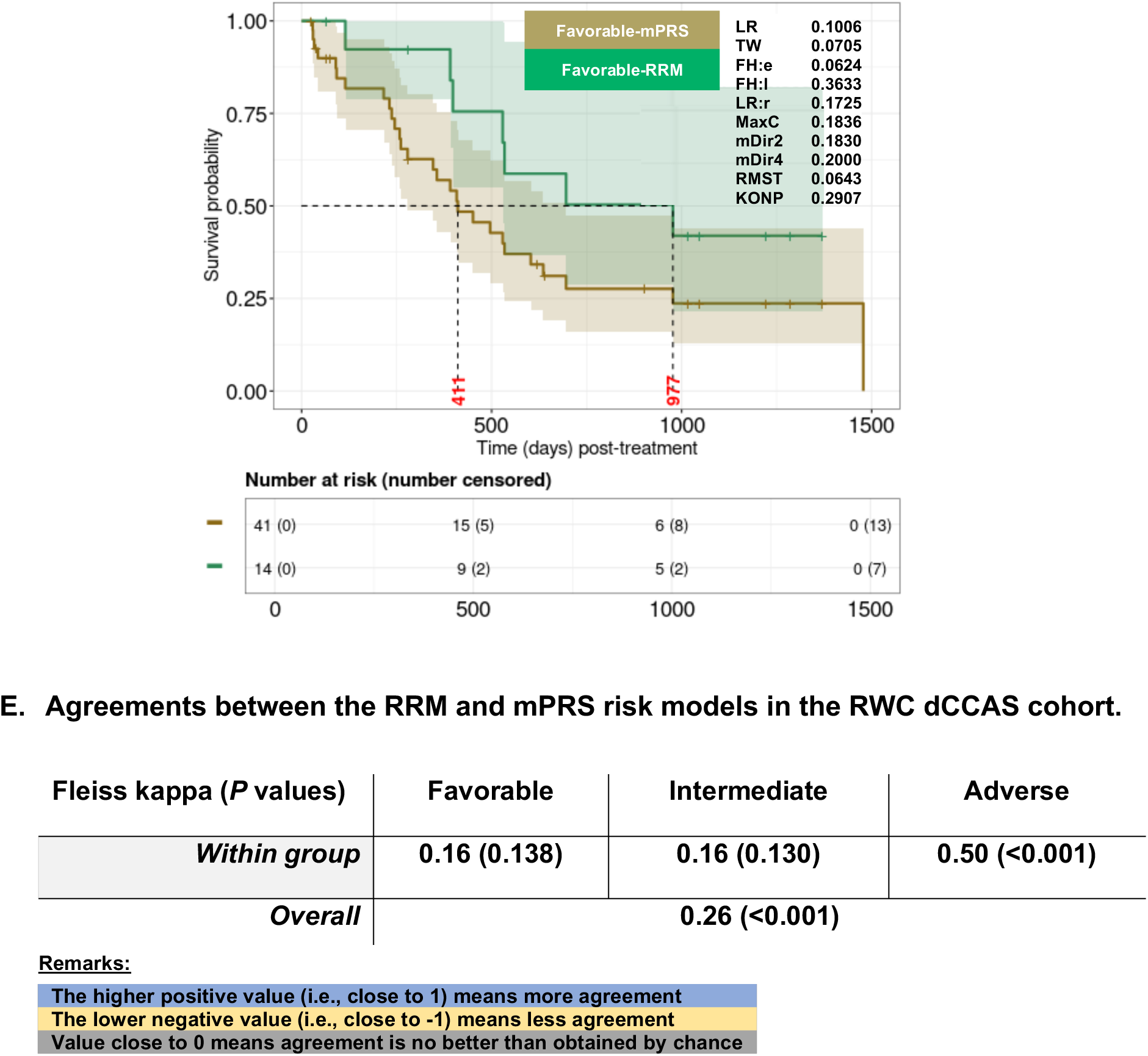
Evaluation of the RRM and mPRS for overall survival using the RWC dataset. A) Application of the RRM to the RWC FAS cohort for OS including allo-HCT patients (left panel) and excluding allo-HCT patients (right panel); B) Application of the RRM to the RWC CCAS cohort for OS including allo-HCT patients (left panel) and excluding allo-HCT patients (right panel); C) Application of the mPRS to the RWC CCAS cohort for OS including allo-HCT patients (left panel) and excluding allo-HCT patients (right panel); D) Pairwise comparison between the RRM and the mPRS model in the RWC dCCAS cohort for Adverse, Intermediate, and Favorable risk groups; E) Agreements across risk groups by RRM and mPRS in the RWC dCCAS.

Fleiss kappa analyses demonstrated the least agreement (0.13; *P*-value 0.001) in the Favorable risk group assignments despite their similar OS behavior (*Supplemental Figure 8D*).

The mPRS model was then tested on the RWC as above. The mPRS was first applied to the RWC CCAS, which had complete data available for the mPRS for 97/971 (∼10%) in the allo-HCT included and 93/911 (∼10%) in the allo-HCT excluded subsets. As with the CU cohort, the mPRS assigned many patients to the Favorable/High benefit cohort (∼50%:22%:27% in three risk groups, respectively in Figure 5C left panel). While the mPRS separated the RWC CCAS subgroups at initial timepoints, crossing patterns in the Intermediate and Low Benefit/Adverse OS curves at later time periods were noted (Figure 5C). Next the mPRS was applied to the RWC FAS cohort (Supplemental Figure 9A, left panel). Limited separation between the High and Intermediate OS curves was noted (median OS 342d in *Supplemental Figure 9A*, left panel). Improved numerical performance (*separability*) was observed for the mPRS based on the RWC IAS (*Supplemental Figure 9A*, right panel). Lastly, pairwise comparisons between the RRM and mPRS were conducted in the RWC using the dCCAS as above (Figure 5D and 5E). Since the number of available patients in the dCCAS was relatively small, these results need to be interpreted with caution; but the biggest survival differences between the RRM and mPRS were noted in the Favorable/High Benefit group (977d vs 411d) where there was also less (0.16) conformity by Fleiss Kappa. When the e-mPRS was tested with the RWC, it demonstrated its best performance in the CCAS and IAS sets (*Supplemental Figure 9B*).

Pairwise comparisons of the RRM and e-mPRS largely paralleled that of the mPRS comparison having the least concordance in the Intermediate groups (*Supplemental Figures 9C-9D*).

Together these findings confirm the consistency of robust numerical performance of the RRM with an external dataset as well as performance comparable to or exceeding the ELN22 and mPRS risk models.

### Predictive comparisons among the RRM, ELN22, mPRS and e-mPRS

Lastly, *predictability* of the RRM, ELN22, mPRS and e-mPRS risk models was assessed at multiple longitudinal follow-up time points by determining cAUC values. First, internal validation for *predictability* over time was performed using cross-validation based on the CU dataset as described in *Supplemental Section A.2*. Treating allo-HCT patients as censored, the CU FAS and an additional dataset termed the total Complete Case Analytic Set (tCCAS, i.e., complete for all variables in all models) were used for predictive evaluation of the RRM, ELN22, mPRS, and e-mPRS. The RRM (cAUC_15_:0.68) demonstrated superior numerical performance over time and at 15 months post-treatment relative to the ELN22 (cAUC_15_:0.52), mPRS (cAUC_15_:0.59), and e-mPRS (cAUC_15_:0.59) (Figure 6A). Similar results were observed in the tCCAS based analyses (Figures 6B-C). Subsequently, using the similar intuition as above, comparative analyses were performed using the external RWC dataset. As before, the RRM exhibited superior predictive performance among the competing models (Figures 6D i-iv). Similar results were noted using the RWC FAS and IAS datasets although the m-PRS and e-mPRS performed like that of the RRM with the IAS set (*Supplemental Figure 10*). Together these findings suggest comparable or superior *predictability* performance of the RRM relative to the ELN22 and mPRS/e-mPRS across the CU and RWC.

**Figure 6.**
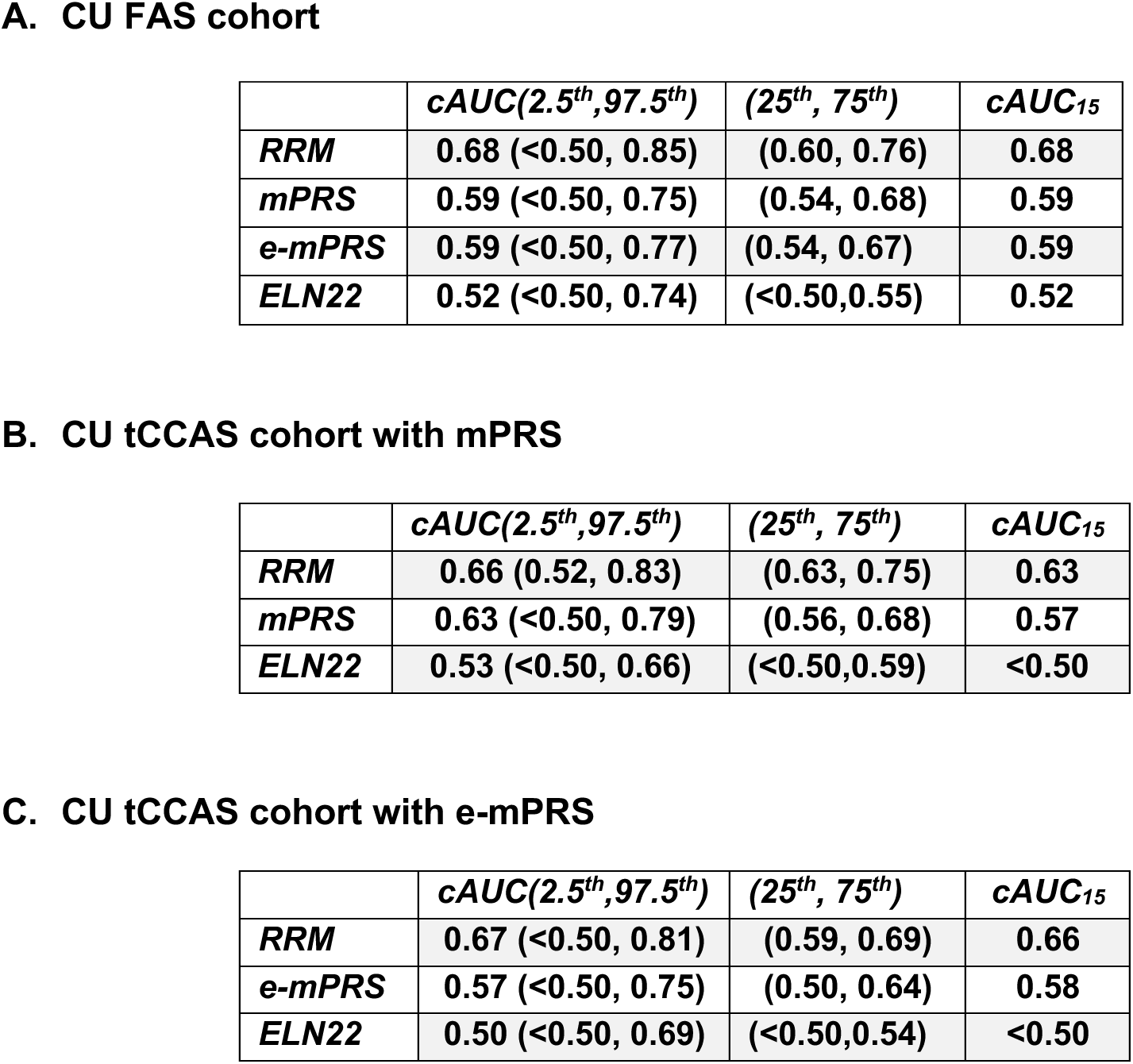
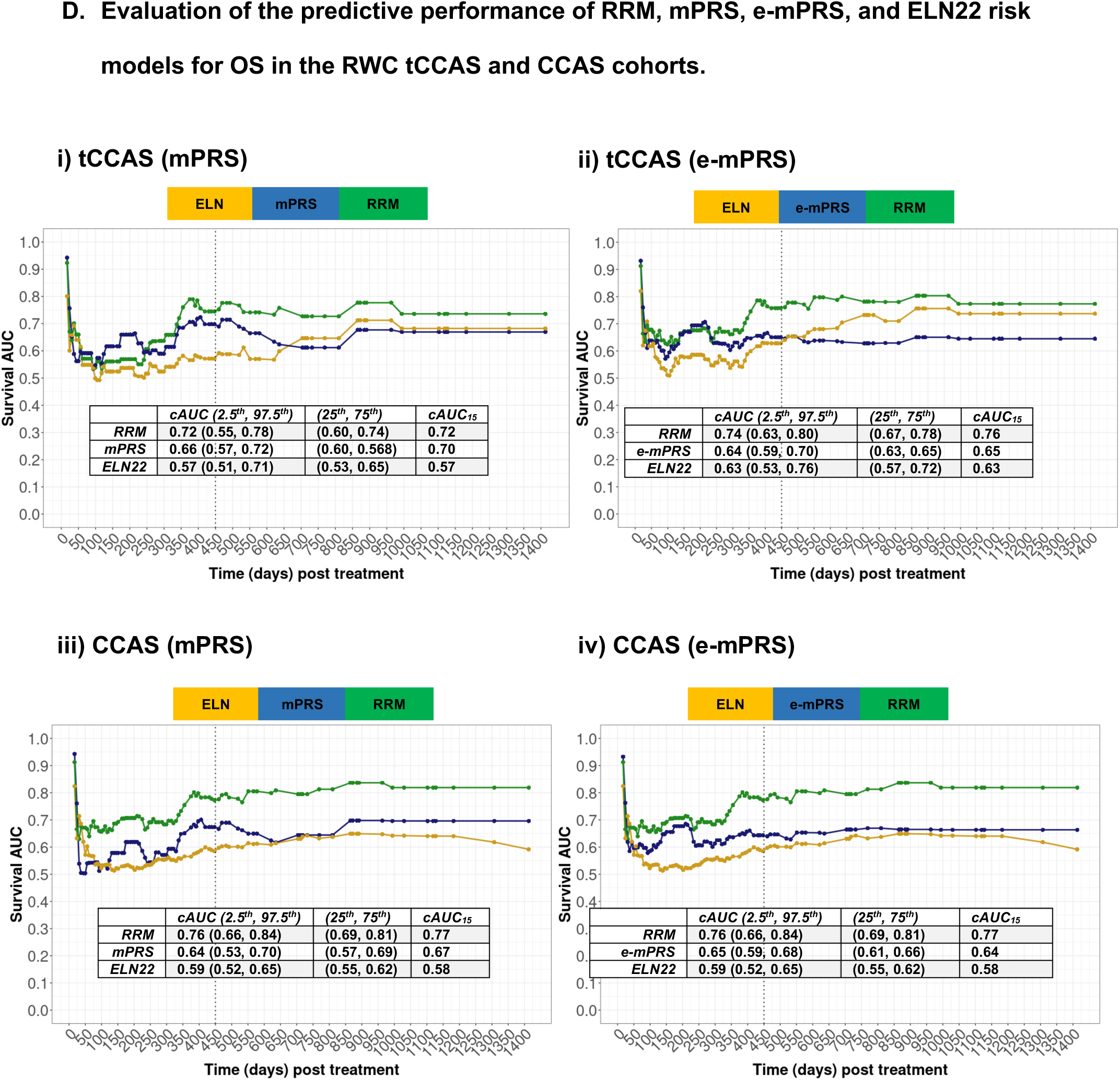
Predictive validation of the RRM, mPRS and ELN22 models using CU and RWC datasets. Predictive analysis was performed using the allo-HCT as censored subset. Results are summarized over 15-fold cross-validation (CV) over unique follow-up times up to 4 years based on penalized Cox-PH model adjusting for age, gender, race, and either RRM, mPRS, e-mPRS, or ELN22 risk variables for the A) CU FAS, B) the CU total Complete Case Analytical Set (i.e., tCCAS, the subset of patients with complete ELN, mPRS and RRM data) for the mPRS and C) the tCCAS for the e-mPRS. Reported are the medians (over CVs) of 2.5^th^, 25^th^, 50^th^, 75^th^, and 97.5^th^ percentile values of AUCs with respect to cumulative case/dynamic control (cAUC) receiver operator curves over time. Evaluations are enumerated at discrete unique follow-up times that are at-least 5-days apart (as described In the Methods); cAUC_15_ refers to cAUC value at 450 days (∼15 months). D) Evaluation of predictive performance of the RRM, mPRS, e-mPRS, and ELN22 risk models for OS up to 4 years in the tCCAS and CCAS of the RWC (allo-HCT censored) based on penalized Cox-PH model. Models were trained on four different complete case (CC) scenarios. i) tCCAS with mPRS; ii) tCCAS with e-mPRS; iii) CCAS with mPRS; iv) CCAS with e-mPRS.

## Discussion

In this study, we developed the RRM as a relatively simple upfront stratification approach for newly diagnosed AML patients specifically treated with ven/aza (see Figure 7 for a summary of goals and considerations). The RRM was based on diagnostic AML genetic tests that are potentially readily-and-widely available, and effectively stratified patients into Adverse, Intermediate, and Favorable risk groups of relatively balanced group sizes with distinct OS and BR behavior. The RRM performed favorably in the subsets that included and excluded allo-HCT recipients, tolerated feature identification using a variety of genetic testing technologies, and effectively stratified patients in both the CU and the multi-institutional RWC cohorts. Additionally, the RRM was designed to manage data missingness which is an important consideration in prospective stratification of patients when complete data is unavailable in the requisite time frame. Missingness is typically dealt with by either discarding patients who have incomplete data or imputing missing instances from observed data ^31^. However, as the magnitude of missingness goes up, discarding or imputing data may introduce selection bias confounding up front patient stratification and interpretation ^32–34^ ^35^. The robust performance of the RRM in managing data missingness in both the CU and RWC may help address these concerns.

**Figure 7.**
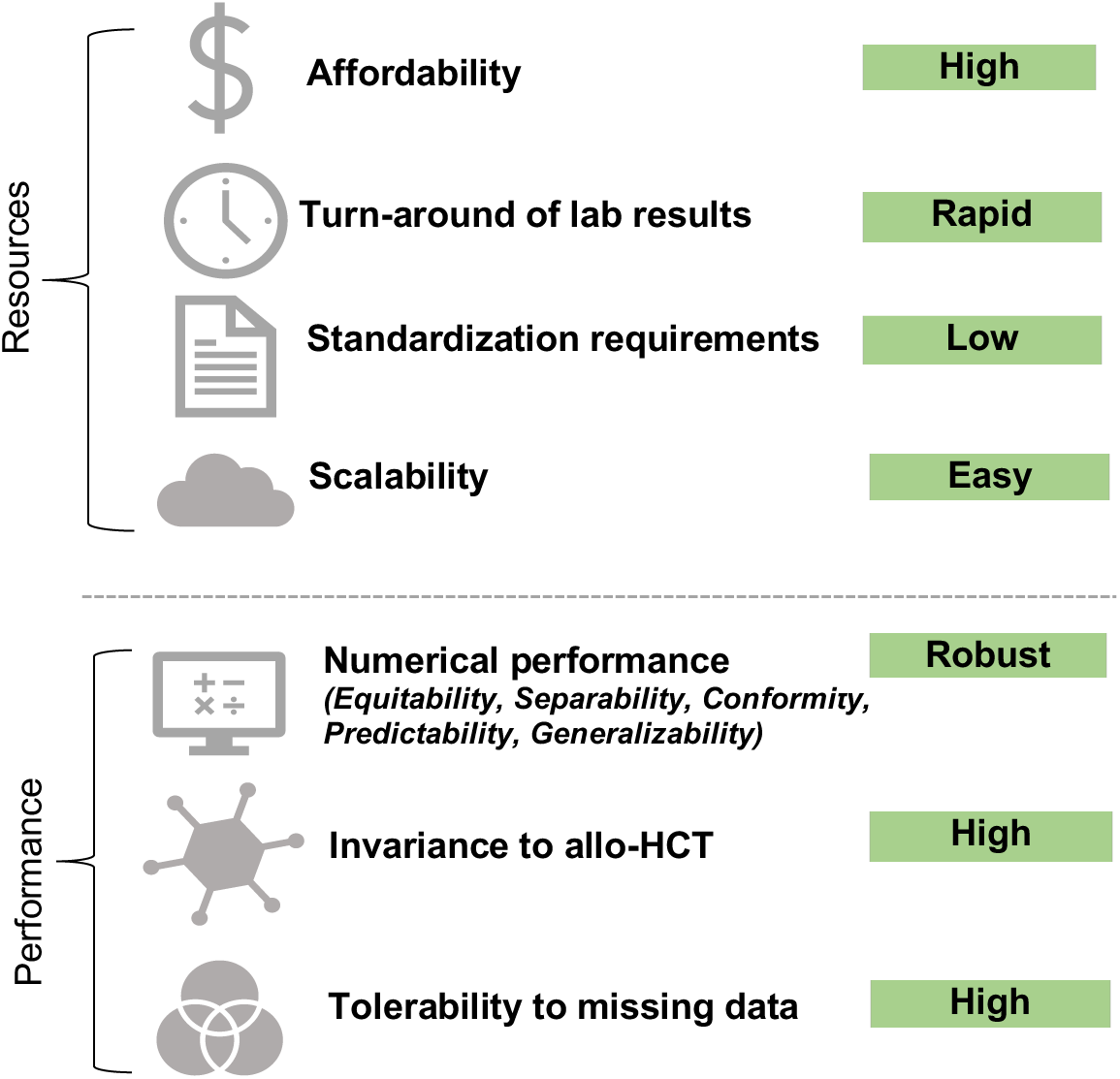
Goals and considerations in developing and testing the RRM. The RRM was developed to be a relatively simple, affordable and widely applicable risk stratification model with the ability to perform well with a variety of testing types, data missingness, variable inclusion of subsequent allo-HCT recipients and other real-world considerations important in an upfront patient stratification strategy.

Additionally, relative to the ELN22 and mPRS, the RRM had comparable or favorable performance based on *equitability, separability, conformity, predictability*, and *generalizability* across both the CU and RWC datasets. Of note, the mPRS performs well with complete datasets confirming its utility in the appropriate settings. The mPRS also outperformed the ELN22 here, adding further evidence that the ELN22 is not ideal for ven/aza or ven/HMA treated patients ^19–22, 30^.

Several additional observations were made. First, the confounding effect of allo-HCT to OS should be considered in future studies as varying frequencies of allo-HCT recipients across different cohorts may impact both median OS and long-term OS, particularly in relatively small datasets, as also reported by others ^19^. However, excluding or censoring allo-HCT patients may also introduce selection or attrition biases. Moreover, allo-HCT is an important and growing therapeutic option for ven/aza treated patients and thus, necessitates careful consideration. To provide a comprehensive and balanced understanding of the effects of ven/aza treatment on survival outcomes, numerical results may need to be presented for multiple patient cohorts: the FAS including all patients, the set excluding allo-HCT recipients, the set treating all-HCT recipients as censored, and the set of allo-HCT recipients alone as we have previously described ^27^. The imputed models utilized here exhibited varying levels of numerical performance – which is not surprising as it depends on factors including, but not limited to, the missingness pattern, magnitude of missingness, and functional forms of the imputed models.

Therefore, using imputation as a method for managing missingness should be interpreted with caution. Lastly, multiple observations related to a non-proportional OS pattern suggest that relying on a single simple summary statistic (e.g., median OS) may be misleading regarding the overall behavior of a particular treatment or cohort subset. As reported here, it may be more informative to report a variety of statistical metrics for both short- and long-term OS behavior to comprehensively interpret survival outcomes.

There are a series of limitations to this study. Because of varied practices in molecular pathology and data reporting for both cohorts as well as data harmonization purposes, the molecular tests used in this study were all treated as equal and in a binary fashion for mutation status, regardless of technology, mutation frequency, type of mutation, or other potential confounding features. Additionally, cytogenetic reporting varied between the CU and RWC datasets as described in *Supplemental Section A.1-A.2*. Differences in test types, technologies, and reporting are likely to be important and further improvements in the RRM are expected when it is trained on larger and more comprehensive datasets that address these important distinguishing features in a more nuanced, harmonized, and consistent fashion. Also, the RRM currently uses features that are available with tests outside NGS; however, as NGS becomes cheaper and more widely available with faster turn-around times, RRM performance may be enhanced by incorporating additional data and features that are only available through NGS. The RRM includes both cytogenetic and molecular features as exploratory analyses using univariate and multivariate methods demonstrated that these features were not completely overlapping. However, there are currently contrasting observations on whether *TP53* abnormalities and poor risk cytogenetics are possible non-overlapping risk factors for ven/aza treated AML patients, so further study of this issue is important ^19, 30, 36^. Additionally, several risk features in the RRM have low prevalence rates and require additional validation to confirm that these features are not merely selected by chance. Further refinement of the RRM, particularly by parsing Intermediate risk features into Adverse and Favorable risk categories while avoiding creation of unacceptable levels of complexity will also be important. The absolute median OS in this report varied between the CU and RWC datasets by several months, in part due to differences in comorbidities. Other possible contributors included differential survival and data reporting, different patterns of salvage therapies after ven/aza, patient selection bias, site-to-site variations, and differences in data definition and management factors. These types of differences will also be important to control for in future confirmatory studies. The ethnic diversity of both datasets used in this study is limited, and studying populations with more diversity is also necessary to further improve the generalizability and fairness of the RRM ^37^.

Lastly, this report relies on retrospective data with its inherent limitations. It will be critical in future studies to address these caveats through larger, multi-center, diverse, harmonized, and comprehensive datasets as well as through prospective confirmation of the RRM performance^30^.

In conclusion, we developed and validated externally the RRM for risk stratification of newly diagnosed AML patients treated with ven/aza. Further validation of the RRM with additional datasets along with the application of more consistent and standardized diagnostic testing will improve and refine the overall RRM empirical performance as well as its applications to upfront patient risk stratification and retrospective analyses.

## Supporting information

Supplementary Sections

## Data Availability

This retrospective study was approved by CU internal review board (IRB) and used a limited dataset with a waiver of consent from the CU IRB. The raw, individual patient data are protected and not available due to data privacy laws. The processed data are available at reasonable request to the corresponding author. The Flatiron Health data that supported the findings of this study were originated by and are the property of Flatiron Health, Inc., which has restrictions prohibiting the authors from making the data set publicly available. Requests for data sharing by license or by permission for the specific purpose of replicating results in this manuscript can be submitted to.

## ACKNOWLEDGEMENT

The authors gratefully thank Dan Pollyea and Andrew Kent from the School of Medicine, University of Colorado Anschutz Campus, Aurora, Colorado, Bin Yao and Adam George from OncoVerity, Aurora, Colorado and Grant Weller from the RefinedScience, Aurora, Colorado for their feedback and reviews during this project.

## AUTHOR CONTRIBUTIONS

CAS and NI designed the study and drafted the manuscript JSR, JLD, KS, JWC, FRM, and UVK processed and pulled the structured analytical datasets. CAS, NI, JSR, JLD, JZ, KS, JWC, and LW assessed the validity and quality of data. NI and LW performed numerical analyses. CAS, NI, MB, and JSR interpreted the results of the analyses. All authors reviewed, provided constructive comments, and agreed to its publication.

## SUPPLEMENTARY MATERIAL

Supplemental materials contain additional details of methodology and numerical results pertinent to the study.

## ABBREVIATIONS

AML: Acute myeloid leukemia
IC: Intensive chemotherapy
Allo-HCT: Allogeneic hematopoietic cell transplant
ELN: European Leukemia Network
CR: Complete response
Cri: CR with incomplete hematologic recovery
CRh: CR with partial hematologic recovery
MLFS: Morphologic leukemia free state
PR: Partial remission
CYT: Cytogenetics
FC: Flow cytometric
FISH: Fluorescence in situ hybridization
NGS: Next generation sequencing
PCR: Polymerase chain reaction
ven/aza: Venetoclax plus azacytidine
OS: Overall survival
IRB: Institutional review board
CU: University of Colorado
RWC: Real-world cohort based on the Flatiron Health AML database
KM: Kaplan-Meier
LR: Log-rank
TW: Tarone-Ware
FH: Fleming-Harrington
mdir: Weighted multiple direction
MC: Max-Combo
KONP: K-sample omnibus non-proportional hazard
RMST: Restricted mean survival times
AUC: Area under the curve

