## Supplementary Sections for "A simplified risk model for pretreatment stratification of newly diagnosed acute myeloid leukemia patients treated with venetoclax and azacitidine"

#### SUPPLEMENTARY MATERIALS

**Supplementary Methods**

**Supplementary Tables**

**Supplementary Figures**

#### A. Supplemental Methods

This section provides additional details related to data definitions, normalization, standardization, and statistical methodology.

##### A1. Populations, definitions, and standardization

***Analytical patient selection***

The Flatiron Health dataset is a licensed, deidentified dataset containing individual patient data on patients who were diagnosed with AML using ICD-9 and ICD-10 codes; had at least two documented clinical visits on different days occurring between January 2014 and December 2023; and received ven/aza as frontline therapy. During the study period, the de-identified data originated from approximately 280 US cancer clinics (~800 sites of care; primarily community oncology settings). The data were de-identified and subject to obligations to prevent re-identification and protect patient confidentiality. For notational convenience, the real-world cohort derived from the Flatiron Health is denoted by RWC throughout the paper. Patients were excluded from both the CU and RWC datasets who had acute promyelocytic leukemia (APL), blast crisis chronic myeloid leukemia (CML), biphenotypic leukemia or central nervous system (CNS) involvement with AML at diagnosis. In general, as summarized in Figure 1, similar exclusion/inclusion criteria were applied to both the CU and RWC datasets to lessen the risk of systematic differences. Patients' summary statistics were provided along with standardized mean difference (SMD) values highlighting considerable differences in the distribution of AML features between the CU and RWC datasets in Table 1; typically a value > 0.10 signaled a meaningful difference between patient cohorts.

***Outcomes analysis***

In overall survival (OS) analyses assuming right censoring, time-to-event was defined as time from ven/aza treatment initiation to all-cause mortality. Short-term treatment responses including complete response (CR), complete response with incomplete hematologic recovery (CRi), CR with partial hematologic recovery (CRh), morphologic leukemia free state (MLFS), partial remission (PR), no response, and relapse were defined based on ELN22 definitions.

***Accounting for the impact of allo-HCT on outcomes***

Initial KM analysis demonstrated higher OS in the CU cohort compared to that of RWC when allo-HCT recipients were included in the analysis (LR $P$ value < 0.0386), Supplemental Figure 1A). However, the CU cohort contained a considerably larger proportion (29%) of allo-HCT recipients compared to that of the RWC (6%) highlighting the degree of treatment variations. When allo-HCT patients were excluded from both datasets, OS seemed relatively similar, strongly suggesting a confounding impact of allo-HCT on OS. To account for this bias in the RRM development specially at the feature selection step, allo-HCT patients were censored at their first transplant date. Performance assessments were then tested comprehensively on the CU and RWC patient cohorts that both included and excluded allo-HCT recipients, separately. Though not reported here – performance assessments were also conducted on the subset where allo-HCT recipients were censored and the subset consisting of allo-HCT recipients only. Using of such multiple patient cohorts in assessments helped to determine whether the RRM could effectively stratify patients over a range of allo-HCT recipient frequencies and be useful for defining prognosis for most patients treated with ven/aza who would not undergo allo-HCT. Of note, there was a median OS difference of 3-months observed between the allo-HCT excluded CU and RWC subsets (Supplemental Figures 1B and 1C). Approximately one month of this could be accounted for by the comorbidity differences noted in Table 1 with the remaining difference unexplained to date.

***Data standardization and AML genetic testing***

In the CU and RWC datasets, AML genetic testing data for each patient was provided on a per marker basis using NGS, PCR, FISH, cytogenetics or other/unknown test types. Complex cytogenetics was defined as ≥3 karyotype abnormalities in the CU and RWC dataset by counting the karyotype abnormality fields that were available and assigning to complex cytogenetics if there were 3 or more. Monosomal karyotype for ELN adverse risk were not assessed due to a lack of information. Of note, the RWC was populated with 21 karyotype abnormalities whereas the CU dataset contained information on full karyotypes. Good risk cytogenetics were defined as *t(8;21)*, *inv(16)*, *t(16;16)*, or *t(15:17)*. In the RWC, partial chromosomal or full chromosomal deletions are used in their term "Chromosome x deletion" whereas the CU karyotype data distinguishes partial and full deletions or other abnormalities. The term "*TP53m*" used in the RRM was defined as a *TP53* abnormality by NGS at any VAF or by FISH. For ELN22 calculations using the CU dataset, NGS testing for the detection of a *TP53* mutation plus a VAF score >10% was utilized. Otherwise *TP53* mutations in the CU dataset was scored as positive, negative, or not performed for the RRM and mPRS models. For all RWC analyses, *TP53* VAF was not available so was also scored as positive, negative, or not performed. Moreover, bzip in frame mutation for *CEBPA* for ELN favorable risk were not assessed in the RWC due to the lack of information. An extension of mPRS model termed as "e-mPRS" was developed by replacing the NGS-based *TP53* and *FLT3-ITD* mutation features with composite *TP53* and *FLT3-ITD* mutations as defined in Supplemental Table 3.

**A2. Risk model development and validation**

Descriptive statistics (e.g., median, counts, percentages) for baseline covariates of AML patients treated with ven/aza were provided. OS estimates based on Kaplan-Meier (KM) analyses along with 95% confidence intervals (CIs) and median time-to-event were reported. The equality of survival curves were evaluated based on Log-rank (LR), its weighted counterparts (i.e., Tarone-Ware (TW), Fleming-Harrington (FH) with an emphasis on early and late time periods, weighted multiple direction (mdir) LR, Max-Combo (MC)), K-sample omnibus non-proportional hazard (KONP), and restricted mean survival times (RMST) tests; the corresponding *P* values were reported. Equality of proportions of CR/CRh among strata was tested via Fisher-exact or Chi-Squared test. A parsimonious list of features was obtained as candidate input variables for the final multivariate training model where noise variables were screened out using univariate filtering approach based on the null hypothesis significance testing with Type-I error rate setting equal to 0.30.

The refined risk stratification model (RRM) was developed using a penalized Cox-PH model with L2 norm (ridge) penalty (mCOXr) coupled with S-learning technique and counterfactual arguments following the procedures described in a separate study [27]. The stratification in the RRM is primarily comprised of two steps: 1) covariate-level risk classification and 2) subject-level risk stratification. A genetic-features specific risk model ($RM_G$) for OS based on the Rule-I as previously described was used as an empirical foundation for the covariate-level classification in the RRM, which was further augmented by clinical intuition, practicality, and published data [1]. The RRM was trained treating allo-HCT recipients as censored at their first transplant date and assuming missing data as a separate category. Covariate-level classification was conducted using 5% risk differences (RD) at 14.7 months (the median OS in the pivotal VIALE-A study) between marginal risk profiles of mortality for wild-type and mutated genes. Subsequently, subject-level risk stratification classified each patient into Adverse, Intermediate, or Favorable risk group depending on the presence or absence of at-least one Adverse or Favorable features.

Comparisons between the RRM vs ELN22 and RRM vs mPRS models using both the CU and RWC datasets were performed using several complementary approaches. In particular, the comparative analyses were based on: 1) **F**ull **A**nalytical **S**et (FAS): the set of all patients. Note that at the patient-specific risk stratification step, missing values for a feature were imputed by mode of the corresponding feature (i.e., imputing values with the most prevalent class label for the corresponding feature); 2) **D**oubly **C**omplete **C**ases **A**nalytical **S**et (dCCAS): the subset of patients with complete data for the RRM and mPRS or e-mPRS models; 3) **T**otal **C**omplete **C**ases **A**nalytical **S**et (tCCAS): the subset of patients with complete data for the RRM, mPRS/e-mPRS, ELN2022 models; and 4) **I**mputed **A**nalytical **S**et (IAS): the pseudo-set of all patients where missing values for a patient were imputed with values computed via multivariate imputation by chained equations (MICE) approach for 10 times based on a combination of the observed CU and RWC dataset (N = 1,287 (316 CU and 971 RWC)). Note that the comparative analyses were performed independently on each IASs and the reported numerical results for *equitability*, separability, and *conformity* were based on a randomly chosen one imputed dataset.

Internal *validation* of *predictability* based on the CU dataset was performed by 15-fold cross-validation. Dynamic survival AUC values were calculated at unique follow-up times at-least 5-days apart up to 4-years for both datasets. Survival cAUC values at $2.5^{th}$, $50^{th}$ (median), $75^{th}$, and $97.5^{th}$ percentile over time were reported. For the internal validation steps, median cAUC values were summarized over 15 CV folds. For the external validation with respect to FAS, CCAS, and tCCAS, the trained models based on the CU analytical sets were applied and tested on the corresponding RWC datasets (FAS, CCAS, and tCCAS). For the evaluation of *predictability* with respect to IASs, test results were averaged over 10 imputed RWC sets.

#### SUPPLEMENTAL TABLES

##### Supplemental Table Legends

**Supplemental Table 1. Definition of patient data sets and applicability for risk model performance testing.**

**Supplemental Table 2. Definition of comorbidity variables by ICD-10 codes.**

**Supplemental Table 3. Definitions of AML mutation variables.**

**Supplemental Table 1. Definition of patient data sets.**

| | |
|---|---|
| **FAS** | **F**ull **A**nalytical **S**et. Note that at the patient-specific risk stratification step, missing values for a feature were imputed by mode of the corresponding feature (i.e., imputing values with the most prevalent class label for the corresponding feature). |
| **IAS** | **I**mputed **A**nalytical **S**et. Multiple imputations (10 imputed samples) were generated for each missing data element using multivariate imputation by chained equations (MICE) approach. |
| **CCAS** | **C**omplete **C**ase **A**nalytical **S**et after excluding missingness of the underlying variables that were used in defining the respective risk stratification variable (i.e., mPRS, e-mPRS, RRM). In the analyses with ELN, patients for who ELN 2022 variable was not feasible to calculate due to unavailability of information were excluded. |
| **dCCAS** | **D**oubly **C**omplete **C**ase **A**nalytical **S**et after excluding missingness of the underlying variables that were used in defining two risk stratification variables (i.e., mPRS and RRM or e-mPRS and RRM). In the analyses with ELN, patients for who ELN 2022 variable was not feasible to calculate due to unavailability of information were excluded. |
| **tCCAS** | **T**otal **C**omplete **C**ase **A**nalytical **S**et after excluding patients for who ELN 2022 variable was not feasible to calculate and excluding missingness of the underlying variables that were used in defining any other risk stratification variables (i.e., mPRS and RRM or e-mPRS and RRM) |

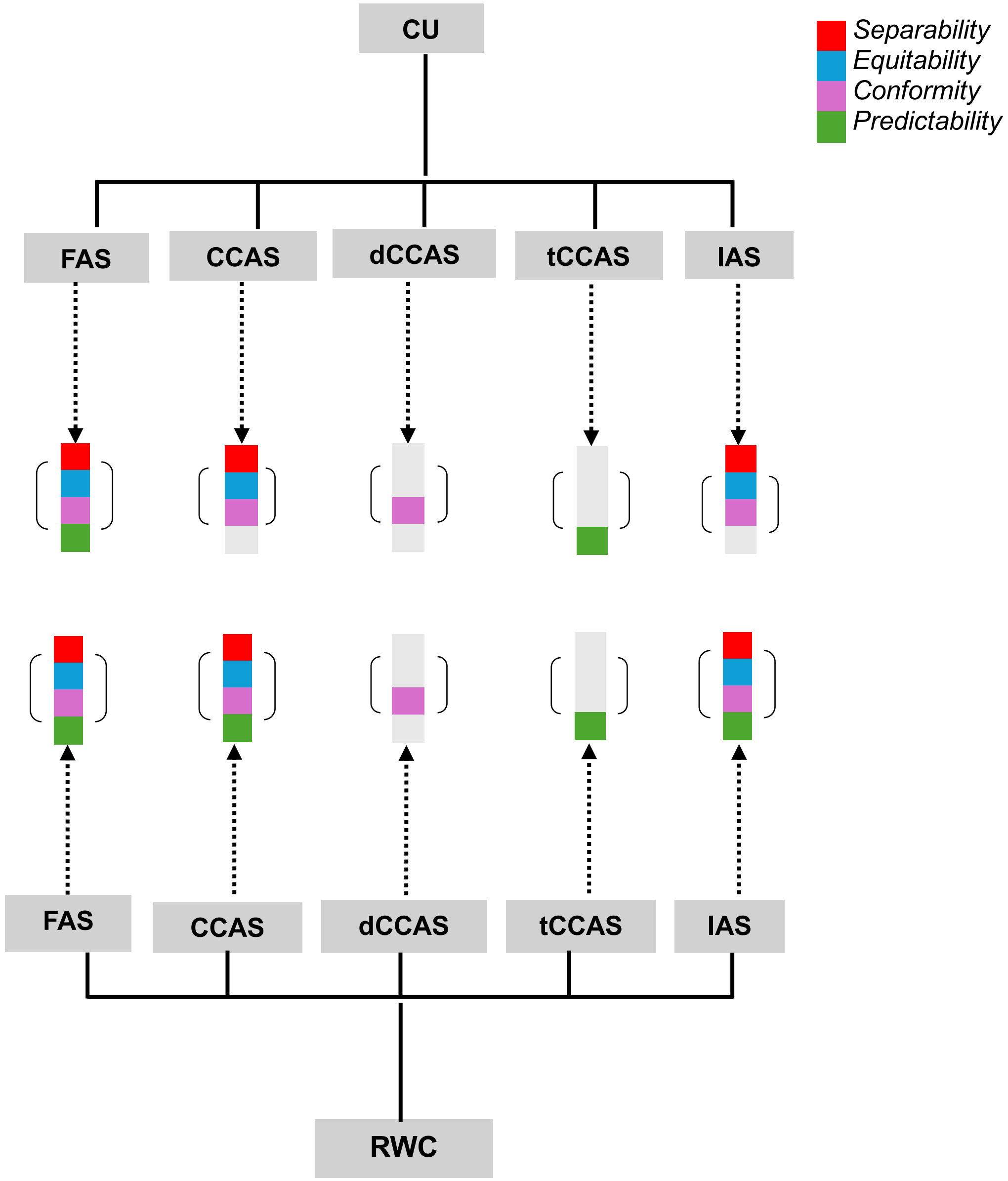

**Supplemental Table 2. ICD-10 codes for comorbidity variables.**

| *ICD-10* | *Descriptions* | |
|---|---|---|
| *I120.9* | Angina pectoris, unspecified | Heart disease |
| *I121.3* | ST elevation (STEMI) myocardial infarction of unspecified site | |
| *I125.1* | Atherosclerotic heart disease of native coronary artery | |
| *I125.2* | Old myocardial infarction | |
| *I125.84* | Coronary atherosclerosis due to calcified coronary lesion | |
| *I125.9* | Chronic ischemic heart disease, unspecified | |
| *I125.10* | Atherosclerotic heart disease of native coronary artery without angina pectoris | |
| *I3%* | Acute pericarditis | |
| *I40%* | Acute myocarditis | |
| *I41%* | Myocarditis in diseases classified elsewhere | |
| *I42%* | Cardiomyopathy | |
| *I43%* | Cardiomyopathy in diseases classified elsewhere | |
| *I44%* | Atrioventricular and left bundle-branch block | |
| *I45%* | Other conduction disorders | |
| *I47%* | Paroxysmal tachycardia | |
| *I48%* | Atrial fibrillation and flutter | |
| *I49%* | Other cardiac arrhythmias | |
| *I50%* | Heart failure | |
| *I51%* | Complications and ill-defined descriptions of heart disease | |
| *I52%* | Other heart disorders in diseases classified elsewhere | |
| *J44%* | Other chronic obstructive pulmonary disease | COPD |
| *I82.4%* | Acute embolism and thrombosis of deep veins of lower extremity | VTE |
| *I82.5%* | Chronic embolism and thrombosis of deep veins of lower extremity | |
| *I82.62%* | Acute embolism and thrombosis of deep veins of upper extremity | |
| *I82.72%* | Chronic embolism and thrombosis of deep veins of upper extremity | |
| *I26.0%* | Pulmonary embolism | |
| *I26.9%* | Pulmonary embolism without acute cor pulmonale | |
| *I27.82* | Chronic pulmonary embolism | |
| *N17%* | Acute kidney failure | CKD |
| *N18%* | Chronic kidney disease (CKD) | |
| *N19%* | Unspecified kidney failure | |
| *C0%* | Malignant neoplasm of: lip, base of tongue, other and unspecified parts of tongue, gum, floor of mouth, palate, other and unspecified parts of mouth, parotid gland, other and unspecified major salivary glands, tonsil | Non-AML cancer |

| | | |
|---|---|---|
| C1% | Malignant neoplasm of: lip, base of tongue, other and unspecified parts of tongue, gum, floor of mouth, palate, other and unspecified parts of mouth, parotid gland, other and unspecified major salivary glands, tonsil, esophagus, stomach, small intestine, colon, rectosigmoid junction, | |
| C2% | Malignant neoplasm of: rectum, anus and anal canal, liver and intrahepatic bile ducts, gallbladder, other and unspecified parts of biliary tract, pancreas, other and ill-defined digestive organs | |
| C3% | Malignant neoplasm of: nasal cavity and middle ear, accessory sinuses, larynx, trachea, bronchus and lung, thymus, heart, mediastinum and pleura, other and ill-defined sites in the respiratory system and intrathoracic organs | |
| C4% | Malignant neoplasm of: bone and articular cartilage of limb, bone and articular cartilage of other and unspecified sites, skin, Other and unspecified malignant neoplasm of skin, Merkel cell carcinoma; Mesothelioma, Kaposi's sarcoma, peripheral nerves and autonomic nervous system, retroperitoneum and peritoneum, other connective and soft tissue | |
| C5% | Malignant neoplasm of: breast, vulva, vagina, cervix uteri, corpus uteri, uterus, part unspecified, ovary, other and unspecified female genital organs, placenta | |
| C6% | Malignant neoplasm of: penis, prostate, testis, other and unspecified male genital organs, kidney, except renal pelvis, renal pelvis, ureter, bladder, other and unspecified urinary organs, eye and adnexa | |
| C7% | Malignant neoplasm of: meninges, brain, spinal cord, cranial nerves and other parts of central nervous system, thyroid gland, adrenal gland, other endocrine glands and related structures; Malignant neuroendocrine tumors, Secondary neuroendocrine tumors; other and ill-defined sites, Secondary and unspecified malignant neoplasm of lymph nodes, Secondary malignant neoplasm of respiratory and digestive organs, Secondary malignant neoplasm of other and unspecified sites, | |
| C8% | Malignant neoplasm without specification of site, Hodgkin lymphoma, Follicular lymphoma, Non-follicular lymphoma, Mature T/NK-cell lymphomas, Other specified and unspecified types of non-Hodgkin lymphoma, Other specified types of T/NK-cell lymphoma, Malignant immunoproliferative diseases and certain other B-cell lymphomas | |
| C90% | Multiple myeloma and malignant plasma cell neoplasms | |
| C91% | Lymphoid leukemia | |
| C93% | Monocytic leukemia | |
| C94% | Other leukemias of specified cell type | |
| C95% | Leukemia of unspecified cell type | |
| C96% | Other and unspecified malignant neoplasms of lymphoid, hematopoietic and related tissue | |
| E66% | Overweight and obesity | Obesity |

| | | |
|---|---|---|
| *E03%* | Other hypothyroidism | Hypothyroidism |
| *I10* | Essential (primary) hypertension | Hypertension |
| *I11%* | Hypertensive heart disease | |
| *I13%* | Hypertensive heart and chronic kidney disease | |
| *E78.5* | Hyperlipidemia, unspecified | Hyperlipidemia |
| *E78.4%* | Other hyperlipidemia | |
| *K21%* | Gastro-esophageal reflux disease | Gerd |
| *D68%* | Other coagulation defects | Coagulopathy |

**Supplemental Table 3. Definitions of AML mutation variables.**

| | *Dataset* | *NGS* | *FISH* | *PCR* | *CYT* | *Unknown test* |
|---|---|---|---|---|---|---|
| *IDH1* | CU | Y | | Y | | |
| | RWC | Y | | Y | | Y |
| *IDH2* | CU | Y | | Y | | |
| | RWC | Y | | Y | | Y |
| *NPM1* | CU | Y | | Y | | |
| | RWC | Y | | Y | | Y |
| *TP53* | CU | Y | Y | | | |
| | RWC | Y | | Y | | Y |
| *FLT3-ITD* | CU | Y | | Y | | |
| | RWC | Y | | Y | | Y |

#### SUPPLEMENTAL FIGURE LEGENDS

**Supplemental Figure 1. Impact of allo-HCT and propensity score (PS) adjusted Kaplan-Meier (KM) for CU and RWC.** A) Comparison of OS for CU vs RWC cohorts with allo-HCT included (right) and excluded (left). Note the CU cohort has ~29% allo-HCT rate and the RWC has ~6% allo-HCT rate; B) PS adjusted standard mean differences (SMDs) for comorbidity; C**)** Left-bottom figure corresponds to unadjusted KM and right-bottom to PS-adjusted KM for the CU and RWC (excluding allo-HCT).

**Supplemental Figure 2. Kaplan-Meier (KM) analyses for overall survival with respect to single biomarkers from the CU dataset.** allo-HCT recipients included (left panels) and excluded (right panels).

**Supplemental Figure 3. Kaplan-Meier (KM) analyses for overall survival with respect to single biomarkers from the RWC dataset.** allo-HCT recipients included (left panels) and excluded (right panels).

**Supplemental Figure 4. Kaplan-Meier (KM) analyses for overall survival with respect to two-factor interactions between biomarkers from the CU dataset.** allo-HCT recipients included (left panels) and excluded (right panels). For all two-factor KMs, "yes/no" refers to the order of features in the Feature figure legend.

**Supplemental Figure 5. Kaplan-Meier (KM) analyses for overall survival with respect to two-factor interactions between biomarkers from the RWC dataset.** allo-HCT recipients included (left panels) and excluded (right panels). For all two-factor KMs, "yes/no" refers to the order of features in the Feature figure legend.

**Supplemental Figure 6. Application of the RRM in defined CU cohorts.**

**Supplemental Figure 7. Application of the mPRS and e-mPRS in the defined CU cohorts.**

**Supplemental Figure 8: Application of the RRM and ELN22 comparisons in the RWC.**

**Supplemental Figure 9. Comparison of the RRM, mPRS, and e-mPRS in the RWC.**

**Supplemental Figure 10. Evaluation of predictive performance of RRM, mPRS, e-mPRS, and ELN22 risk models for overall survival for up to 4 years in the RWC (allo-HCT censored) based on penalized Cox-PH model.** Models were evaluated on four scenarios. A) FAS with mPRS; B) IAS with mPRS; C) FAS with e-mPRS; D) IAS with e-mPRS. Results are averaged over 10 imputations.

**Supplemental Figure 1. A) Kaplan-Meier (KM) analyses for overall survival (OS) in the CU patient cohort compared to the RWC.** OS with allo-HCT patients included (left panel) and with allo-HCT patients excluded (right panel).

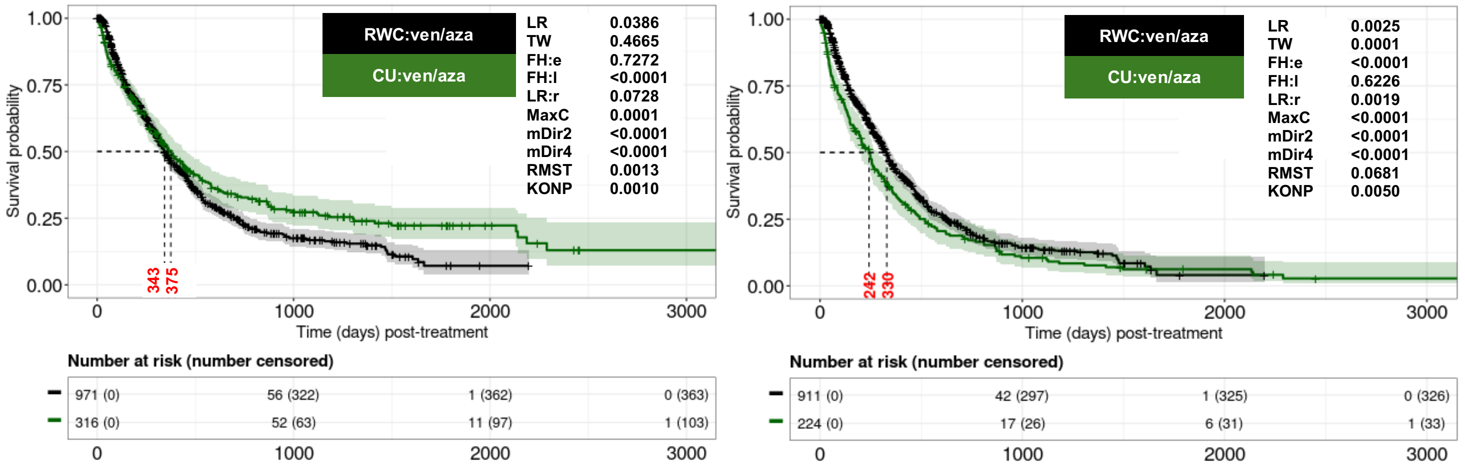


**B. Standardized mean differences (SMD) for co-morbidities in the CU and RWC.**

| Co-morbidities | SMD | w-SMD |
|---|---|---|
| Obesity | 0.90 | 0.02 |
| Prior non-AML cancer | 0.56 | 0.01 |
| Prior heart disease | 0.60 | 0.01 |
| Prior MDS | 0.43 | 0.12 |
| Prior CKD | 0.45 | 0.04 |
| Prior coagulopathy | 0.36 | 0.03 |
| Prior VTE | 0.37 | 0.09 |
| Prior COPD | 0.21 | 0.10 |
| Prior GERD | 0.52 | 0.03 |
| Prior hyperlipidemia | 0.46 | 0.01 |
| Prior hypertension | 0.46 | 0.10 |
| Prior hypothyroidism | 0.43 | 0.05 |

*Remarks:*
Propensity scores (PS) were estimated via generalized linear model with logit link adjusting for comorbidities as reported in the above table. Standardized mean difference (SMD) values were computed after adjusting for propensity scores which are termed as weighted SMD (w-SMD).

**Supplemental Figure 1, cont'd**

**C. KM of original samples (left) and KM based on PS weighted samples (right).**

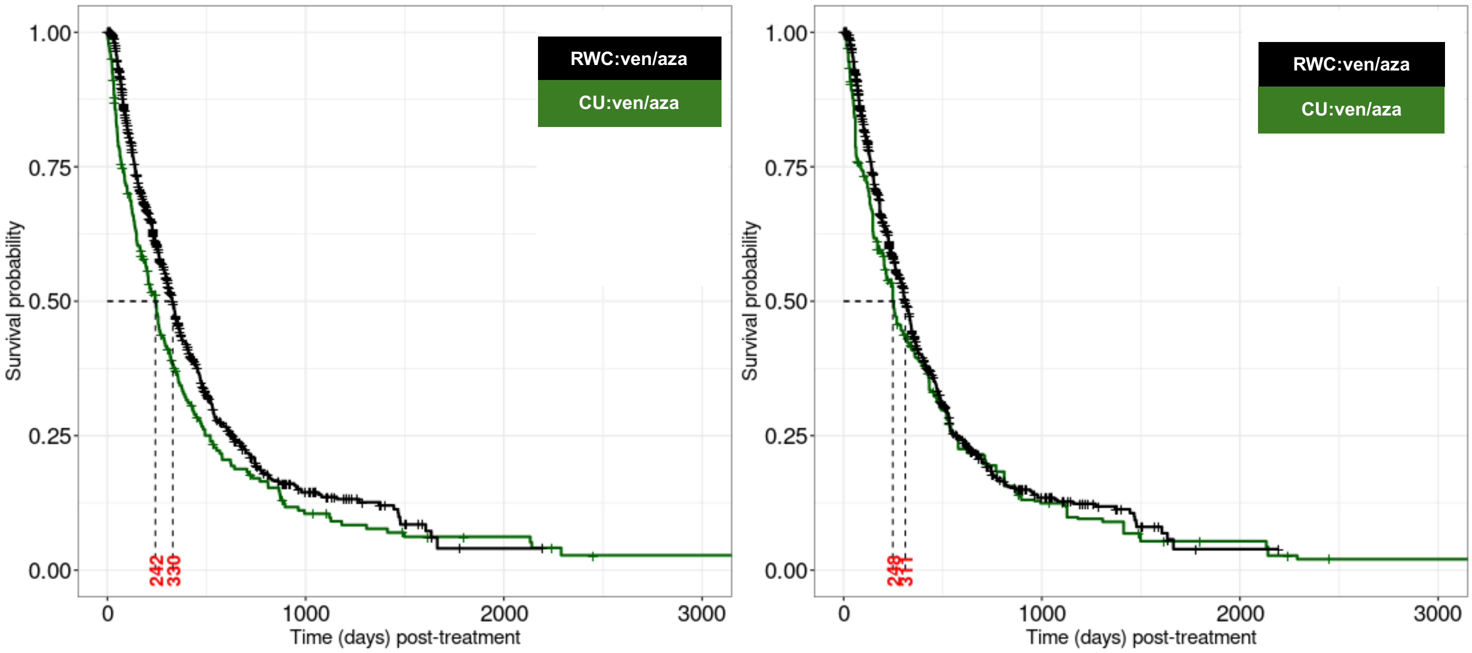

**Supplemental Figure 2. Kaplan-Meier analyses of features used in the Refined Risk Model (RRM) in the CU cohort.** Left panels: Including allo-HCT and Right panels: Excluding allo-HCT.

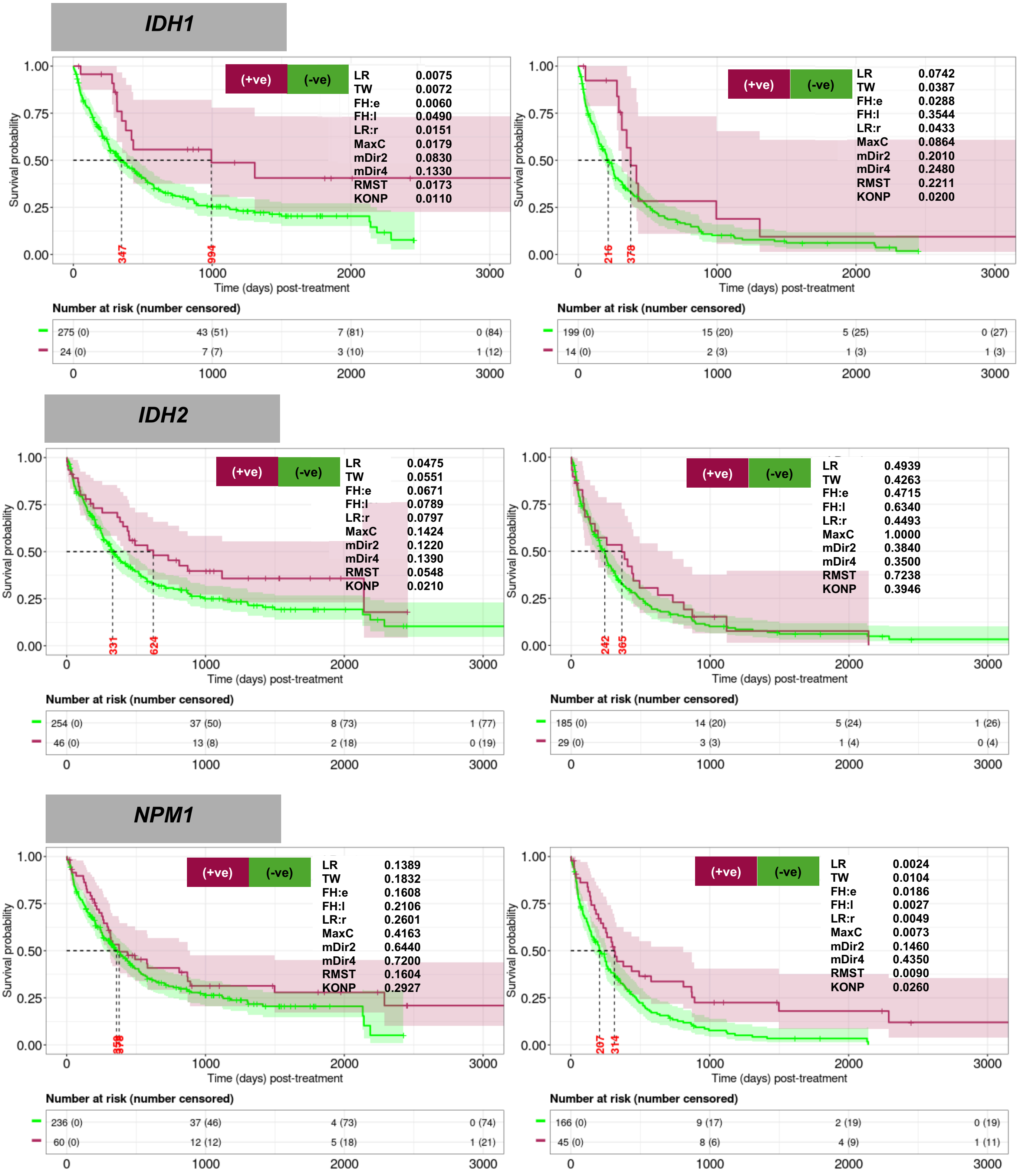

#### Supplemental Figure 2, cont'd

##### *Cytogenetics: good risk*

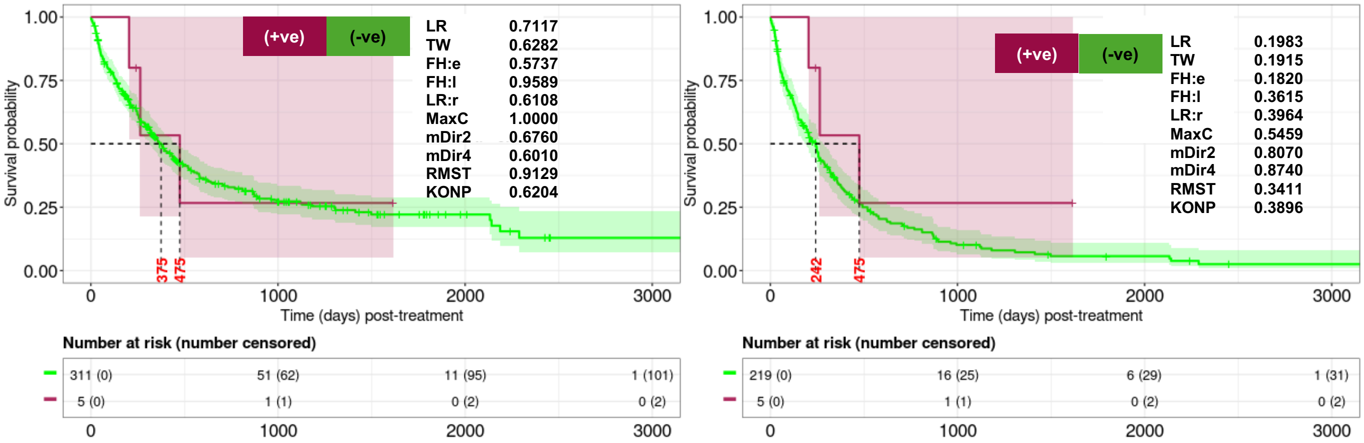


##### *Del7q*

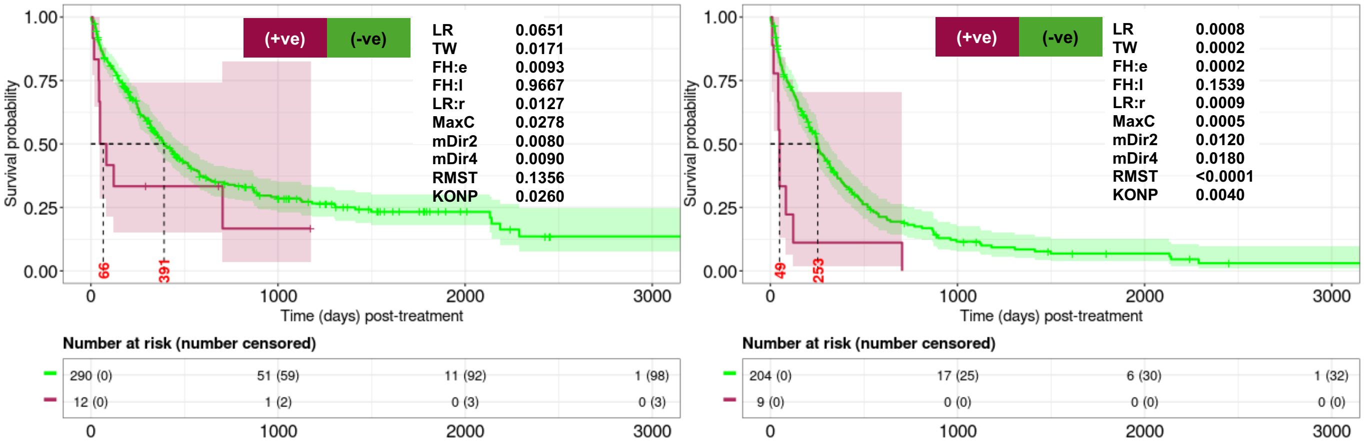


##### *Minus5*

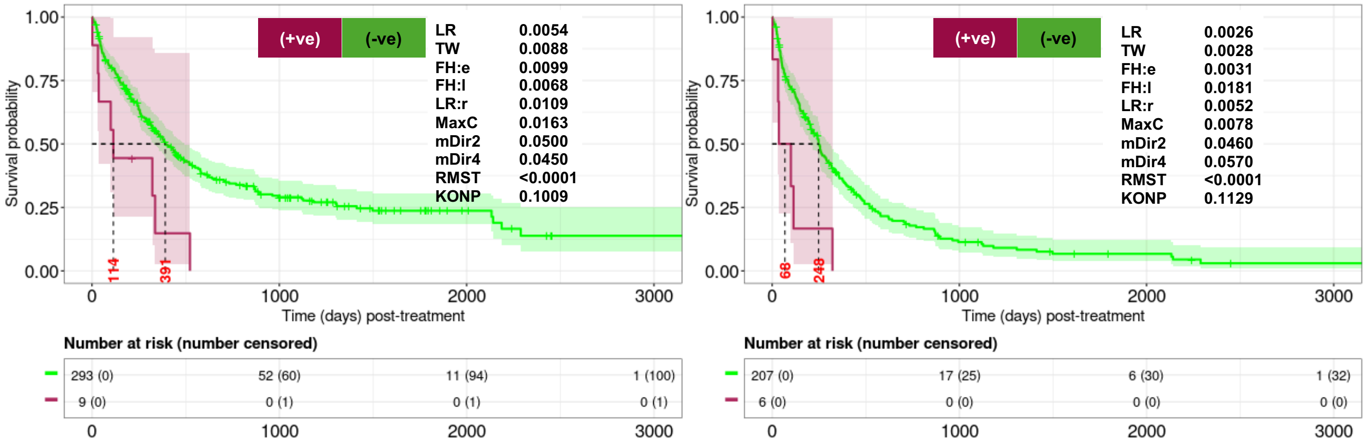

### Supplemental Figure 2, cont'd

#### *Minus17*

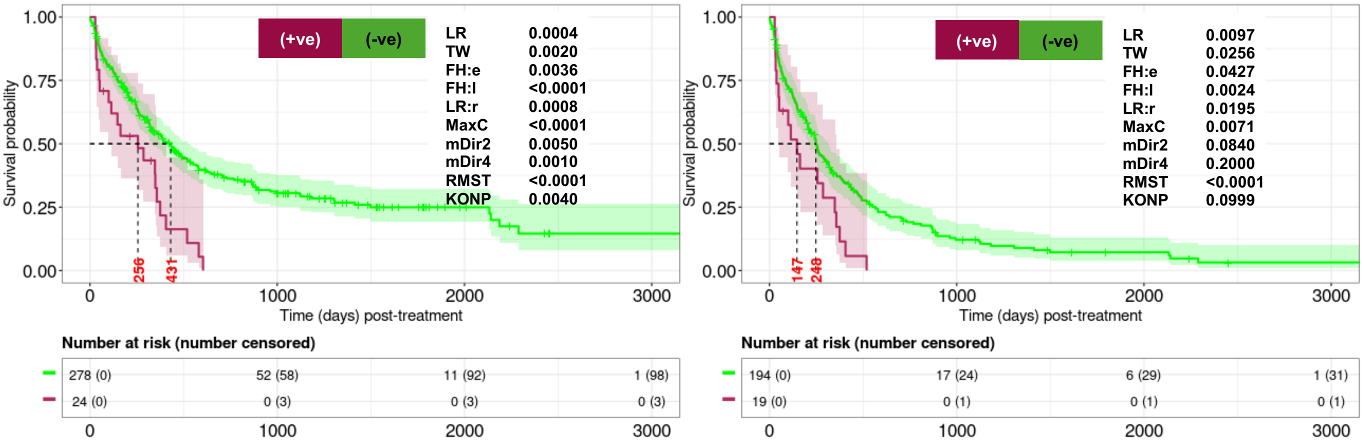


#### *Inv3*

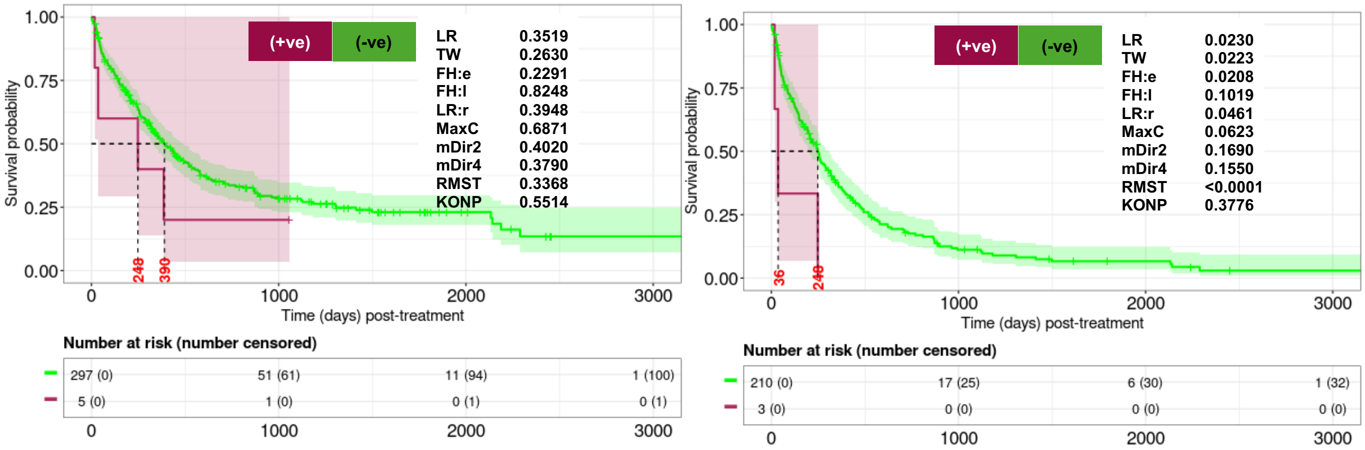


#### *Complex*

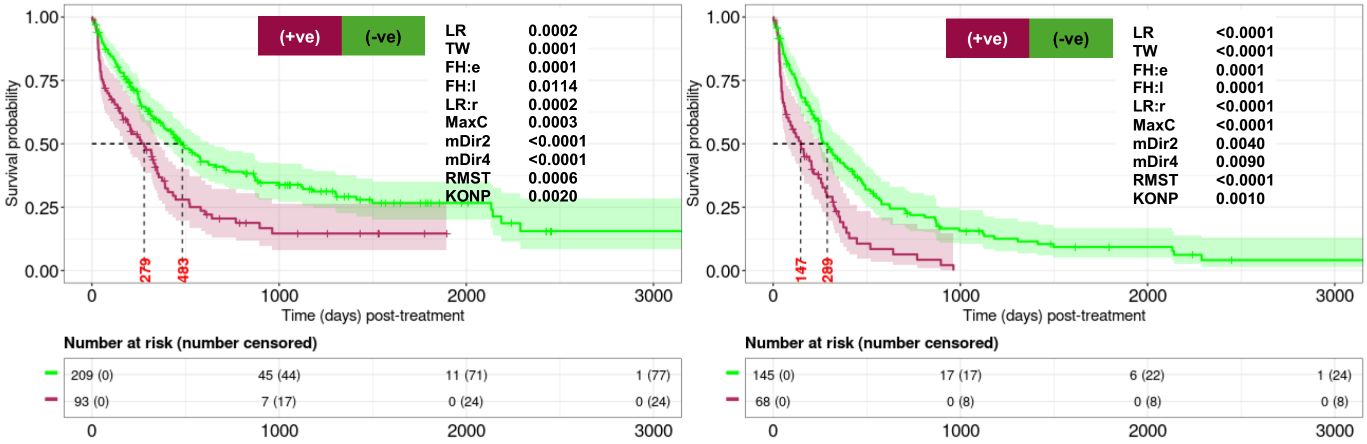

#### Supplemental Figure 2, cont'd

### *T9;11*

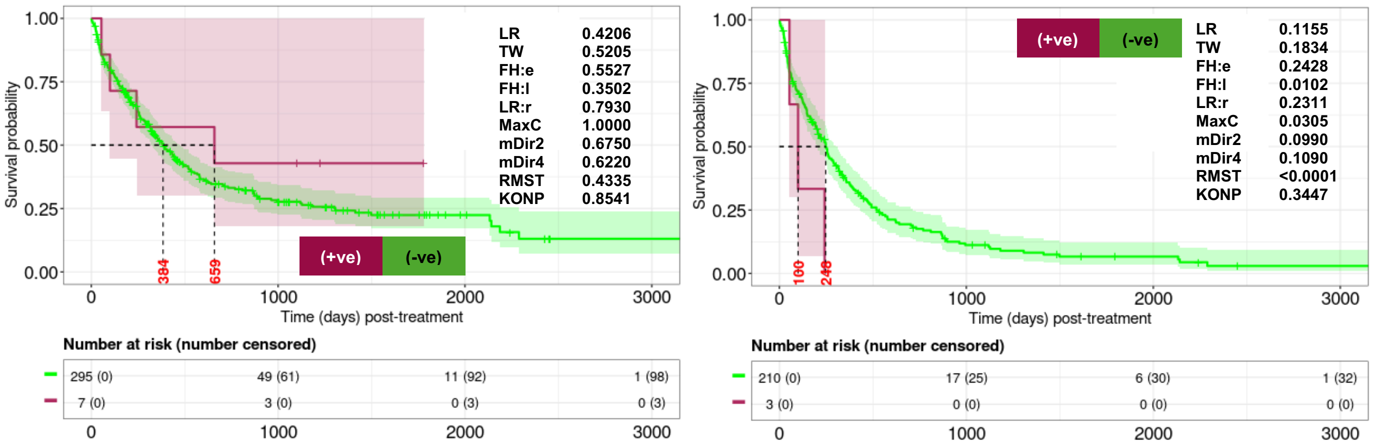


### *TP53*

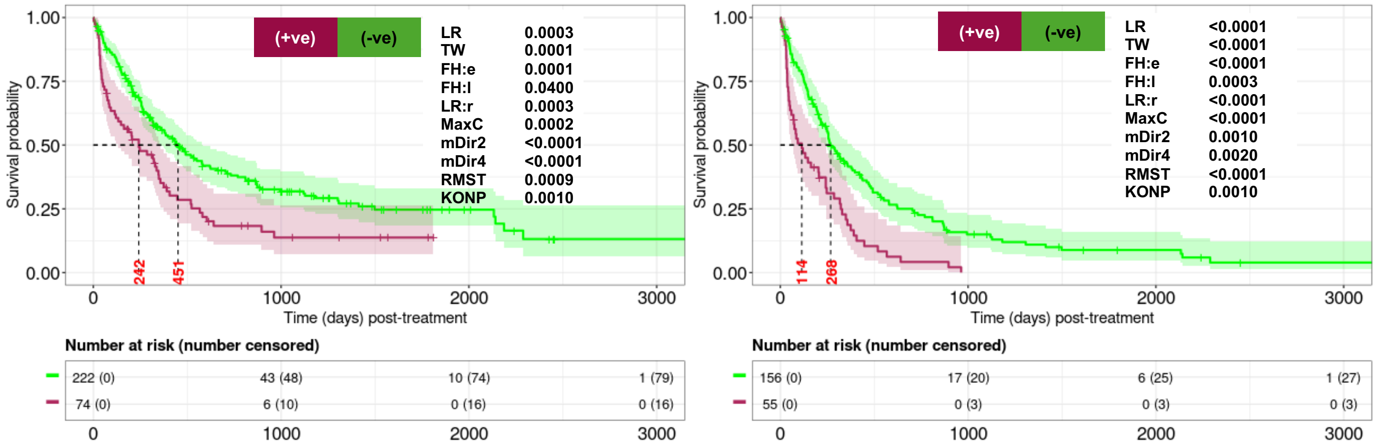


##### *FLT3-ITD*

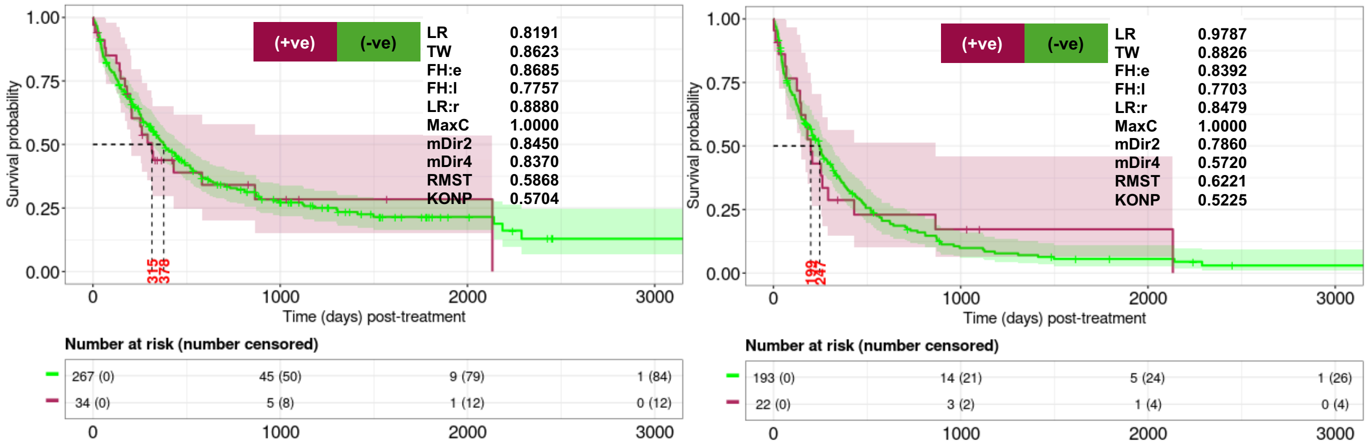

#### Supplemental Figure 2, cont'd

##### *NRAS*

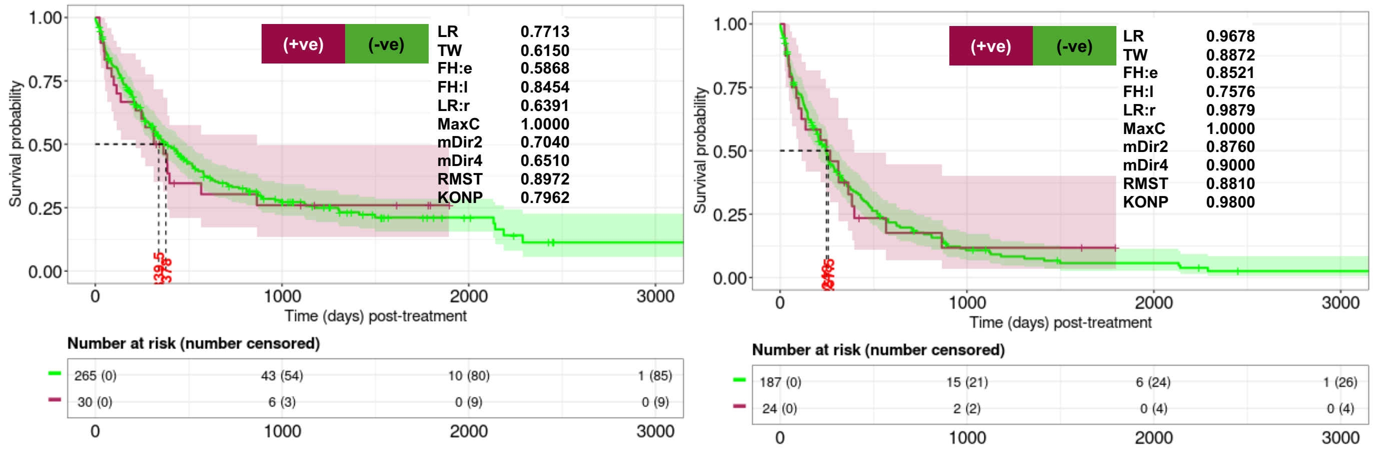


##### *KRAS*

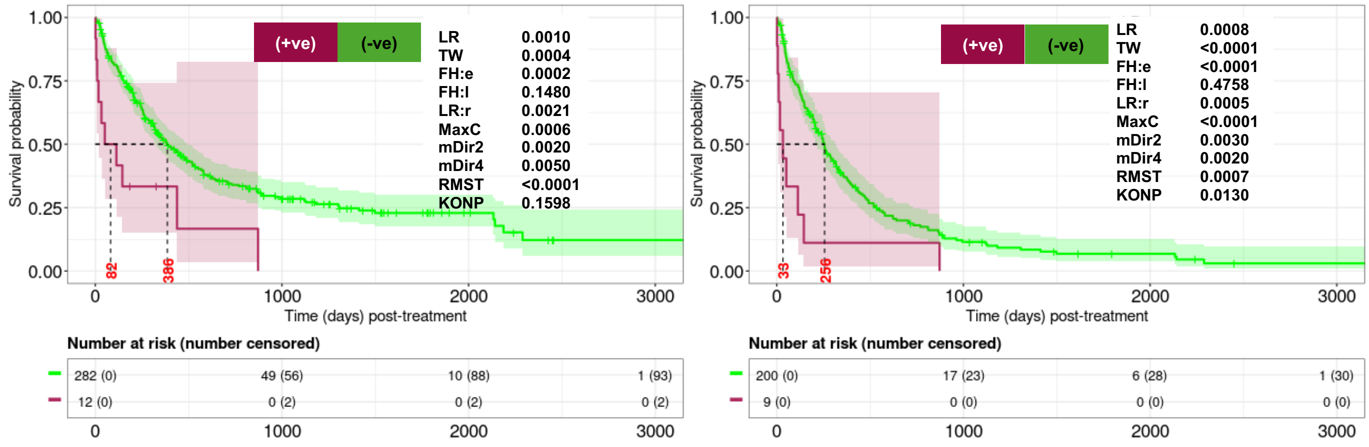

**Supplemental Figure 3. Kaplan-Meier analyses for single biomarkers from the RWC.** OS including allo-HCT patients (left panel) and excluding allo-HCT patients (right panel).

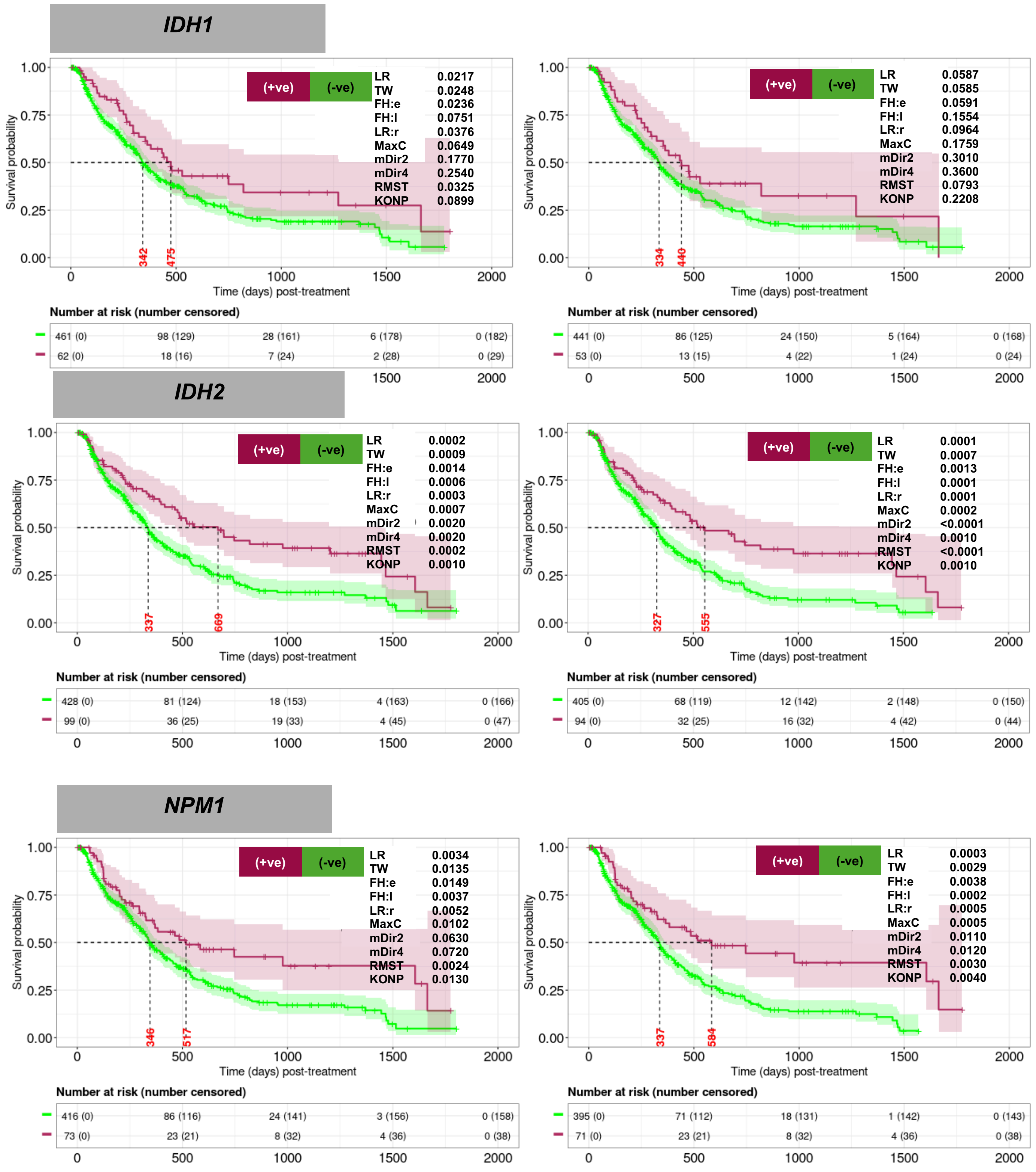

#### Supplemental Figure 3, cont'd

##### *Cytogenetics good*

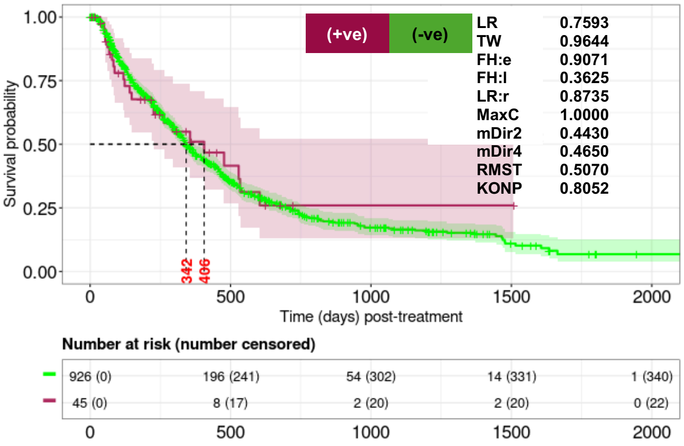


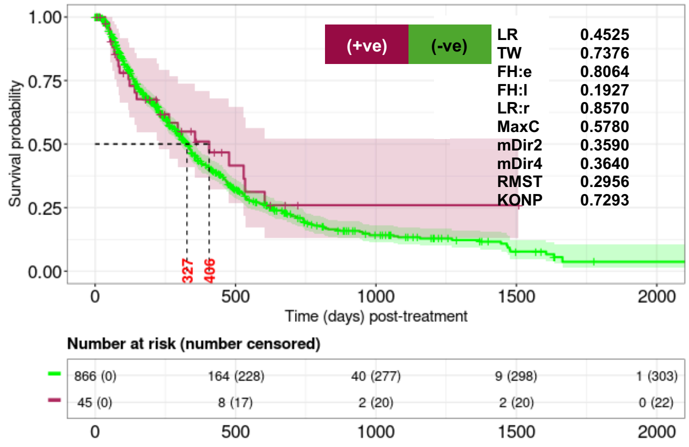


##### *Minus7*

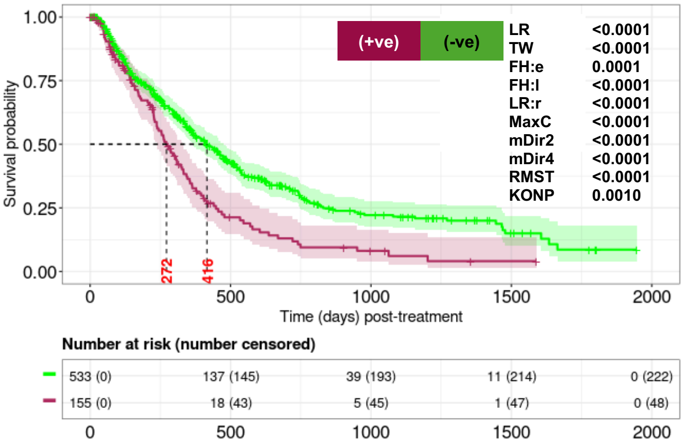


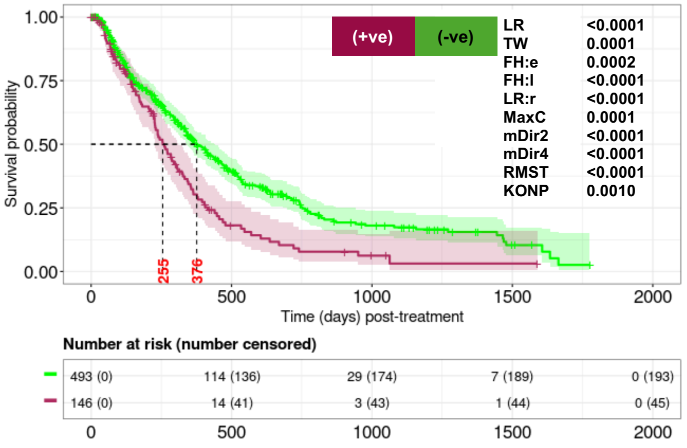


##### *Minus5*

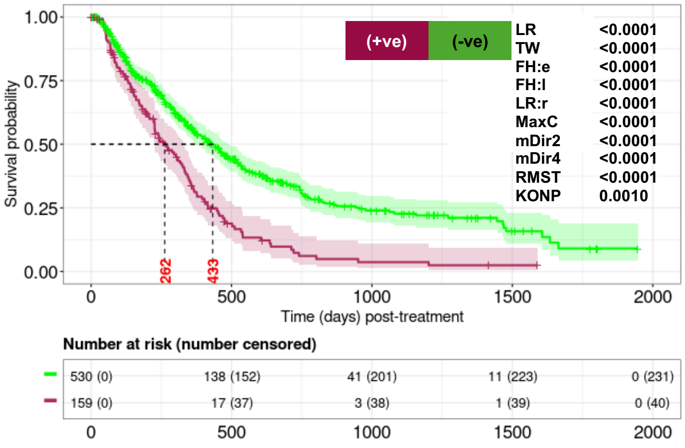


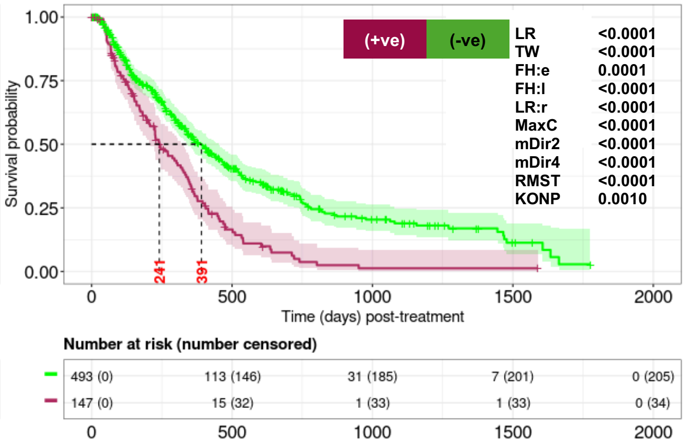

#### Supplemental Figure 3, cont'd

##### *Minus17*

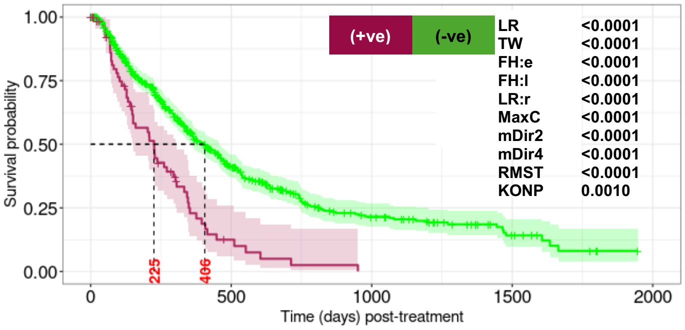


**Number at risk (number censored)**

| | 0 | 500 | 1000 | 1500 | 2000 |
|---|---|---|---|---|---|
| green | 619 (0) | 150 (176) | 44 (226) | 12 (249) | 0 (258) |
| red | 69 (0) | 5 (14) | 0 (14) | 0 (14) | 0 (14) |

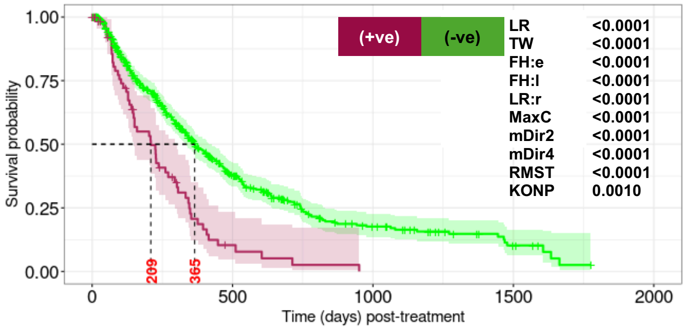


**Number at risk (number censored)**

| | 0 | 500 | 1000 | 1500 | 2000 |
|---|---|---|---|---|---|
| green | 572 (0) | 124 (166) | 32 (206) | 8 (222) | 0 (227) |
| red | 67 (0) | 4 (13) | 0 (13) | 0 (13) | 0 (13) |

##### *Inv3*

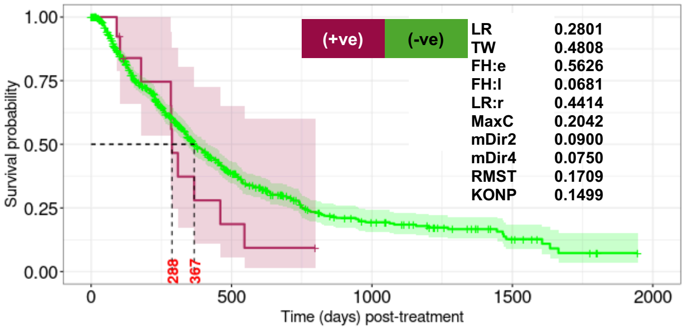


**Number at risk (number censored)**

| | 0 | 500 | 1000 | 1500 | 2000 |
|---|---|---|---|---|---|
| green | 675 (0) | 153 (187) | 44 (236) | 12 (259) | 0 (268) |
| red | 13 (0) | 2 (2) | 0 (3) | 0 (3) | 0 (3) |

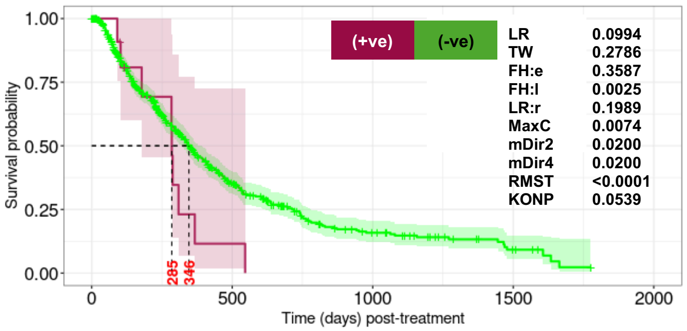


**Number at risk (number censored)**

| | 0 | 500 | 1000 | 1500 | 2000 |
|---|---|---|---|---|---|
| green | 628 (0) | 127 (176) | 32 (216) | 8 (232) | 0 (237) |
| red | 11 (0) | 1 (2) | 0 (2) | 0 (2) | 0 (2) |

##### *Complex*

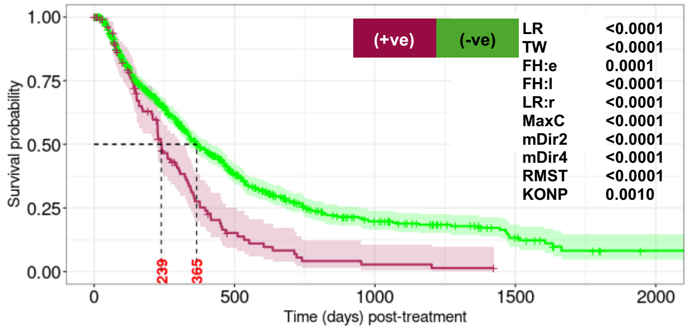


**Number at risk (number censored)**

| | 0 | 500 | 1000 | 1500 | 2000 |
|---|---|---|---|---|---|
| green | 856 (0) | 193 (233) | 54 (297) | 16 (325) | 1 (336) |
| red | 115 (0) | 11 (25) | 2 (25) | 0 (26) | 0 (26) |

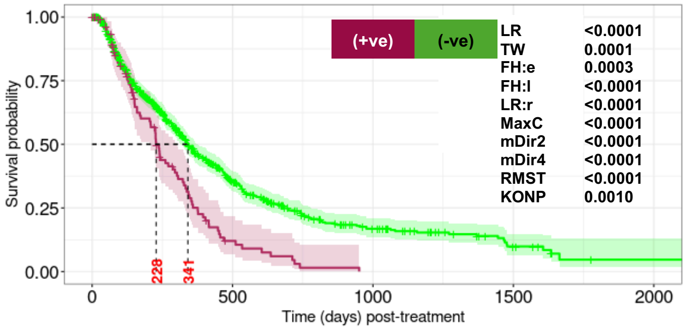


**Number at risk (number censored)**

| | 0 | 500 | 1000 | 1500 | 2000 |
|---|---|---|---|---|---|
| green | 803 (0) | 164 (222) | 42 (274) | 11 (295) | 1 (302) |
| red | 108 (0) | 8 (23) | 0 (23) | 0 (23) | 0 (23) |

#### Supplemental Figure 3, cont'd

### *T9;11*

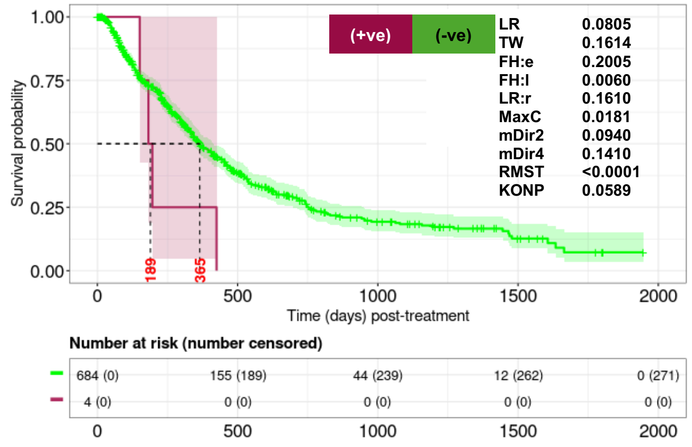


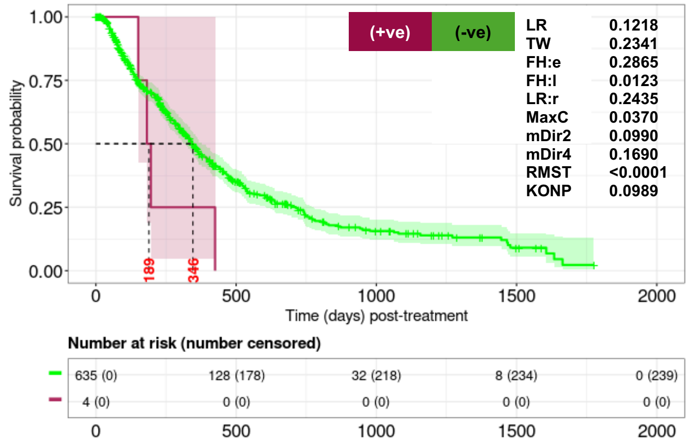


### *TP53*

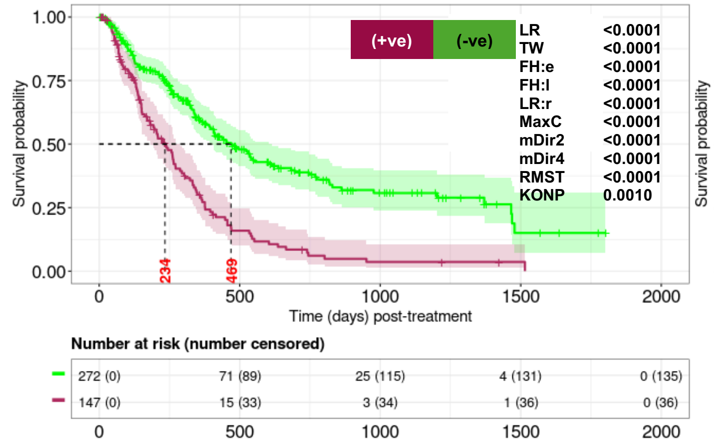


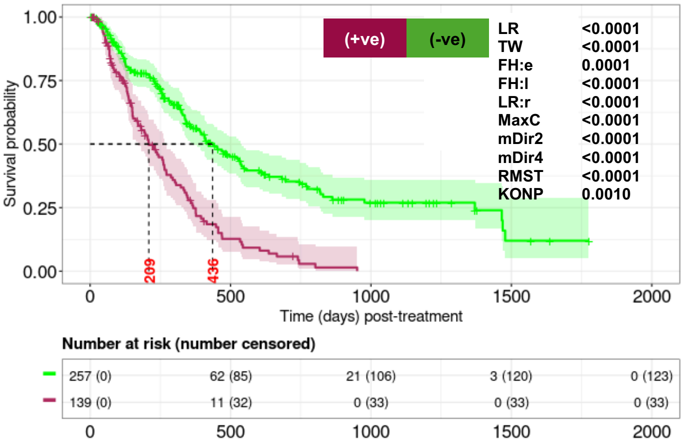


##### *FLT3-ITD*

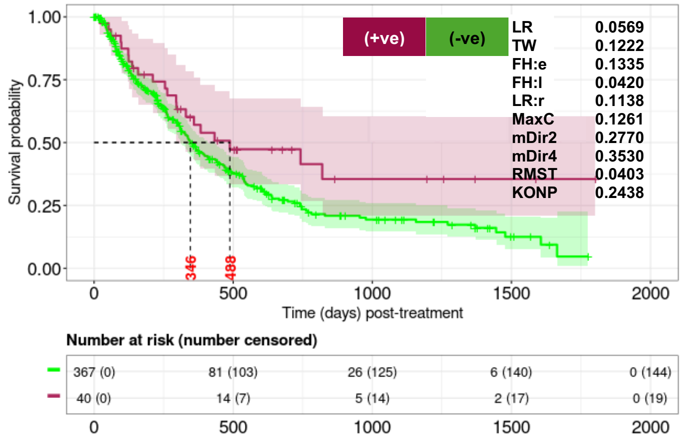


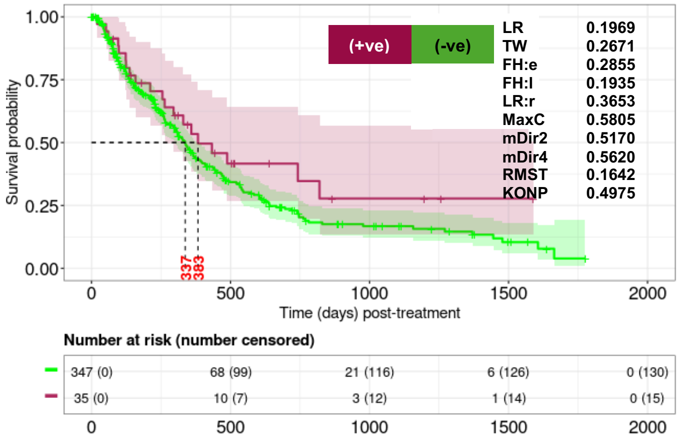

#### Supplemental Figure 3, cont'd

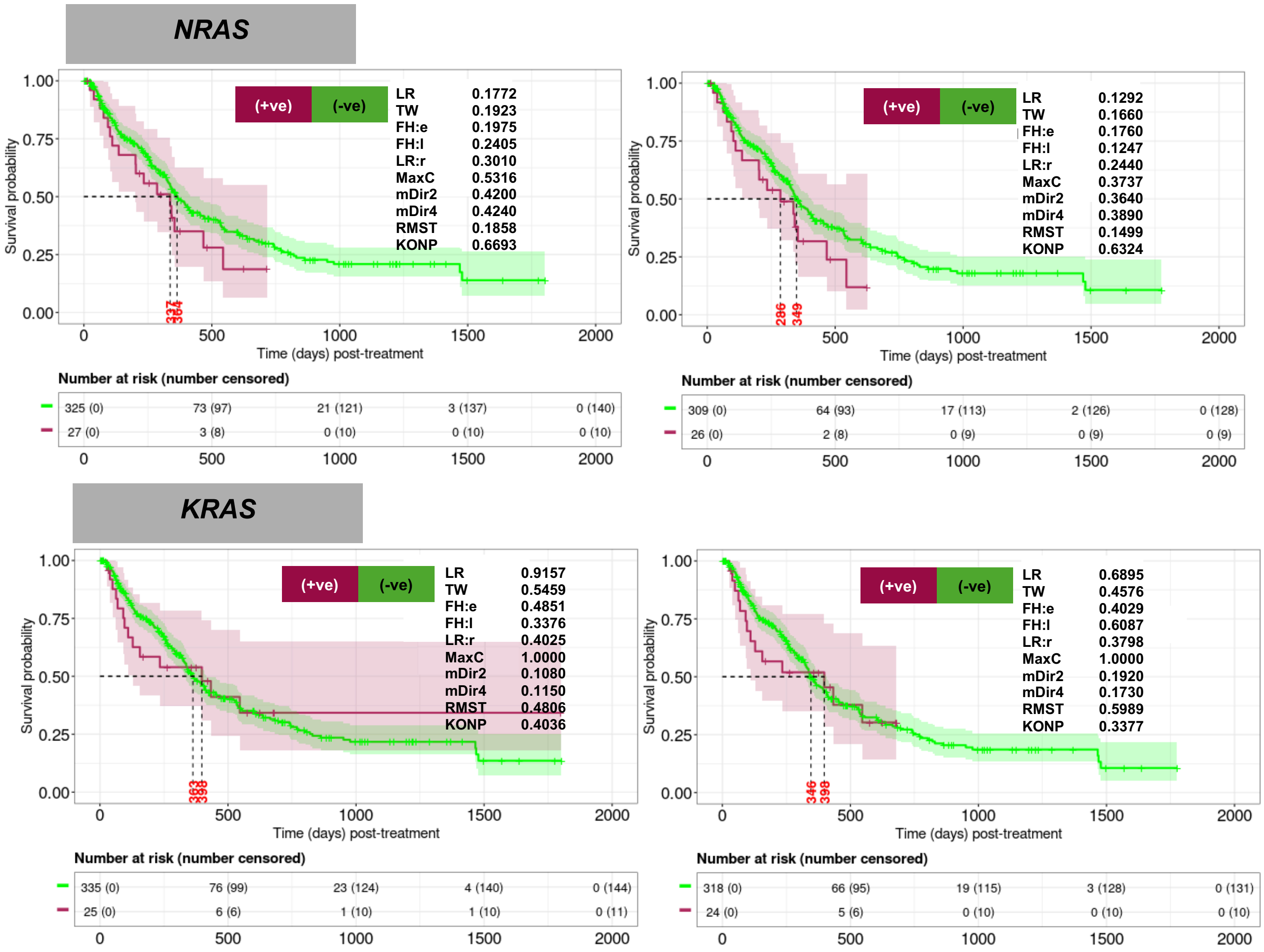

**Supplemental Figure 4. Kaplan-Meier analyses for two-factor interactions between biomarkers from the CU cohort.** OS including allo-HCT patients (left panel) and excluding allo-HCT patients (right panel).

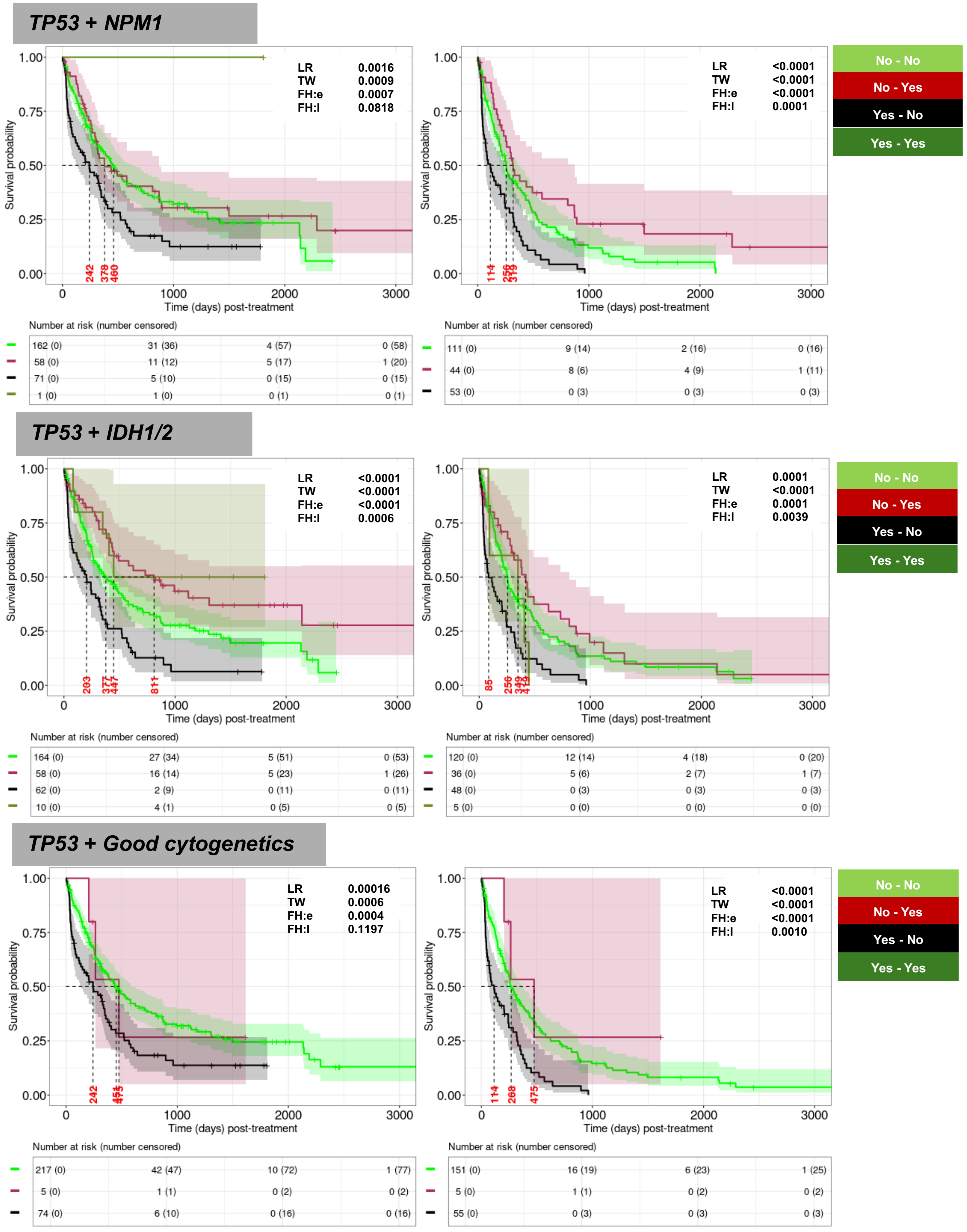

#### Supplemental Figure 4, cont'd

##### *TP53 + IDH1/IDH2/NPM1/Good cytogenetics*

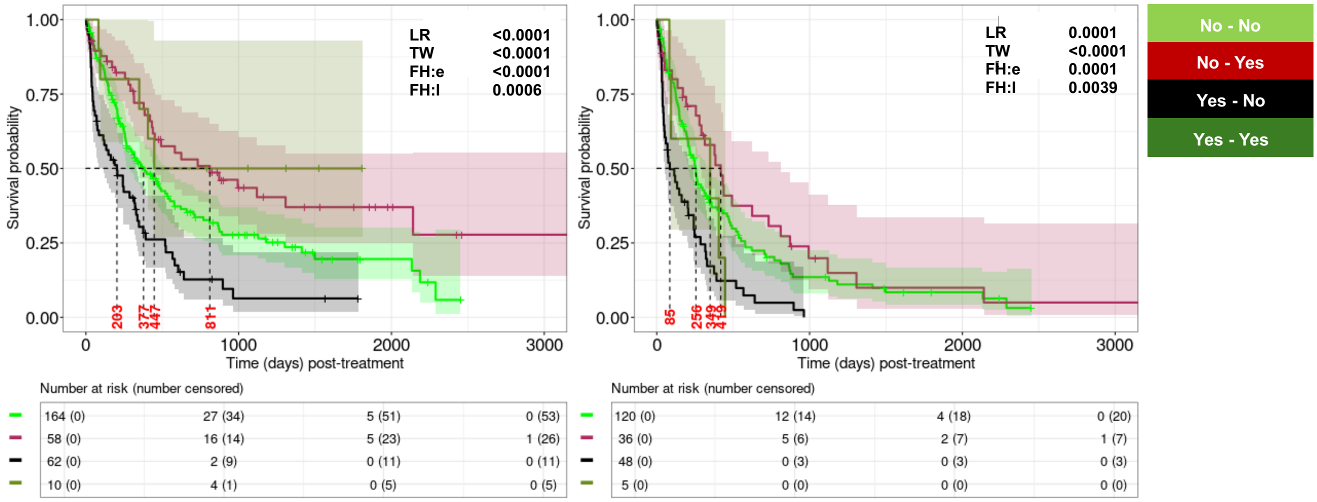


##### *NPM1 + FLT3-ITD*

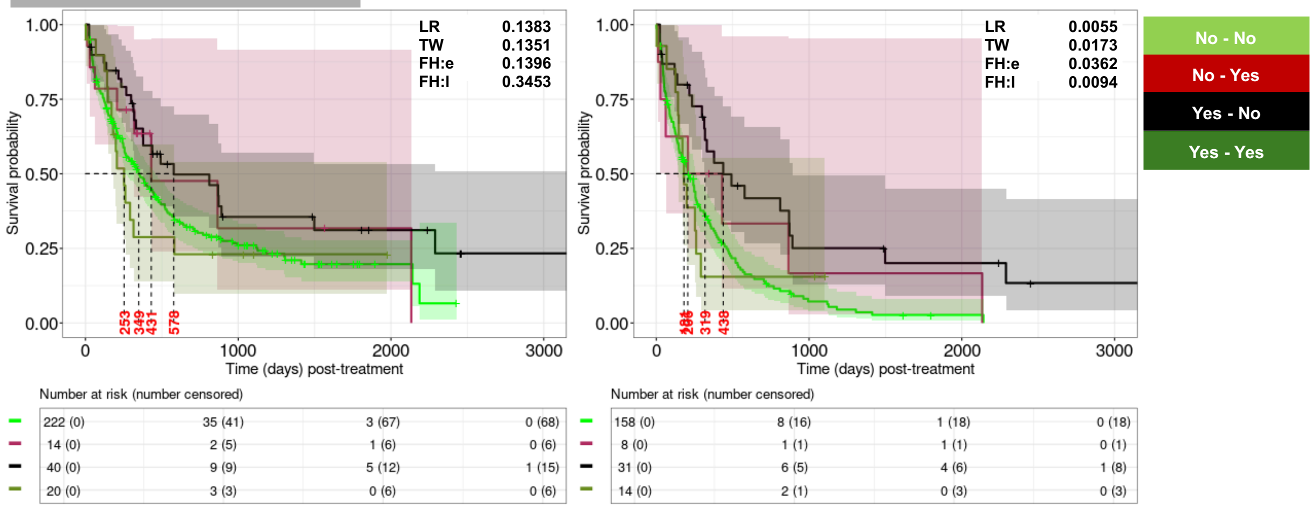


##### *FLT3-ITD + IDH1/2*

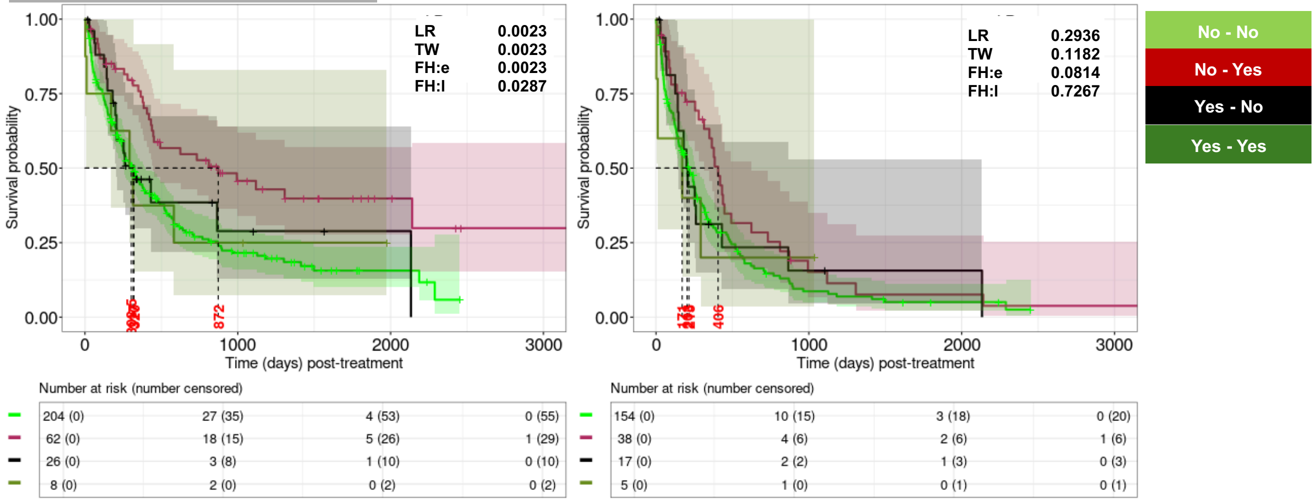

#### Supplemental Figure 4, cont'd

##### *FLT3-ITD + Good cytogenetics*

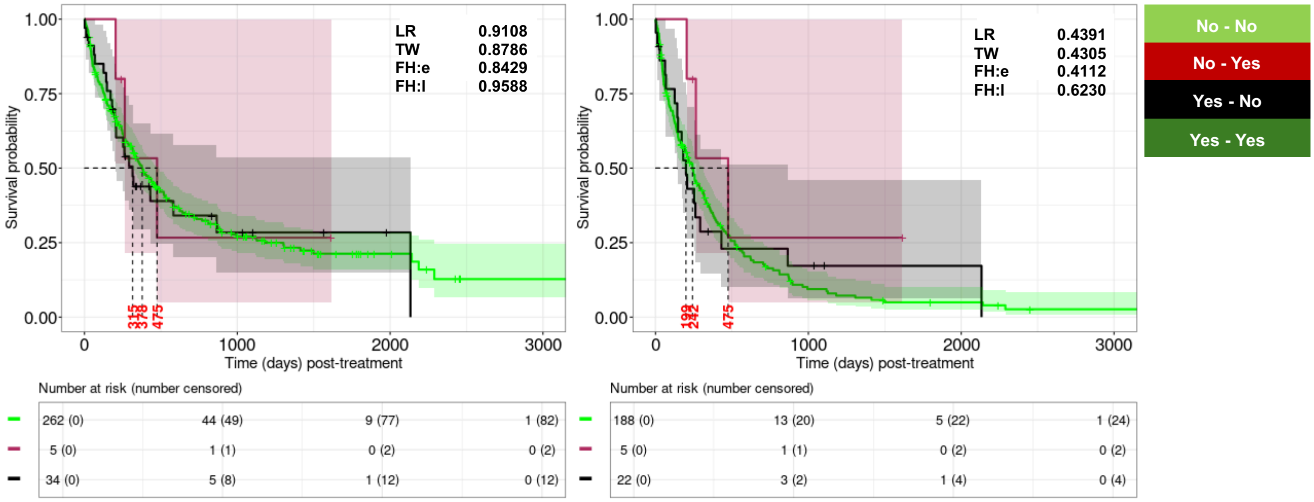


##### *FLT3-ITD + IDH1/IDH2/NPM1/Good cytogenetics*

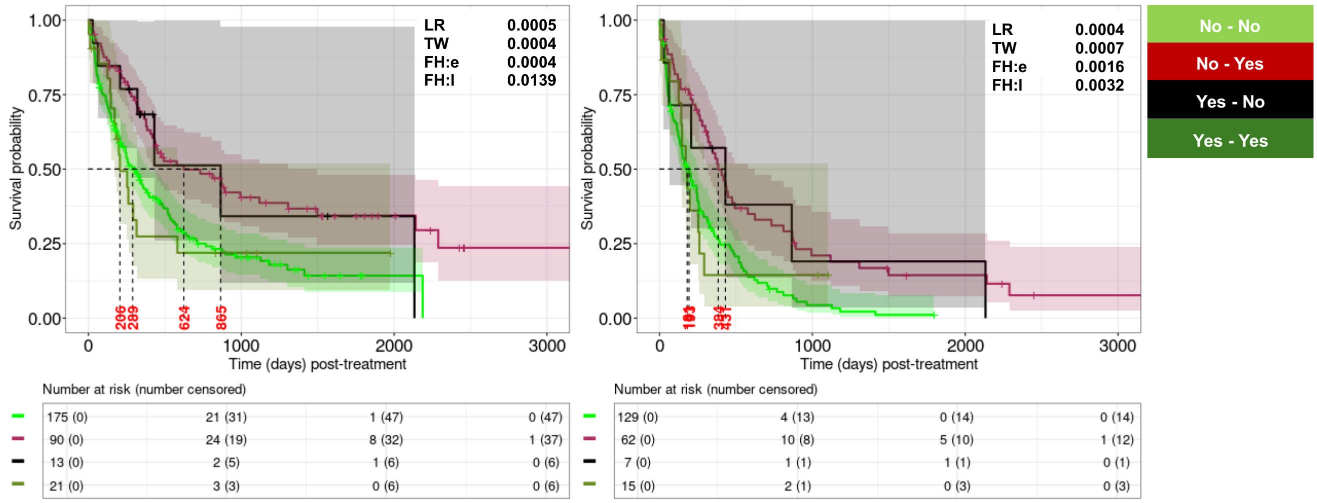


##### *IDH1/IDH2/NPM1/Good cytogenetics + Complex cytogenetics*

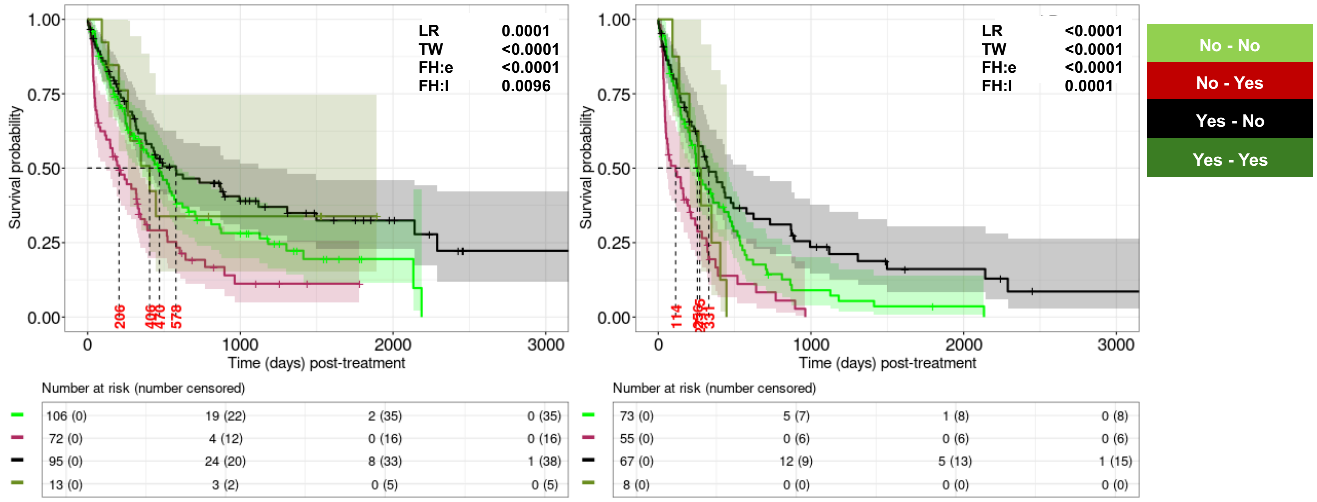

#### Supplemental Figure 4, cont'd

##### *TP53 + complex cytogenetics*

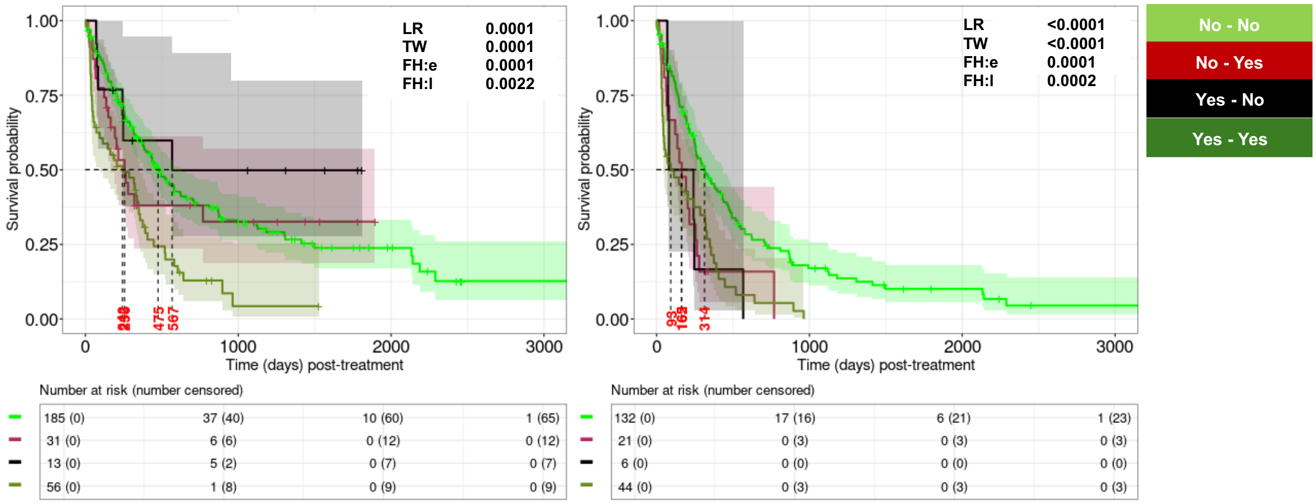


##### *TP53 + Minus17*

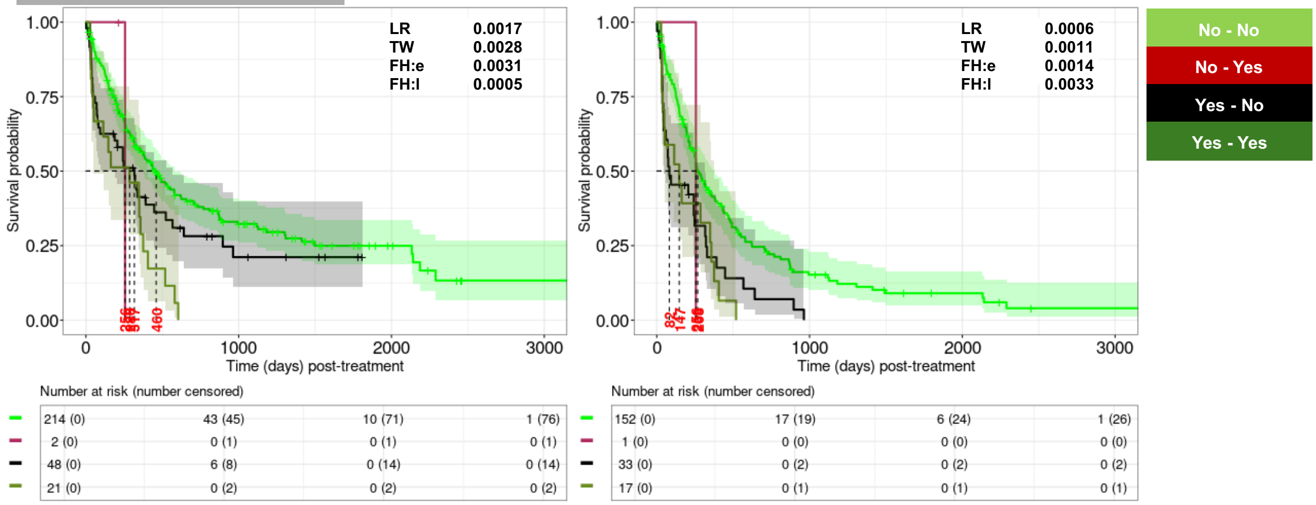


##### *TP53 + Minus5*

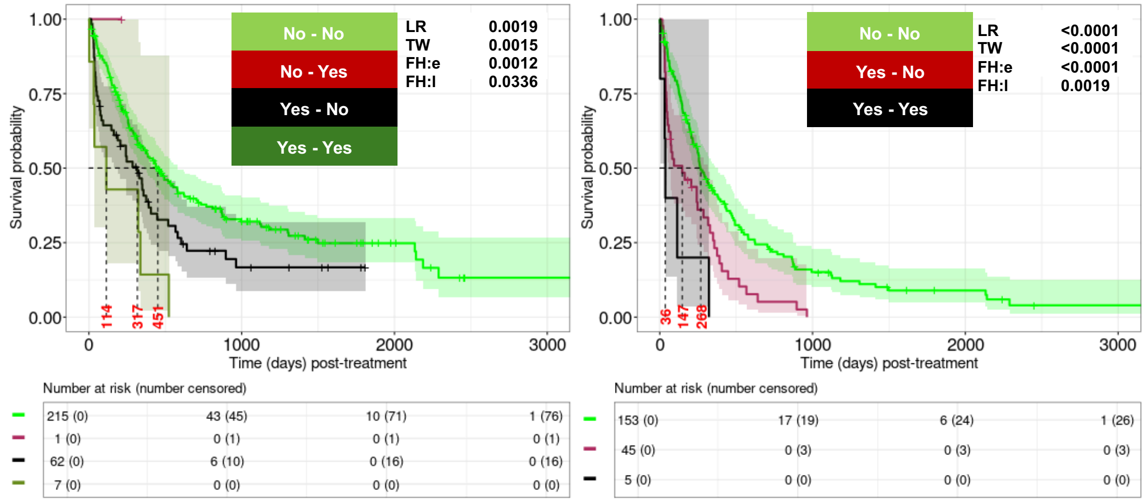

#### Supplemental Figure 4, cont'd

##### *TP53 + Del7q*

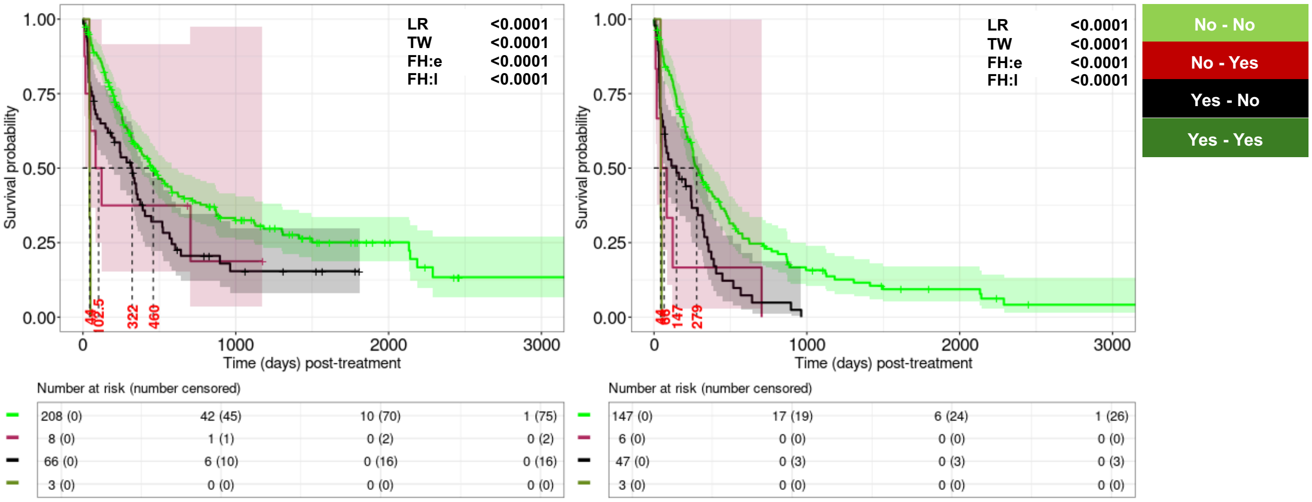


##### *TP53 + Inv3*

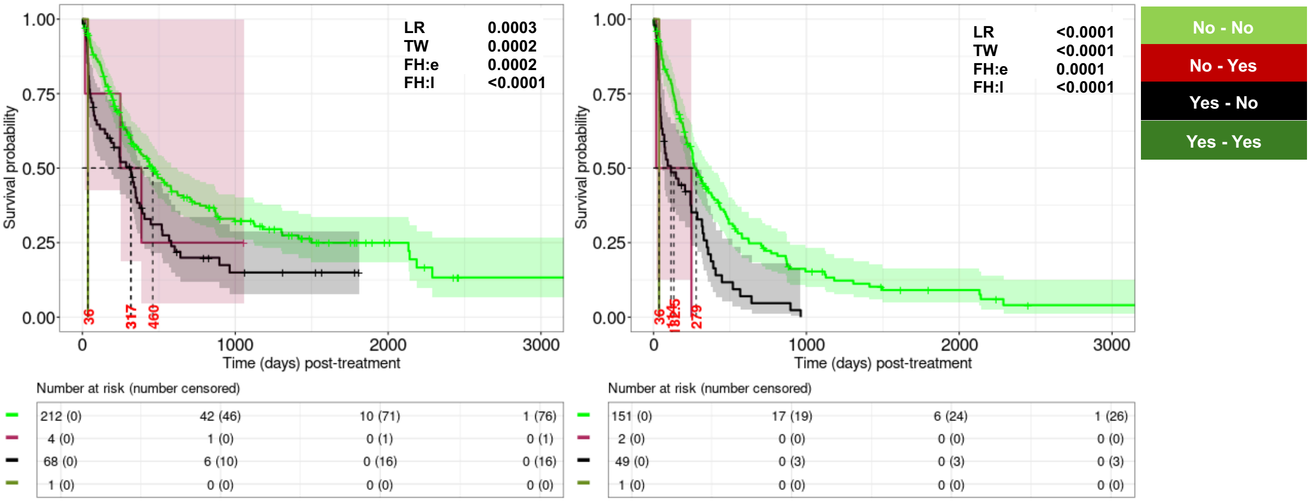


##### *FLT3-ITD + TP53*

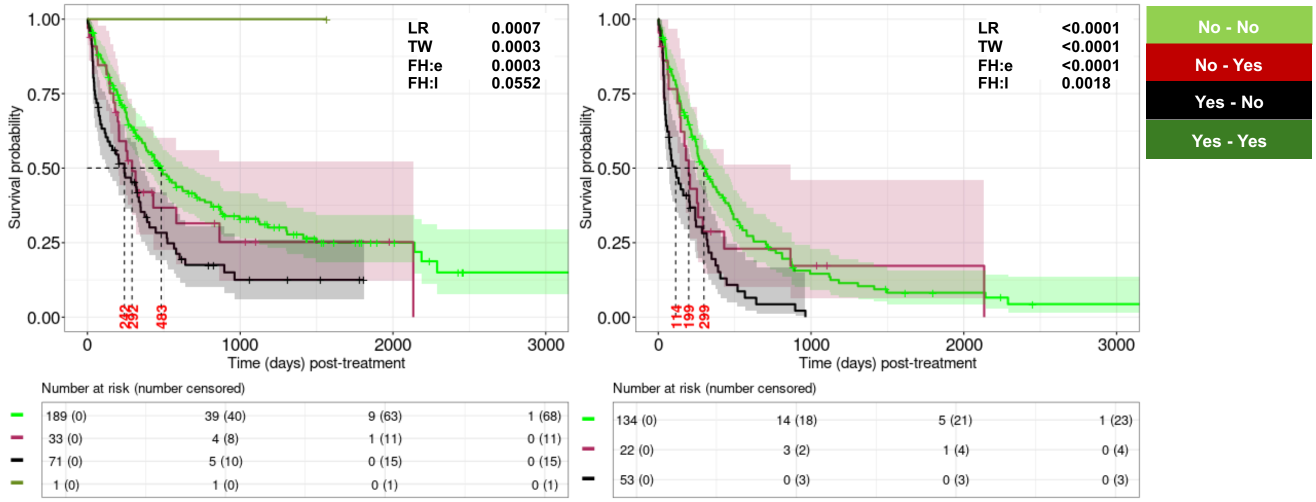

### Supplemental Figure 4, cont'd

#### *FLT3-ITD + KRAS*

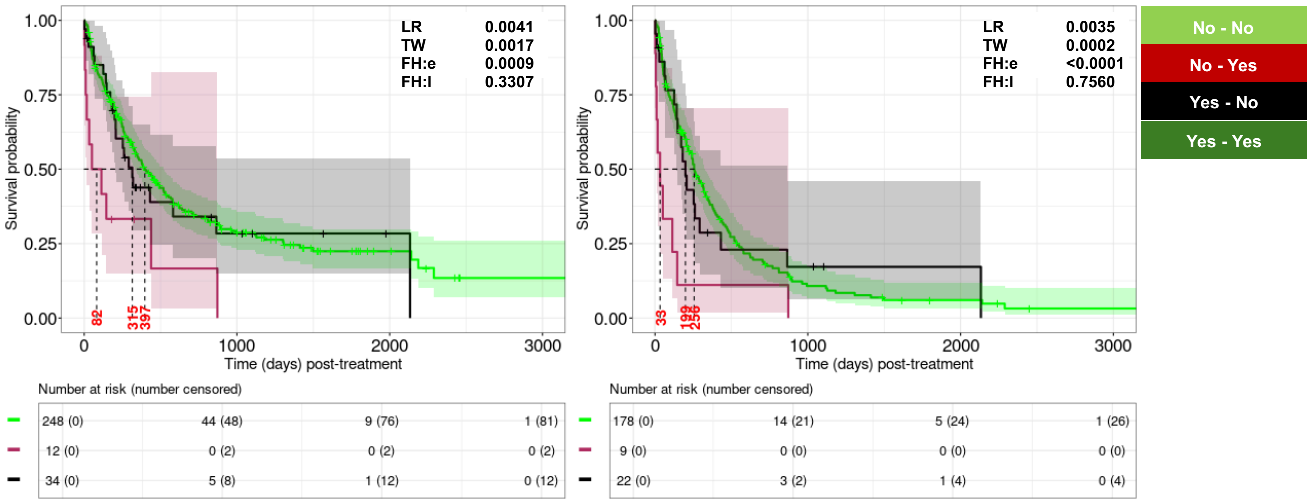


#### *NRAS + FLT3-ITD*

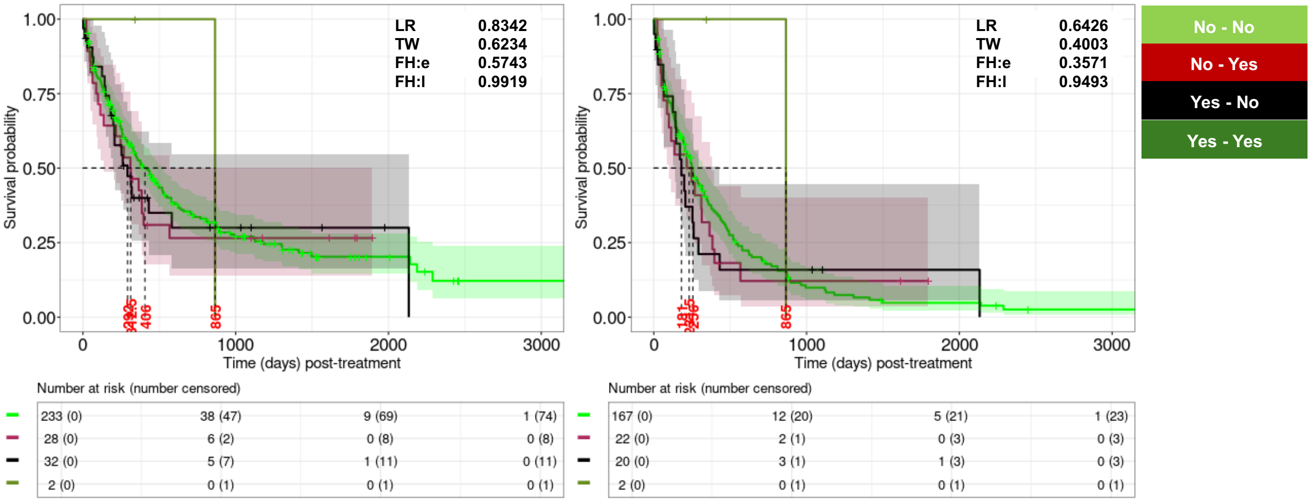


## *TP53 + T9;11*

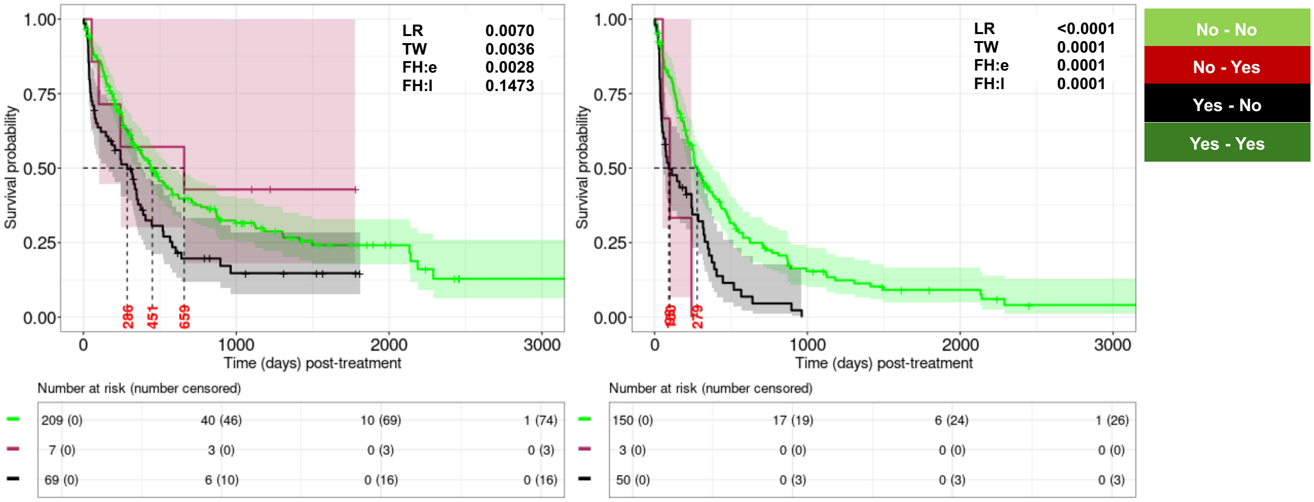

#### Supplemental Figure 4, cont'd

##### *IDH1/IDH2/NPM1/Good cytogenetics + T9;11*

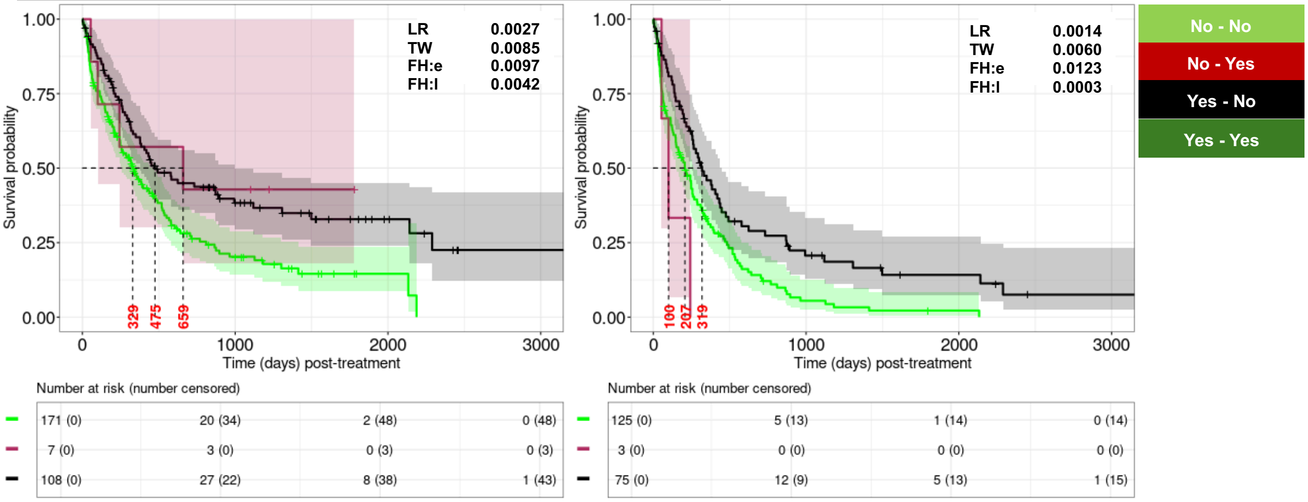

**Supplemental Figure 5. Kaplan-Meier analyses for two-factor interactions between biomarkers from the RWC.** OS including allo-HCT patients (left panel) and excluding allo-HCT patients (right panel).

#### *NPM1 + TP53*

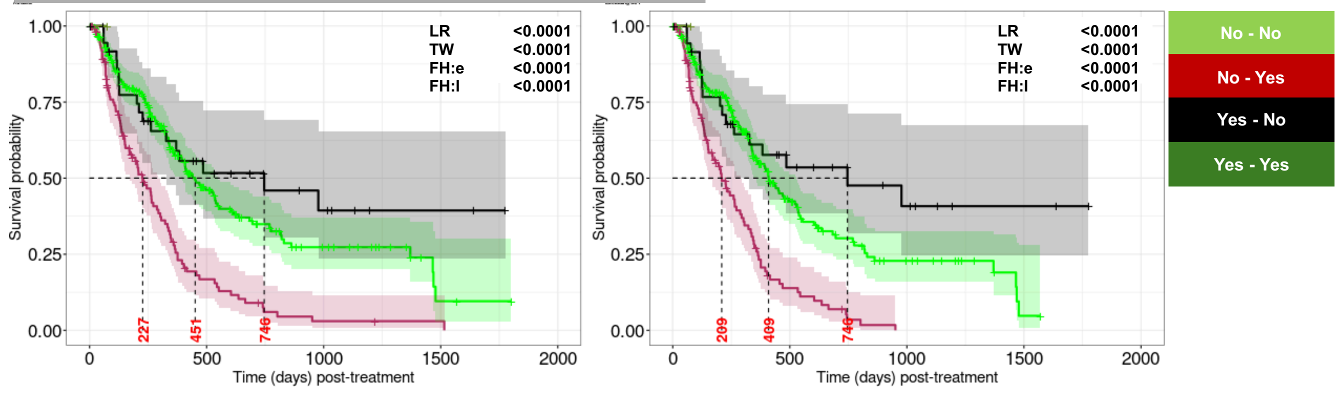


Number at risk (number censored) — left panel

| | 0 | 500 | 1000 | 1500 | 2000 |
|---|---|---|---|---|---|
| No - No | 229 (0) | 55 (79) | 17 (99) | 2 (110) | 0 (112) |
| No - Yes | 117 (0) | 13 (25) | 2 (26) | 1 (27) | 0 (27) |
| Yes - No | 38 (0) | 13 (9) | 6 (14) | 2 (18) | 0 (20) |
| Yes - Yes | 1 (0) | 0 (1) | 0 (1) | 0 (1) | 0 (1) |

Number at risk (number censored) — right panel

| | 0 | 500 | 1000 | 1500 | 2000 |
|---|---|---|---|---|---|
| No - No | 216 (0) | 47 (75) | 14 (90) | 1 (99) | 0 (100) |
| No - Yes | 113 (0) | 10 (25) | 0 (26) | 0 (26) | 0 (26) |
| Yes - No | 37 (0) | 13 (9) | 6 (14) | 2 (18) | 0 (20) |
| Yes - Yes | 1 (0) | 0 (1) | 0 (1) | 0 (1) | 0 (1) |

#### *TP53 + IDH1/2*

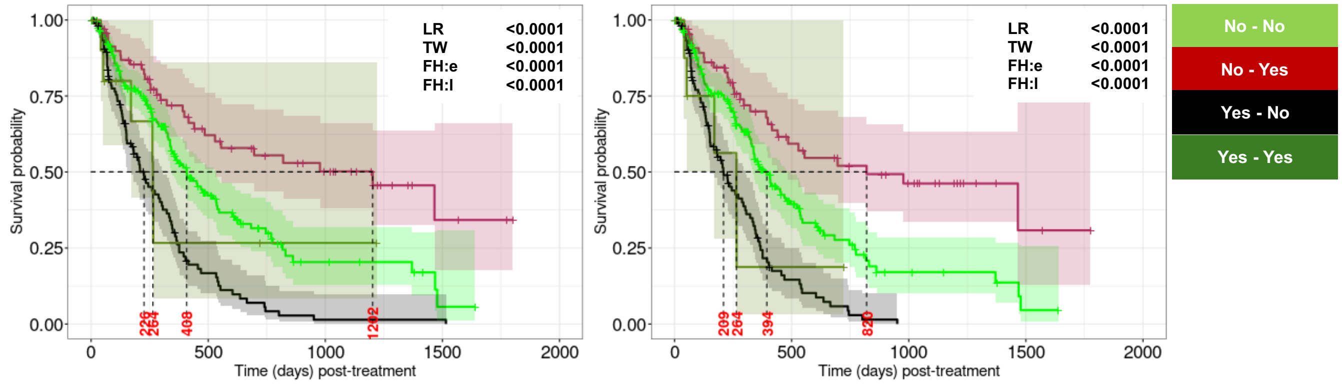


Number at risk (number censored) — left panel

| | 0 | 500 | 1000 | 1500 | 2000 |
|---|---|---|---|---|---|
| No - No | 196 (0) | 41 (68) | 8 (86) | 1 (90) | 0 (91) |
| No - Yes | 71 (0) | 30 (18) | 17 (26) | 3 (38) | 0 (41) |
| Yes - No | 110 (0) | 12 (24) | 1 (24) | 1 (24) | 0 (24) |
| Yes - Yes | 12 (0) | 2 (4) | 1 (5) | 0 (6) | 0 (6) |

Number at risk (number censored) — right panel

| | 0 | 500 | 1000 | 1500 | 2000 |
|---|---|---|---|---|---|
| No - No | 185 (0) | 36 (64) | 7 (78) | 1 (81) | 0 (82) |
| No - Yes | 67 (0) | 26 (18) | 14 (25) | 2 (36) | 0 (38) |
| Yes - No | 108 (0) | 10 (24) | 0 (24) | 0 (24) | 0 (24) |
| Yes - Yes | 10 (0) | 1 (4) | 0 (5) | 0 (5) | 0 (5) |

#### *TP53 + Good cytogenetics*

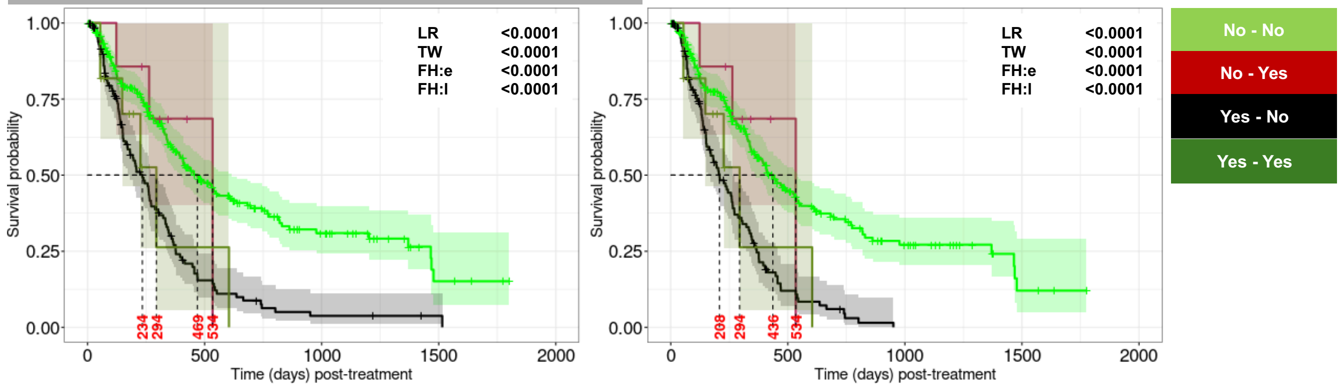


Number at risk (number censored) — left panel

| | 0 | 500 | 1000 | 1500 | 2000 |
|---|---|---|---|---|---|
| No - No | 265 (0) | 70 (85) | 25 (111) | 4 (127) | 0 (131) |
| No - Yes | 7 (0) | 1 (4) | 0 (4) | 0 (4) | 0 (4) |
| Yes - No | 136 (0) | 14 (28) | 3 (29) | 1 (31) | 0 (31) |
| Yes - Yes | 11 (0) | 1 (5) | 0 (5) | 0 (5) | 0 (5) |

Number at risk (number censored) — right panel

| | 0 | 500 | 1000 | 1500 | 2000 |
|---|---|---|---|---|---|
| No - No | 250 (0) | 61 (81) | 21 (102) | 3 (116) | 0 (119) |
| No - Yes | 7 (0) | 1 (4) | 0 (4) | 0 (4) | 0 (4) |
| Yes - No | 128 (0) | 10 (27) | 0 (28) | 0 (28) | 0 (28) |
| Yes - Yes | 11 (0) | 1 (5) | 0 (5) | 0 (5) | 0 (5) |

### Supplemental Figure 5, cont'd

#### *TP53 + IDH1/IDH2/NPM1/Good cytogenetics*

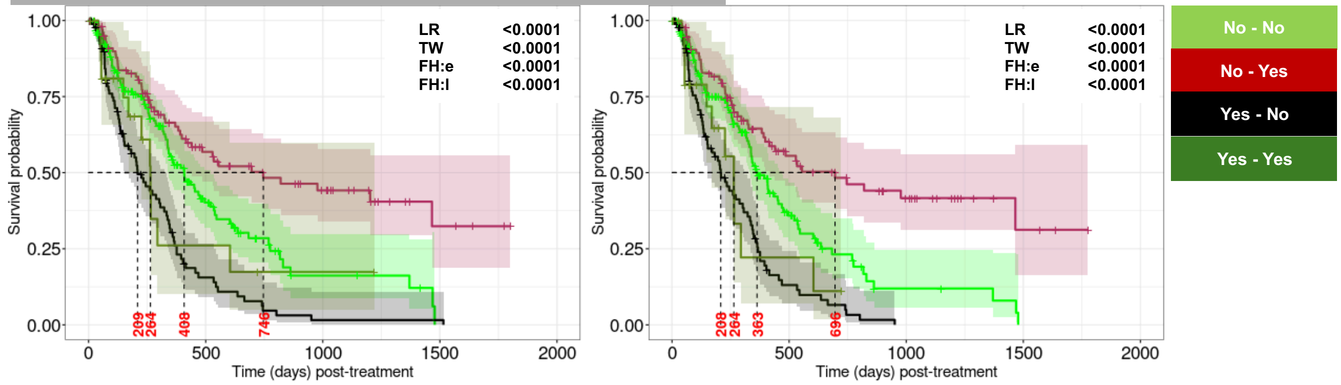


Number at risk (number censored)

| | | | | |
|---|---|---|---|---|
| 163 (0) | 32 (58) | 5 (72) | 0 (74) | 0 (74) |
| 102 (0) | 37 (27) | 19 (38) | 4 (51) | 0 (55) |
| 95 (0) | 10 (19) | 1 (19) | 1 (19) | 0 (19) |
| 23 (0) | 3 (9) | 1 (10) | 0 (11) | 0 (11) |

Number at risk (number censored)

| | | | | |
|---|---|---|---|---|
| 153 (0) | 27 (54) | 4 (64) | 0 (65) | 0 (65) |
| 97 (0) | 33 (27) | 16 (37) | 3 (49) | 0 (52) |
| 93 (0) | 8 (19) | 0 (19) | 0 (19) | 0 (19) |
| 21 (0) | 2 (9) | 0 (10) | 0 (10) | 0 (10) |

#### *NPM1 + FLT3-ITD*

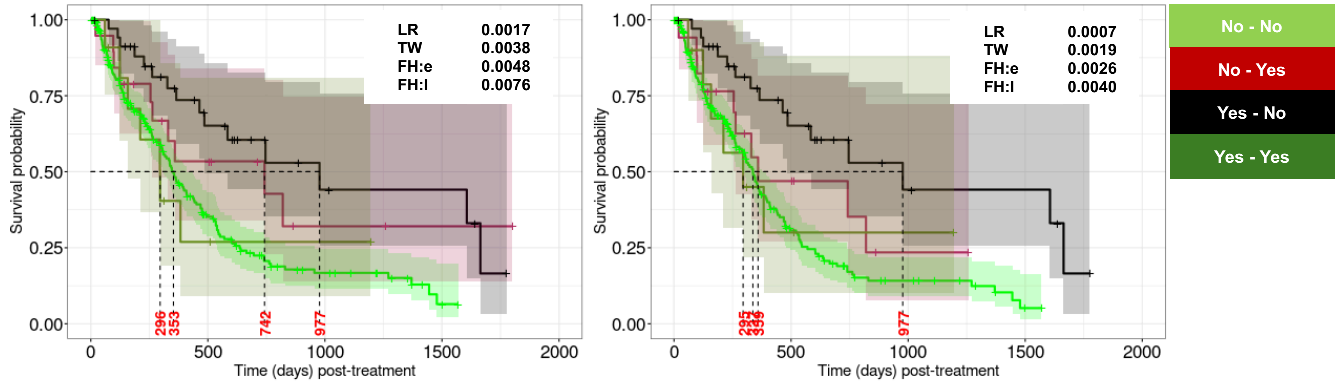


Number at risk (number censored)

| | | | | |
|---|---|---|---|---|
| 226 (0) | 49 (58) | 15 (69) | 1 (79) | 0 (80) |
| 19 (0) | 8 (3) | 2 (7) | 1 (8) | 0 (9) |
| 35 (0) | 15 (10) | 5 (17) | 4 (18) | 0 (20) |
| 11 (0) | 2 (2) | 1 (3) | 0 (4) | 0 (4) |

Number at risk (number censored)

| | | | | |
|---|---|---|---|---|
| 213 (0) | 40 (55) | 12 (63) | 1 (70) | 0 (71) |
| 17 (0) | 6 (3) | 1 (6) | 0 (7) | 0 (7) |
| 35 (0) | 15 (10) | 5 (17) | 4 (18) | 0 (20) |
| 10 (0) | 2 (2) | 1 (3) | 0 (4) | 0 (4) |

#### *IDH1/2 + FLT3-ITD*

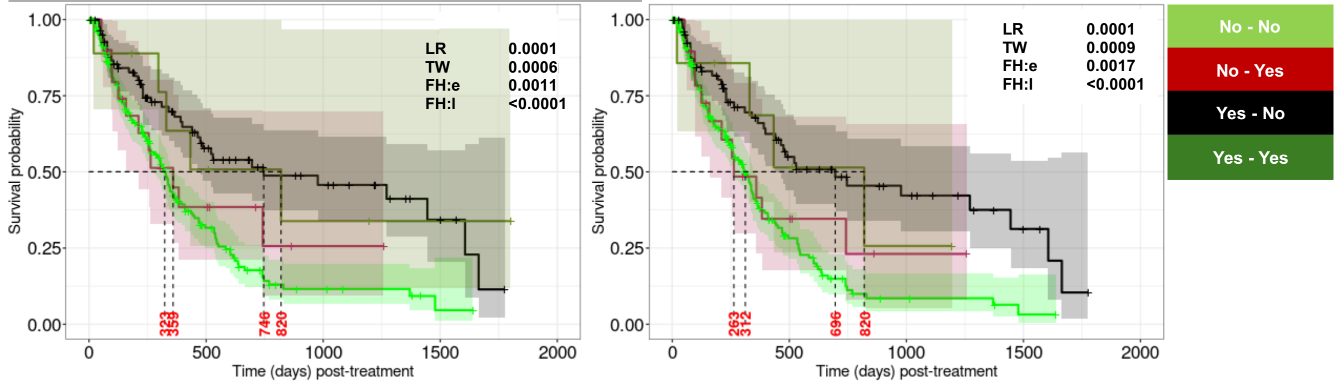


Number at risk (number censored)

| | | | | |
|---|---|---|---|---|
| 209 (0) | 37 (56) | 7 (66) | 1 (70) | 0 (71) |
| 20 (0) | 6 (3) | 1 (7) | 0 (8) | 0 (8) |
| 87 (0) | 31 (26) | 15 (37) | 4 (46) | 0 (48) |
| 9 (0) | 4 (1) | 2 (2) | 1 (3) | 0 (4) |

Number at risk (number censored)

| | | | | |
|---|---|---|---|---|
| 197 (0) | 31 (53) | 5 (60) | 1 (62) | 0 (63) |
| 19 (0) | 5 (3) | 1 (6) | 0 (7) | 0 (7) |
| 82 (0) | 27 (25) | 13 (34) | 4 (41) | 0 (43) |
| 7 (0) | 3 (1) | 1 (2) | 0 (3) | 0 (3) |

#### Supplemental Figure 5, cont'd

##### *FLT3-ITD + Good risk cytogenetics*

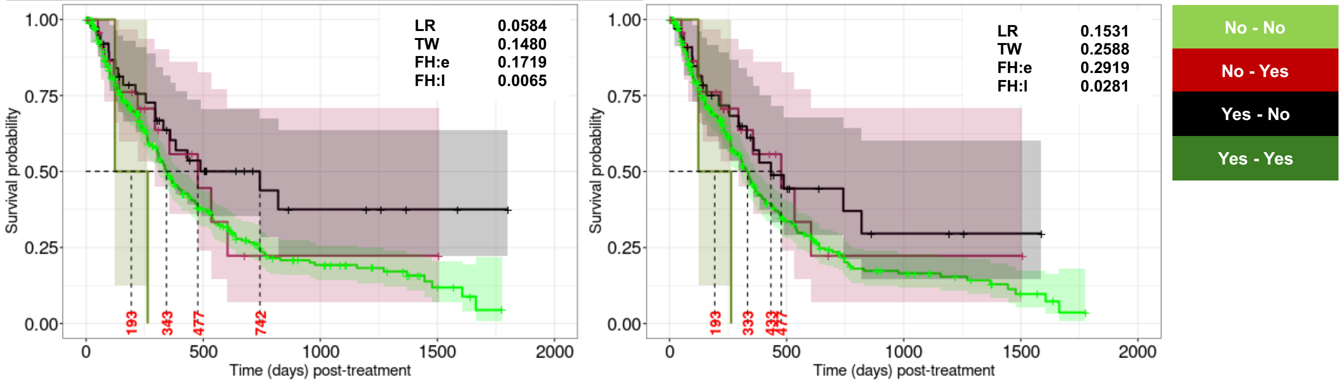


Number at risk (number censored)

| | | | | |
|---|---|---|---|---|
| 343 (0) | 77 (92) | 25 (113) | 5 (128) | 0 (131) |
| 24 (0) | 4 (11) | 1 (12) | 1 (12) | 0 (13) |
| 38 (0) | 14 (7) | 5 (14) | 2 (17) | 0 (19) |
| 2 (0) | 0 (0) | 0 (0) | 0 (0) | 0 (0) |

Number at risk (number censored)

| | | | | |
|---|---|---|---|---|
| 323 (0) | 64 (88) | 20 (104) | 5 (114) | 0 (117) |
| 24 (0) | 4 (11) | 1 (12) | 1 (12) | 0 (13) |
| 33 (0) | 10 (7) | 3 (12) | 1 (14) | 0 (15) |
| 2 (0) | 0 (0) | 0 (0) | 0 (0) | 0 (0) |

##### *IDH1/IDH2/NPM1/Good risk cytogenetics + FLT3-ITD*

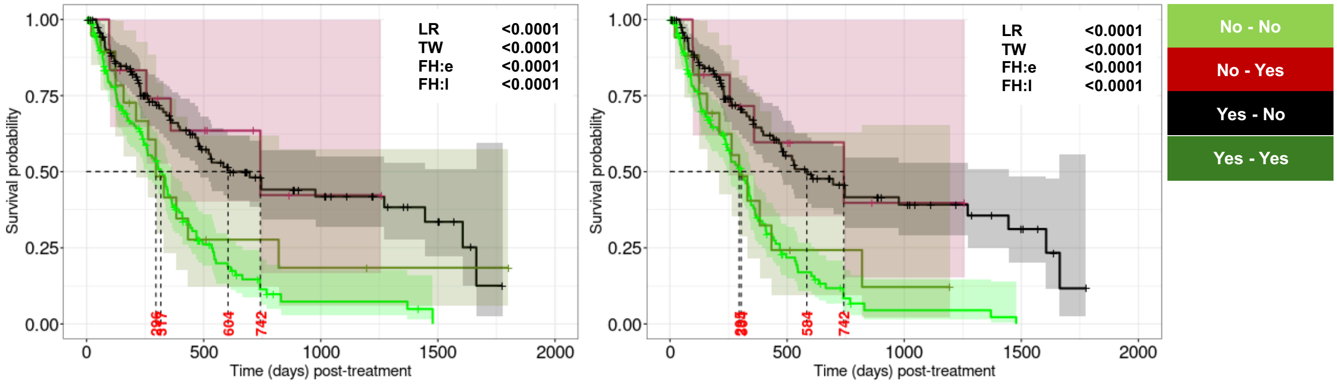


Number at risk (number censored)

| | | | | |
|---|---|---|---|---|
| 146 (0) | 22 (37) | 3 (43) | 0 (44) | 0 (44) |
| 12 (0) | 6 (2) | 1 (6) | 0 (7) | 0 (7) |
| 127 (0) | 42 (42) | 18 (57) | 6 (67) | 0 (71) |
| 19 (0) | 4 (3) | 2 (4) | 1 (5) | 0 (6) |

Number at risk (number censored)

| | | | | |
|---|---|---|---|---|
| 138 (0) | 18 (34) | 2 (38) | 0 (38) | 0 (38) |
| 11 (0) | 5 (2) | 1 (5) | 0 (6) | 0 (6) |
| 122 (0) | 38 (41) | 16 (54) | 6 (62) | 0 (66) |
| 17 (0) | 3 (3) | 1 (4) | 0 (5) | 0 (5) |

##### *IDH1/IDH2/NPM1/Good cytogenetics + Complex cytogenetics*

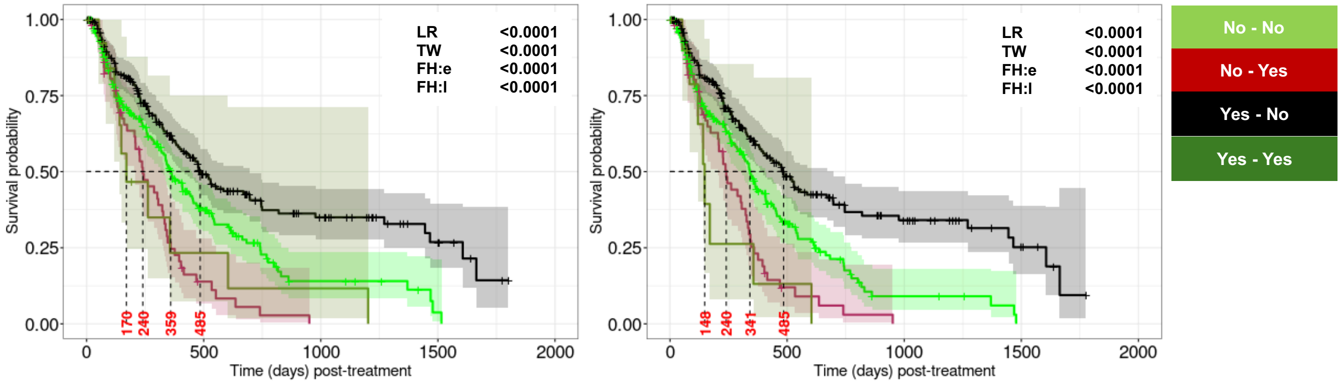


Number at risk (number censored)

| | | | | |
|---|---|---|---|---|
| 228 (0) | 45 (71) | 8 (88) | 1 (92) | 0 (92) |
| 59 (0) | 5 (10) | 0 (10) | 0 (10) | 0 (10) |
| 224 (0) | 66 (65) | 27 (89) | 8 (105) | 0 (111) |
| 14 (0) | 2 (4) | 1 (4) | 0 (4) | 0 (4) |

Number at risk (number censored)

| | | | | |
|---|---|---|---|---|
| 215 (0) | 37 (67) | 5 (79) | 0 (81) | 0 (81) |
| 58 (0) | 4 (10) | 0 (10) | 0 (10) | 0 (10) |
| 213 (0) | 59 (65) | 23 (87) | 7 (100) | 0 (105) |
| 11 (0) | 1 (3) | 0 (3) | 0 (3) | 0 (3) |

#### Supplemental Figure 5, cont'd

##### *TP53 + complex cytogenetics*

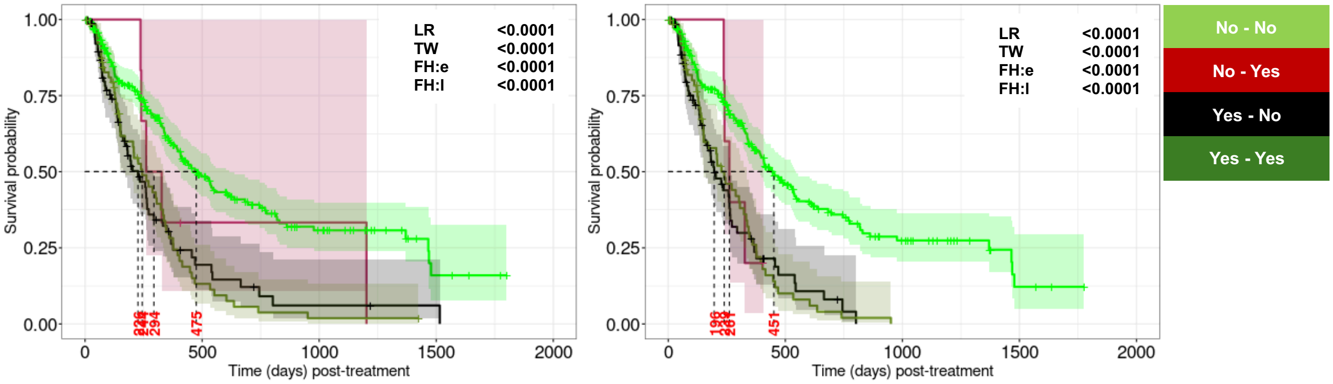


Number at risk (number censored)

| | | | | |
|---|---|---|---|---|
| 266 (0) | 70 (88) | 24 (114) | 4 (130) | 0 (134) |
| 6 (0) | 1 (1) | 1 (1) | 0 (1) | 0 (1) |
| 80 (0) | 8 (22) | 2 (23) | 1 (24) | 0 (24) |
| 67 (0) | 7 (11) | 1 (11) | 0 (12) | 0 (12) |

Number at risk (number censored)

| | | | | |
|---|---|---|---|---|
| 252 (0) | 62 (84) | 21 (105) | 3 (119) | 0 (122) |
| 5 (0) | 0 (1) | 0 (1) | 0 (1) | 0 (1) |
| 75 (0) | 6 (21) | 0 (22) | 0 (22) | 0 (22) |
| 64 (0) | 5 (11) | 0 (11) | 0 (11) | 0 (11) |

##### *TP53 + Minus5*

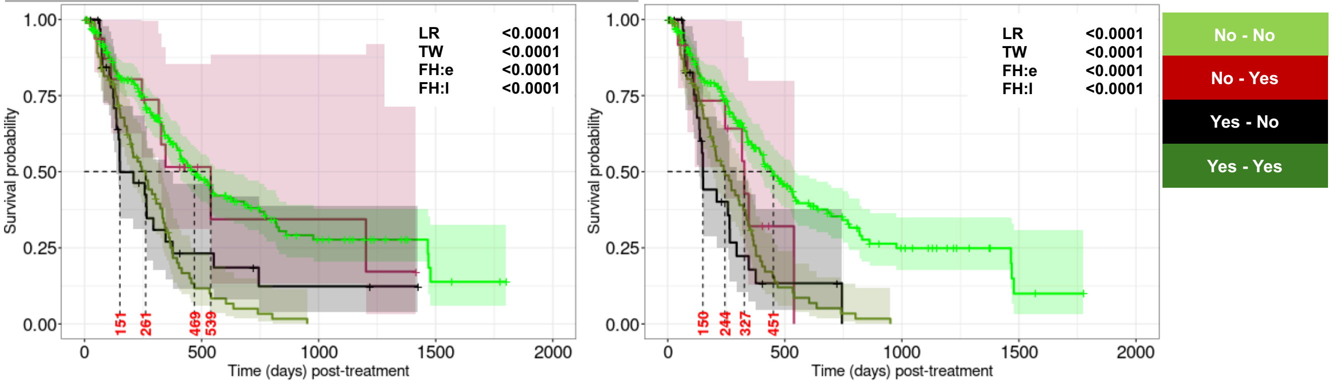


Number at risk (number censored)

| | | | | |
|---|---|---|---|---|
| 225 (0) | 58 (75) | 18 (97) | 3 (109) | 0 (112) |
| 16 (0) | 3 (6) | 2 (6) | 0 (7) | 0 (7) |
| 35 (0) | 5 (8) | 2 (9) | 0 (11) | 0 (11) |
| 85 (0) | 7 (17) | 0 (17) | 0 (17) | 0 (17) |

Number at risk (number censored)

| | | | | |
|---|---|---|---|---|
| 216 (0) | 52 (73) | 16 (91) | 2 (102) | 0 (104) |
| 12 (0) | 1 (4) | 0 (4) | 0 (4) | 0 (4) |
| 32 (0) | 2 (8) | 0 (9) | 0 (9) | 0 (9) |
| 81 (0) | 7 (16) | 0 (16) | 0 (16) | 0 (16) |

##### *TP53 + Inv3*

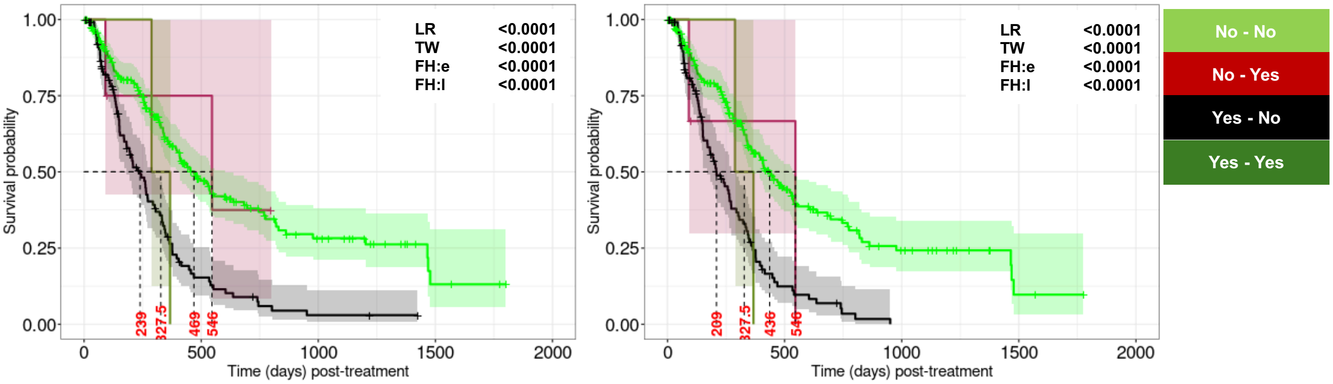


Number at risk (number censored)

| | | | | |
|---|---|---|---|---|
| 237 (0) | 59 (80) | 20 (101) | 3 (114) | 0 (117) |
| 4 (0) | 2 (1) | 0 (2) | 0 (2) | 0 (2) |
| 118 (0) | 12 (25) | 2 (26) | 0 (28) | 0 (28) |
| 2 (0) | 0 (0) | 0 (0) | 0 (0) | 0 (0) |

Number at risk (number censored)

| | | | | |
|---|---|---|---|---|
| 225 (0) | 52 (76) | 16 (94) | 2 (105) | 0 (107) |
| 3 (0) | 1 (1) | 0 (1) | 0 (1) | 0 (1) |
| 111 (0) | 9 (24) | 0 (25) | 0 (25) | 0 (25) |
| 2 (0) | 0 (0) | 0 (0) | 0 (0) | 0 (0) |

#### Supplemental Figure 5, cont'd

##### *TP53 + Minus17*

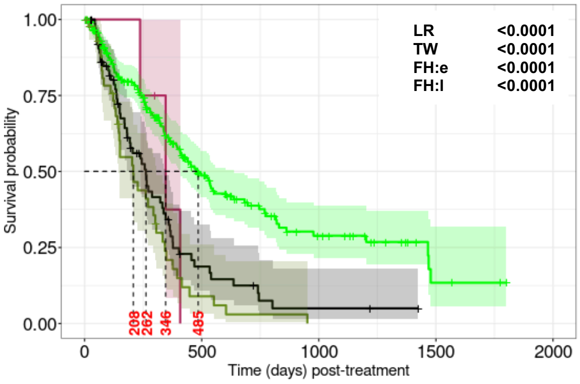


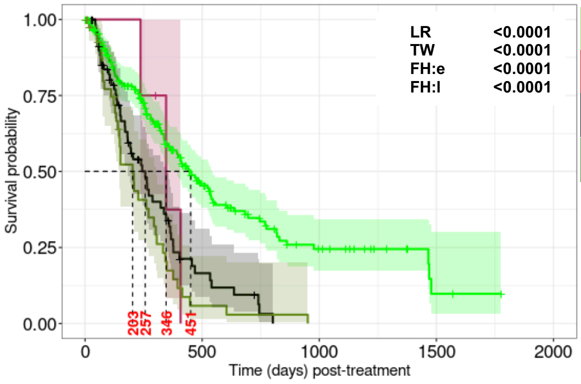


No - No
No - Yes
Yes - No
Yes - Yes

Number at risk (number censored)

| | | | | |
|---|---|---|---|---|
| 236 (0) | 61 (80) | 20 (102) | 3 (115) | 0 (118) |
| 4 (0) | 0 (1) | 0 (1) | 0 (1) | 0 (1) |
| 76 (0) | 9 (18) | 2 (19) | 0 (21) | 0 (21) |
| 45 (0) | 3 (8) | 0 (8) | 0 (8) | 0 (8) |

Number at risk (number censored)

| | | | | |
|---|---|---|---|---|
| 223 (0) | 53 (76) | 16 (94) | 2 (105) | 0 (107) |
| 4 (0) | 0 (1) | 0 (1) | 0 (1) | 0 (1) |
| 71 (0) | 7 (18) | 0 (19) | 0 (19) | 0 (19) |
| 43 (0) | 2 (7) | 0 (7) | 0 (7) | 0 (7) |

##### *TP53 + Minus7*

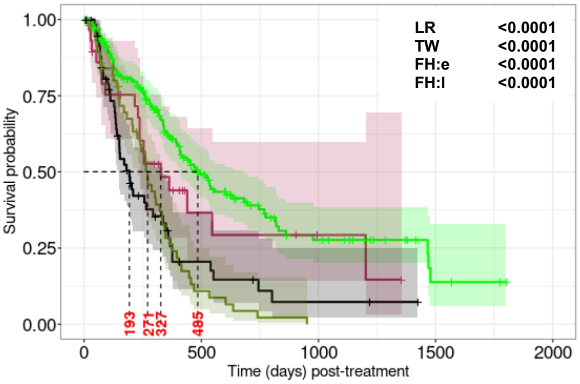


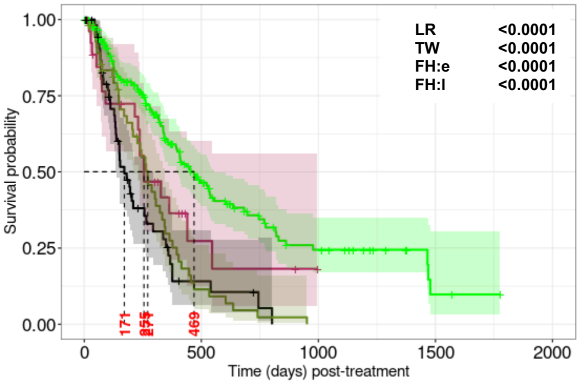


No - No
No - Yes
Yes - No
Yes - Yes

Number at risk (number censored)

| | | | | |
|---|---|---|---|---|
| 210 (0) | 56 (71) | 18 (91) | 3 (103) | 0 (106) |
| 31 (0) | 5 (10) | 2 (12) | 0 (13) | 0 (13) |
| 61 (0) | 7 (14) | 2 (15) | 0 (17) | 0 (17) |
| 59 (0) | 5 (11) | 0 (11) | 0 (11) | 0 (11) |

Number at risk (number censored)

| | | | | |
|---|---|---|---|---|
| 200 (0) | 50 (68) | 16 (84) | 2 (95) | 0 (97) |
| 28 (0) | 3 (9) | 0 (11) | 0 (11) | 0 (11) |
| 56 (0) | 4 (13) | 0 (14) | 0 (14) | 0 (14) |
| 57 (0) | 5 (11) | 0 (11) | 0 (11) | 0 (11) |

##### *TP53 + FLT3-ITD*

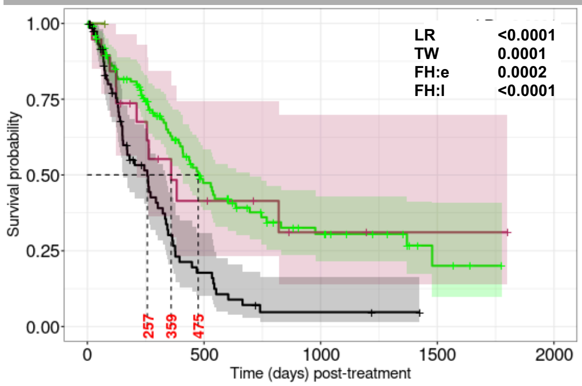


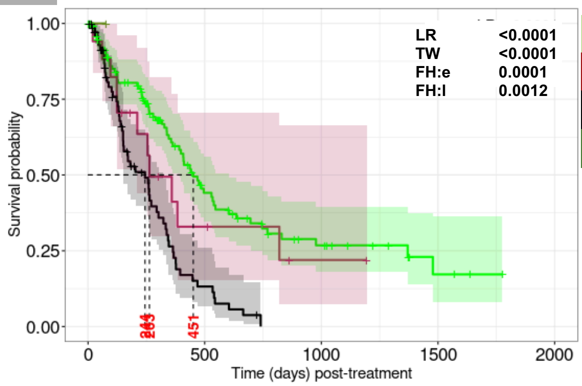


No - No
No - Yes
Yes - No
Yes - Yes

Number at risk (number censored)

| | | | | |
|---|---|---|---|---|
| 128 (0) | 36 (38) | 15 (48) | 3 (58) | 0 (61) |
| 19 (0) | 6 (3) | 2 (6) | 1 (7) | 0 (8) |
| 78 (0) | 10 (17) | 2 (18) | 0 (20) | 0 (20) |
| 1 (0) | 0 (1) | 0 (1) | 0 (1) | 0 (1) |

Number at risk (number censored)

| | | | | |
|---|---|---|---|---|
| 120 (0) | 32 (35) | 13 (43) | 3 (51) | 0 (54) |
| 17 (0) | 4 (3) | 1 (5) | 0 (6) | 0 (6) |
| 75 (0) | 7 (17) | 0 (18) | 0 (18) | 0 (18) |
| 1 (0) | 0 (1) | 0 (1) | 0 (1) | 0 (1) |

#### Supplemental Figure 5, cont'd

##### *FLT3-ITD + KRAS*

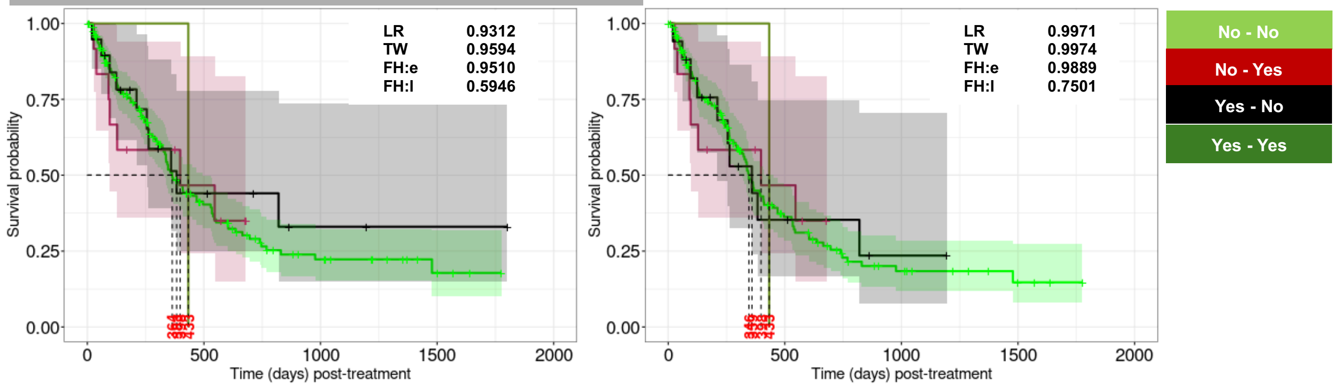


Number at risk (number censored)

| | | | | | |
|---|---|---|---|---|---|
| green | 164 (0) | 41 (43) | 14 (54) | 3 (64) | 0 (67) |
| red | 12 (0) | 4 (2) | 0 (5) | 0 (5) | 0 (5) |
| black | 19 (0) | 6 (4) | 2 (7) | 1 (8) | 0 (9) |
| dark green | 1 (0) | 0 (0) | 0 (0) | 0 (0) | 0 (0) |

Number at risk (number censored)

| | | | | | |
|---|---|---|---|---|---|
| green | 154 (0) | 35 (40) | 11 (49) | 3 (56) | 0 (59) |
| red | 12 (0) | 4 (2) | 0 (5) | 0 (5) | 0 (5) |
| black | 17 (0) | 4 (4) | 1 (6) | 0 (7) | 0 (7) |
| dark green | 1 (0) | 0 (0) | 0 (0) | 0 (0) | 0 (0) |

##### *FLT3-ITD + NRAS*

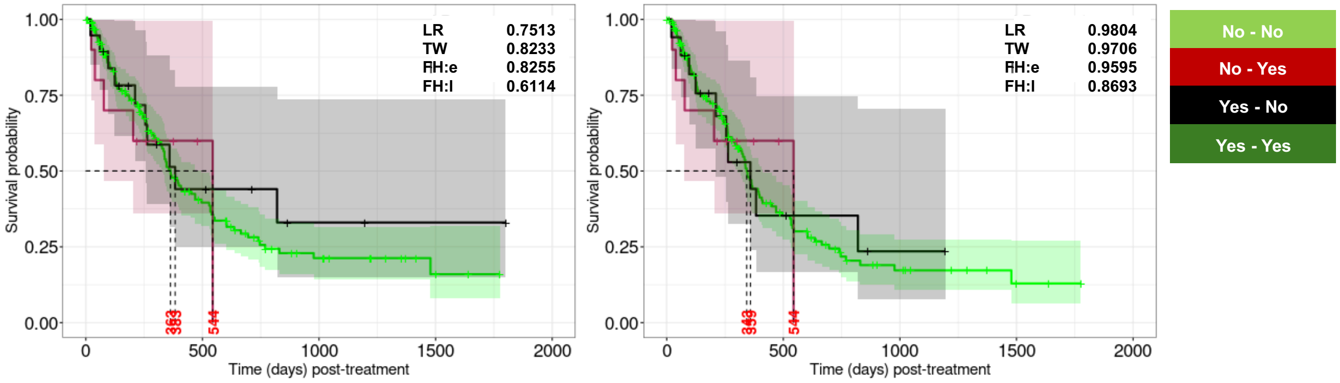


Number at risk (number censored)

| | | | | | |
|---|---|---|---|---|---|
| green | 158 (0) | 40 (40) | 13 (51) | 2 (61) | 0 (63) |
| red | 10 (0) | 1 (5) | 0 (5) | 0 (5) | 0 (5) |
| black | 19 (0) | 6 (4) | 2 (7) | 1 (8) | 0 (9) |

Number at risk (number censored)

| | | | | | |
|---|---|---|---|---|---|
| green | 148 (0) | 34 (37) | 10 (46) | 2 (53) | 0 (55) |
| red | 10 (0) | 1 (5) | 0 (5) | 0 (5) | 0 (5) |
| black | 17 (0) | 4 (4) | 1 (6) | 0 (7) | 0 (7) |

**Supplemental Figure 6. RRM in different CU cohorts.**

**A. RRM: CU CCAS with allo-HCT included (left) and excluded (right) recipients.**

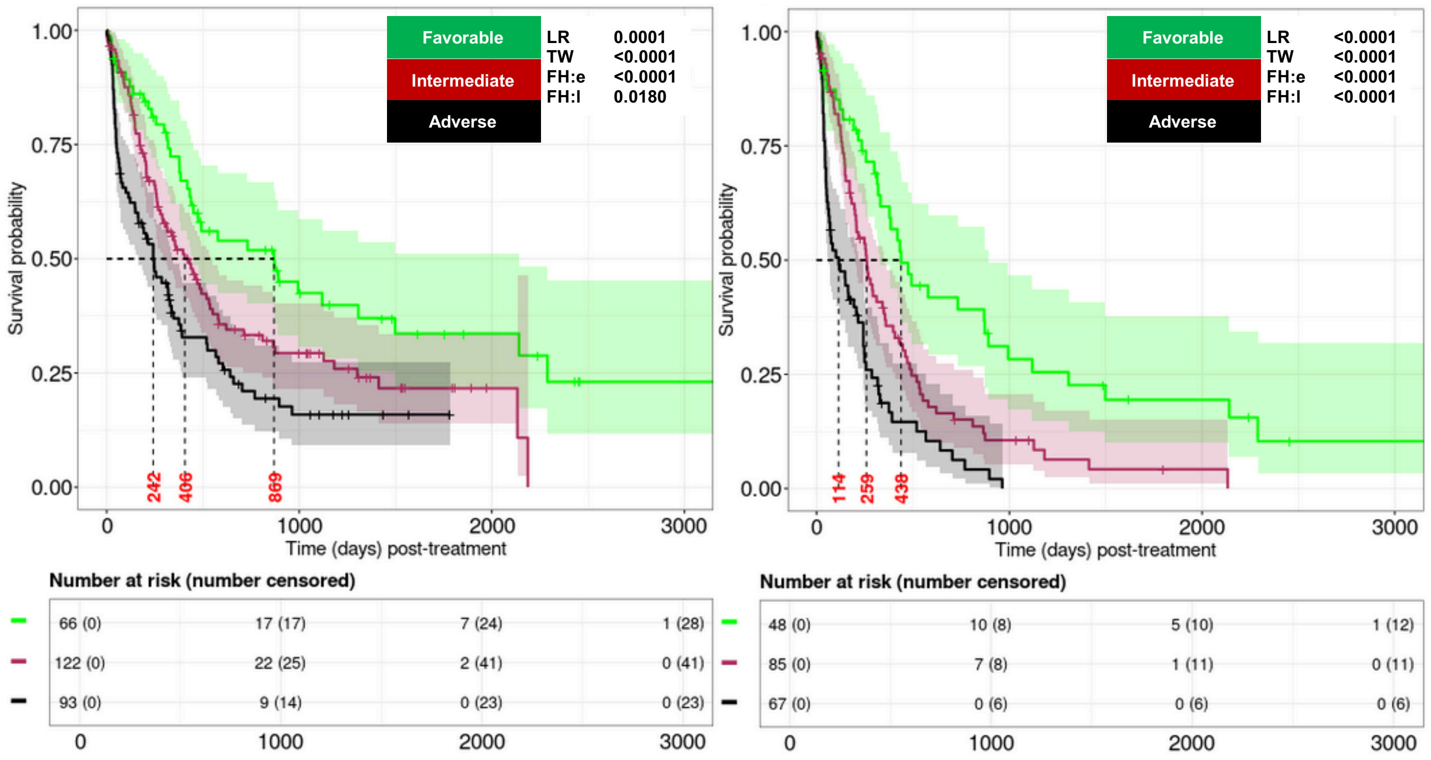


**B. RRM: CU IAS with allo-HCT included (left panel) and excluded (right panel) recipients.**

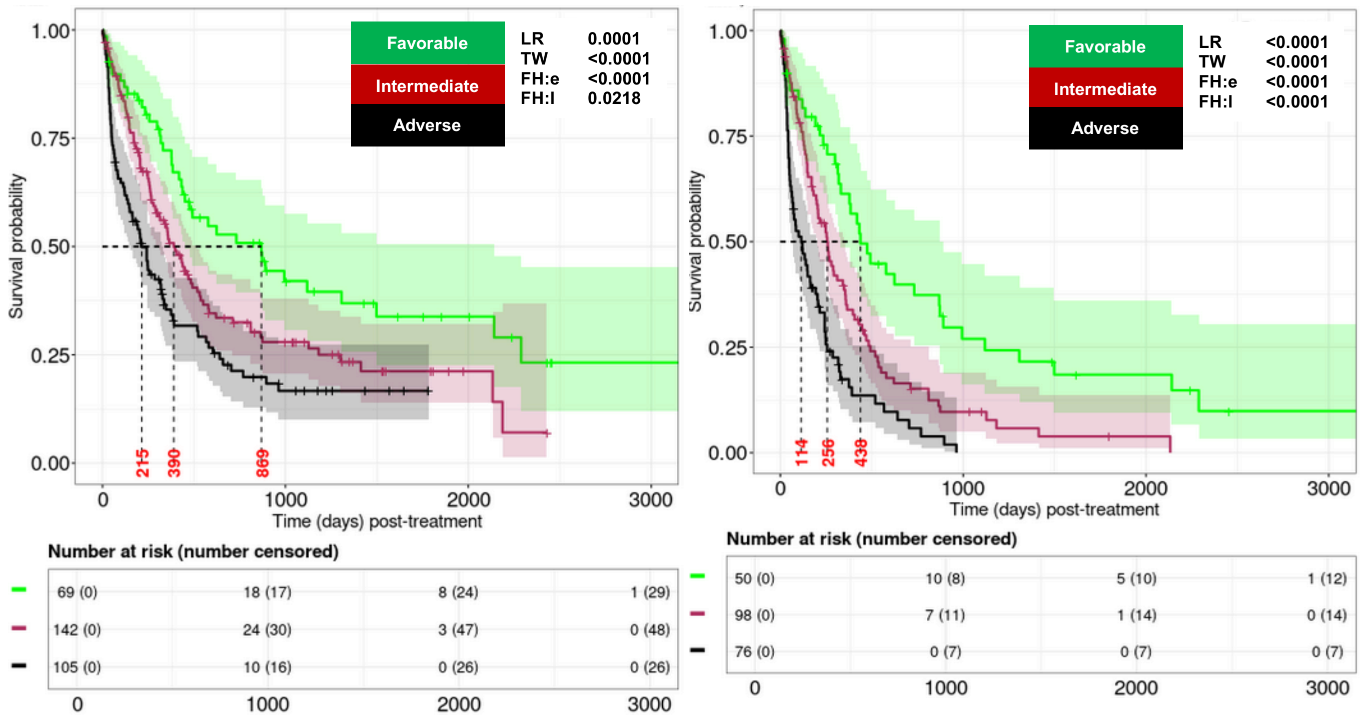

**Supplemental Figure 7. Application of mPRS in CU cohorts.**

**A. mPRS: CU FAS with allo-HCT included (left) and excluded (right) recipients.**

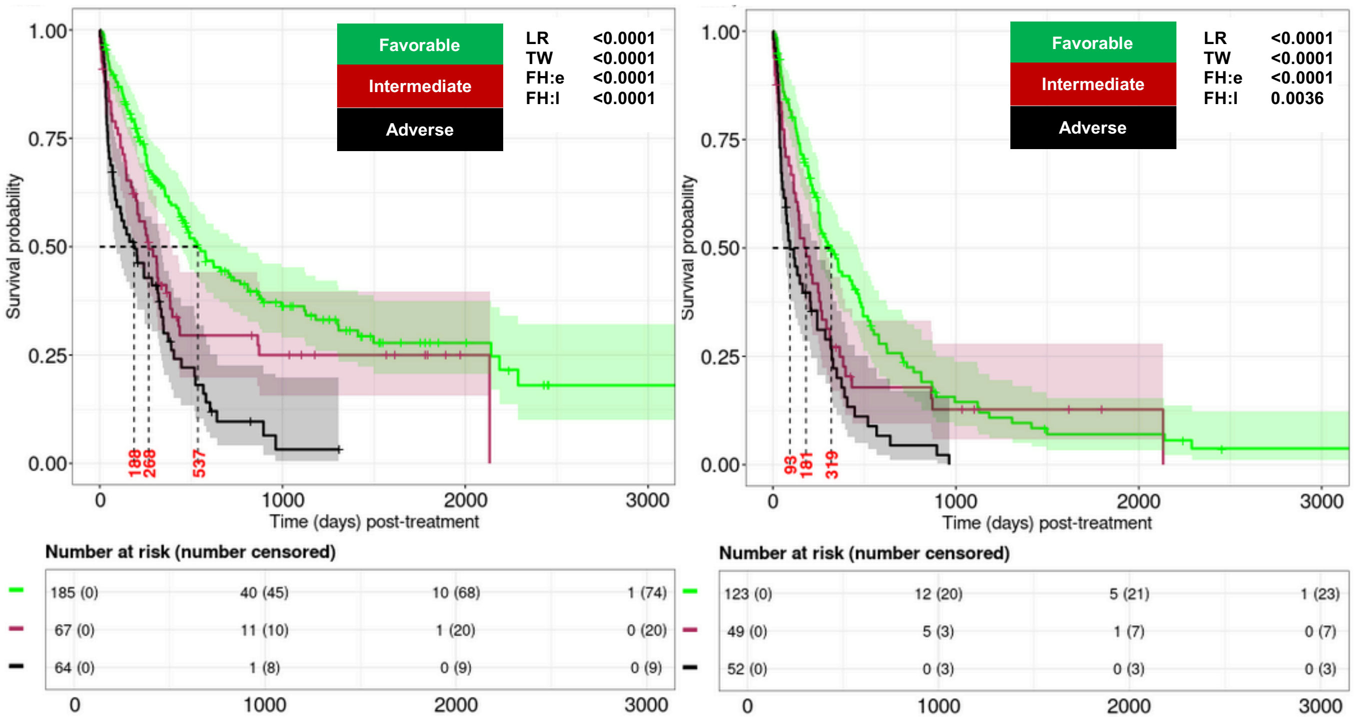


**B. mPRS: CU IAS with allo-HCT included (left) and excluded (right) recipients.**

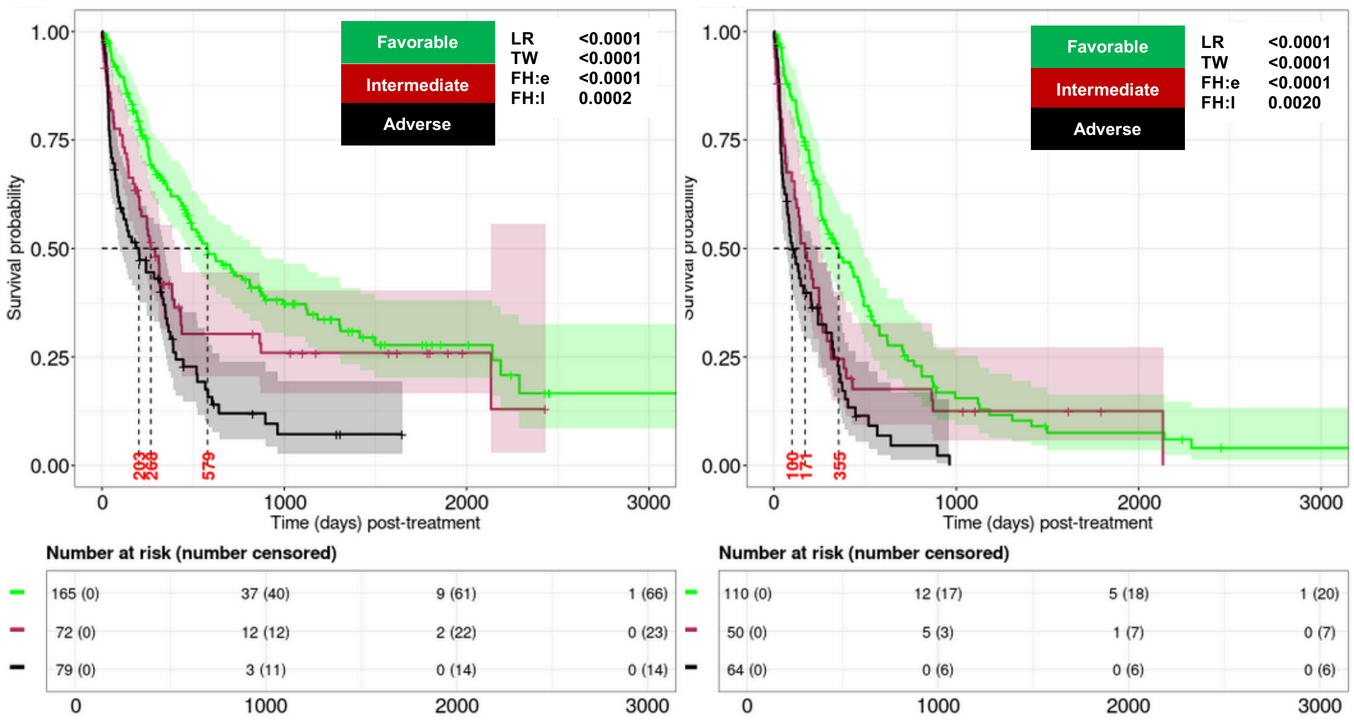

**Supplemental Figure 7, cont'd**

**C. Extended mPRS (e-mPRS): CU CCAS with allo-HCT included (left) and excluded (right) recipients.**

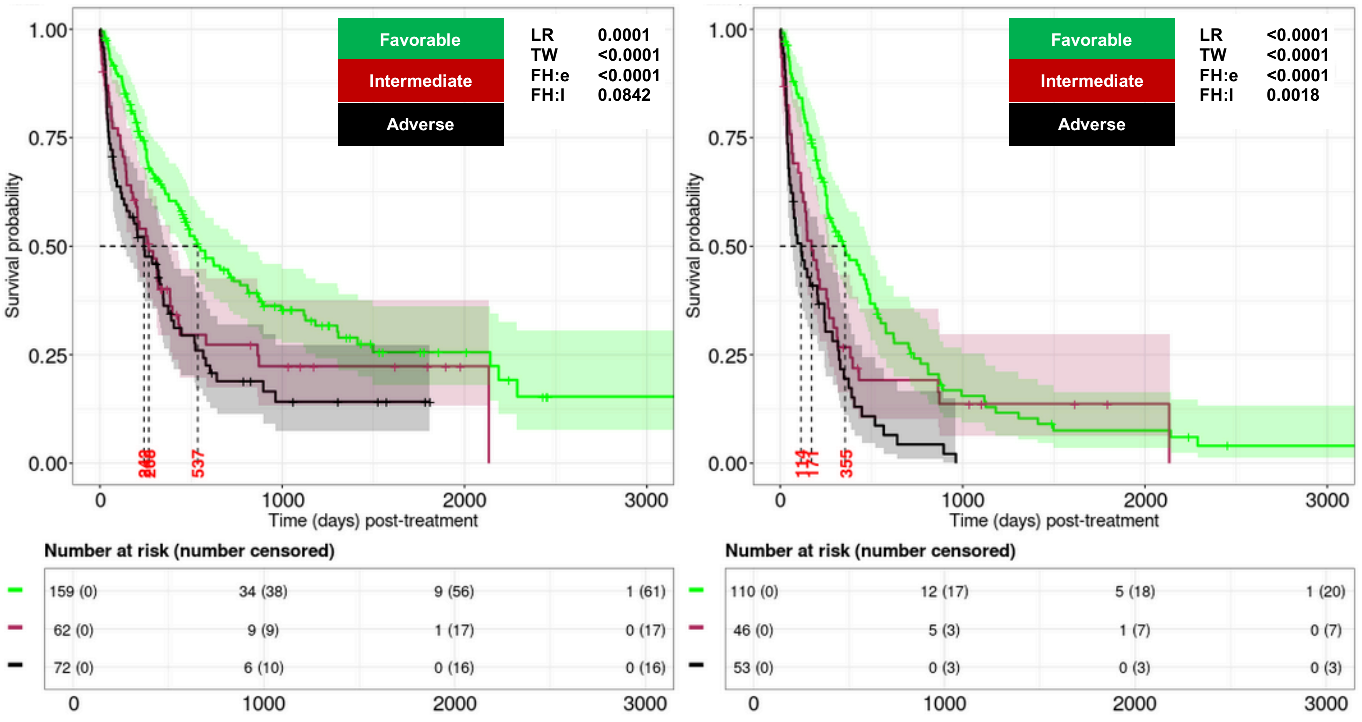


**D. e-mPRS: CU FAS with allo-HCT included (left) and excluded (right) recipients.**

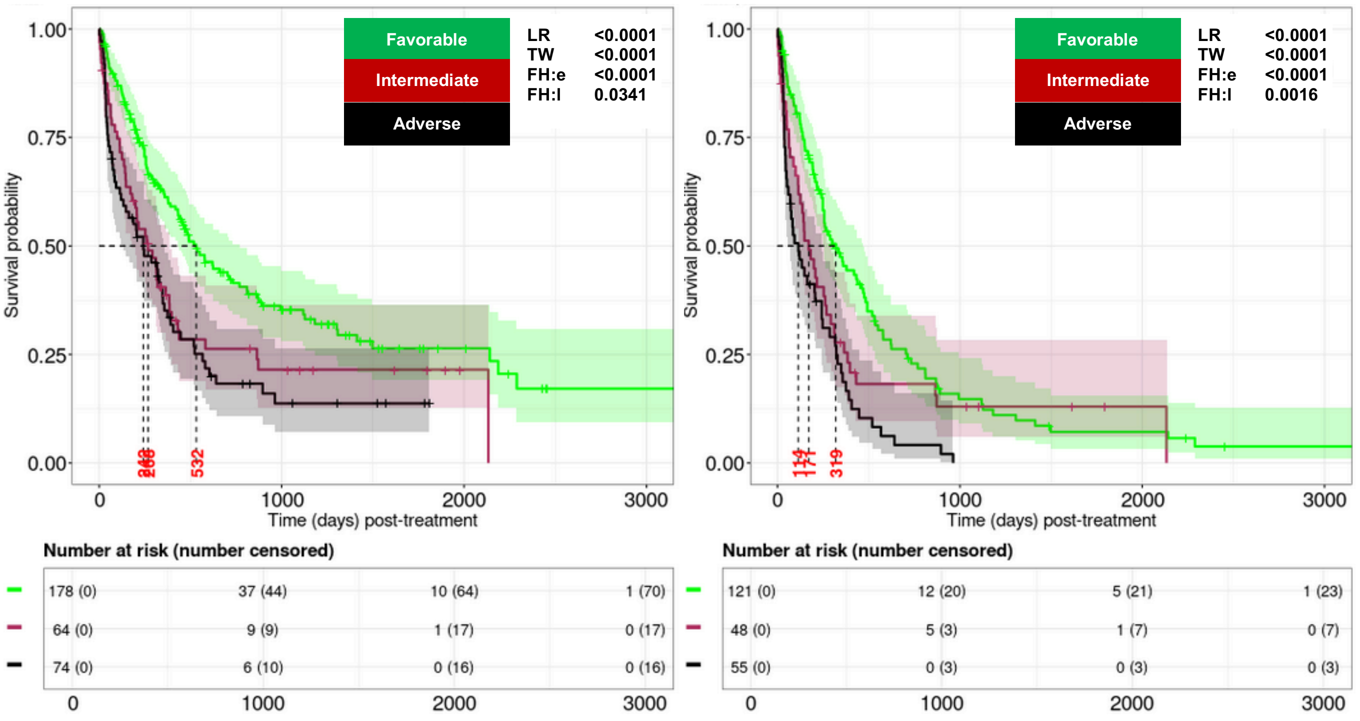

**Supplemental Figure 7, cont'd**

**E. e-mPRS: CU IAS with allo-HCT included (left) and excluded (right) recipients.**

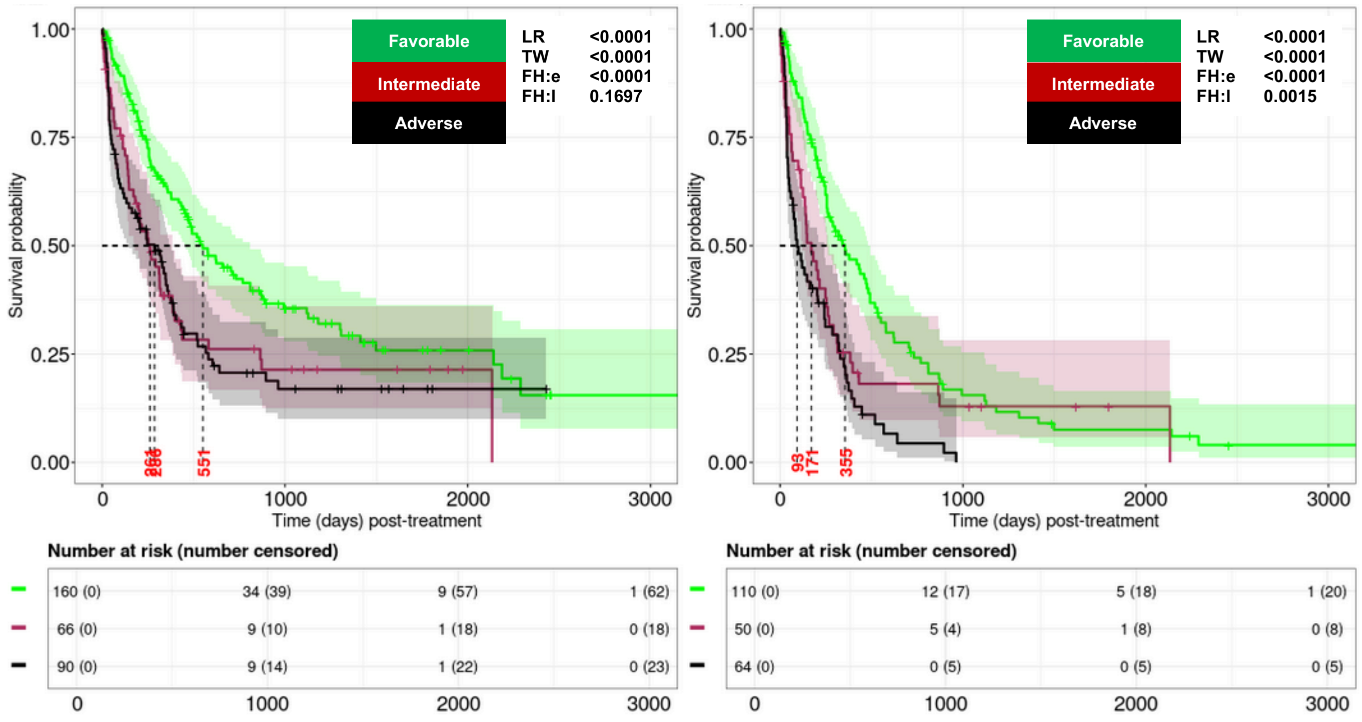

**Supplemental Figure 7, cont'd**

**F. Pairwise comparisons between the RRM and e-mPRS in the CU dCCAS cohort (allo-HCT patients excluded).**

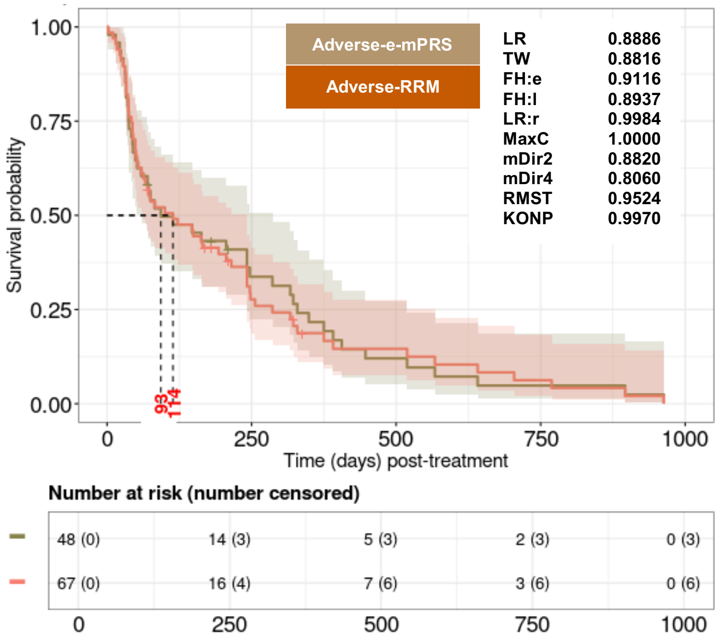


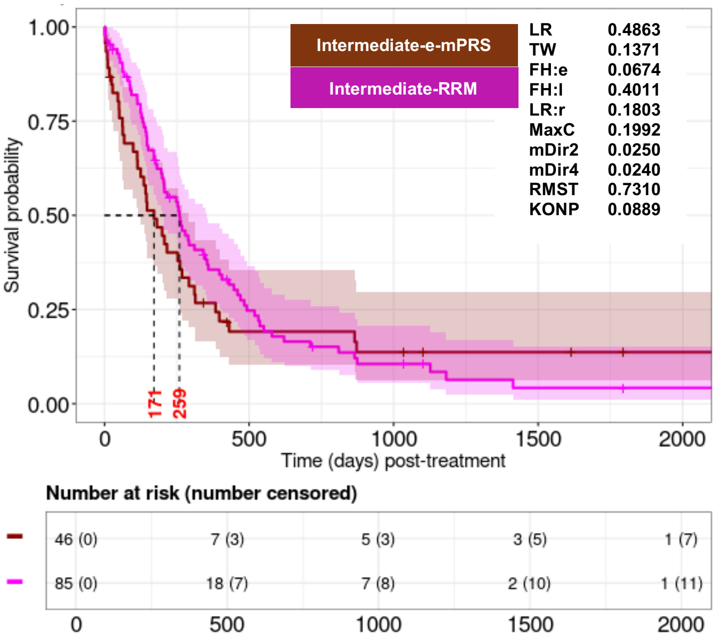


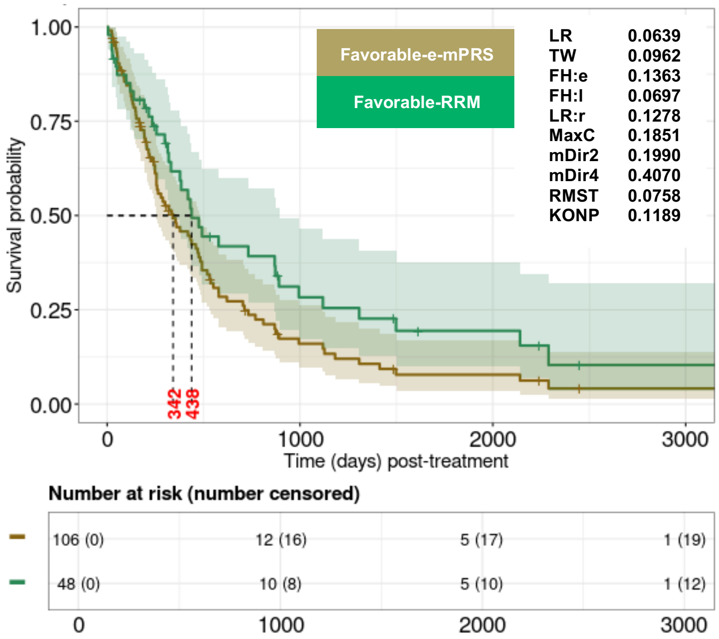

**Supplemental Figure 8: RRM and ELN22 comparisons in the RWC.**

**A. RRM: RWC IAS with allo-HCT included (left) and excluded (right) recipients.**

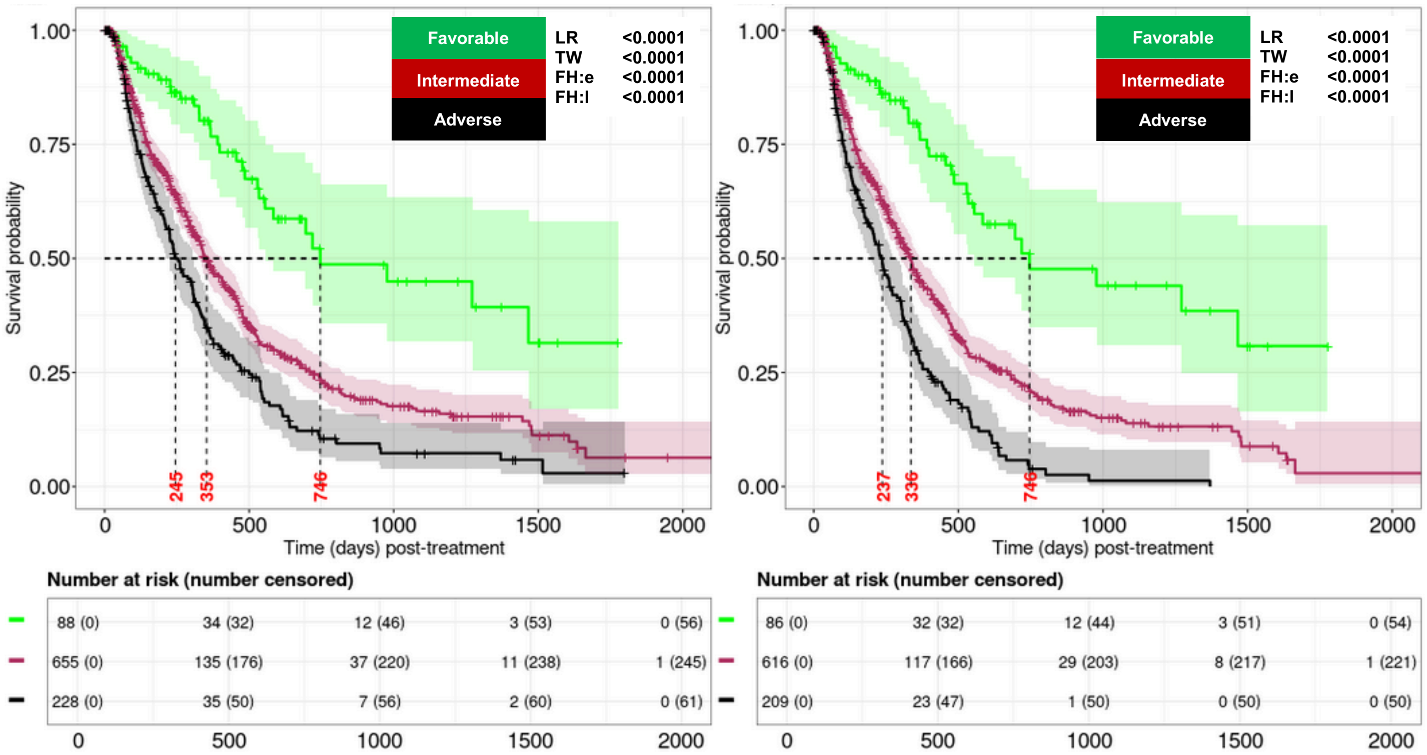

**Supplemental Figure 8, cont'd**

**B. ELN22 in the RWC CCAS cohort with allo-HCT included (left) and excluded (right) recipients.**

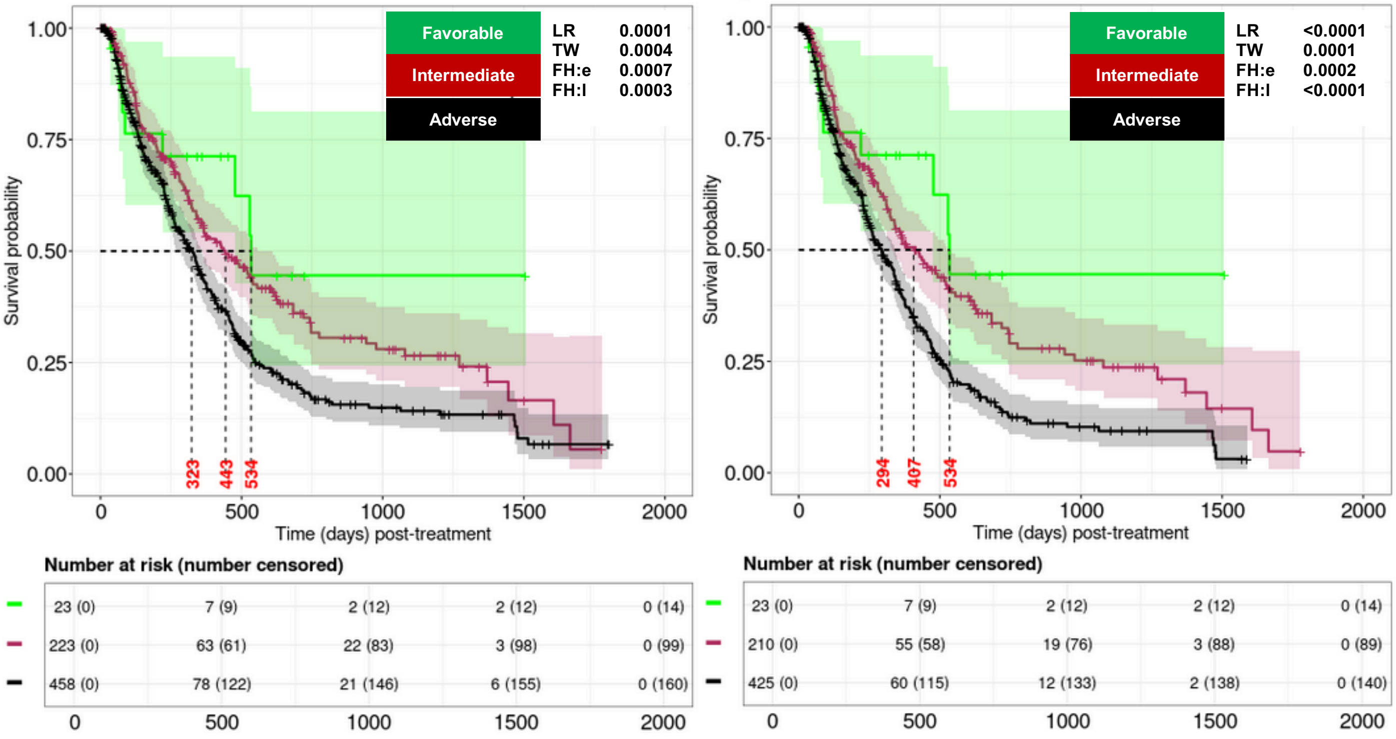

**Supplemental Figure 8, cont'd**

#### C. Pairwise comparisons between ELN22 and RRM (FAS for RRM and CCAS for ELN)

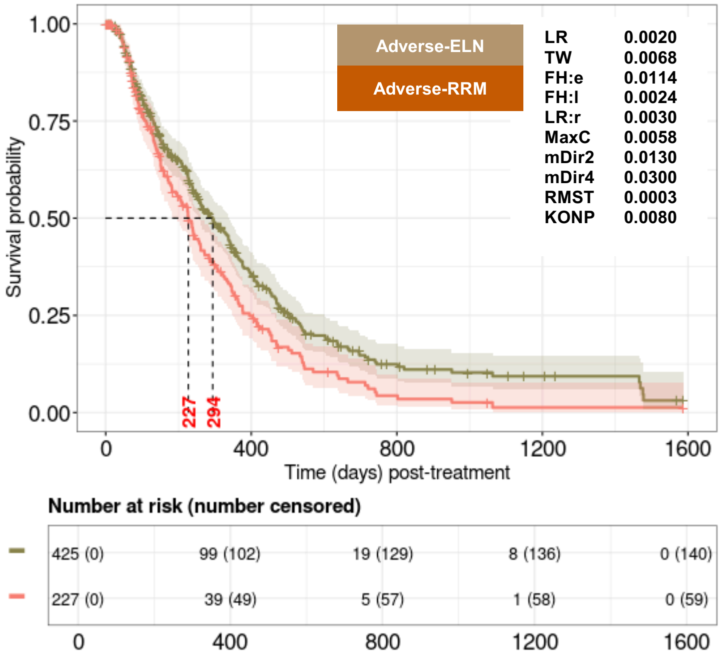


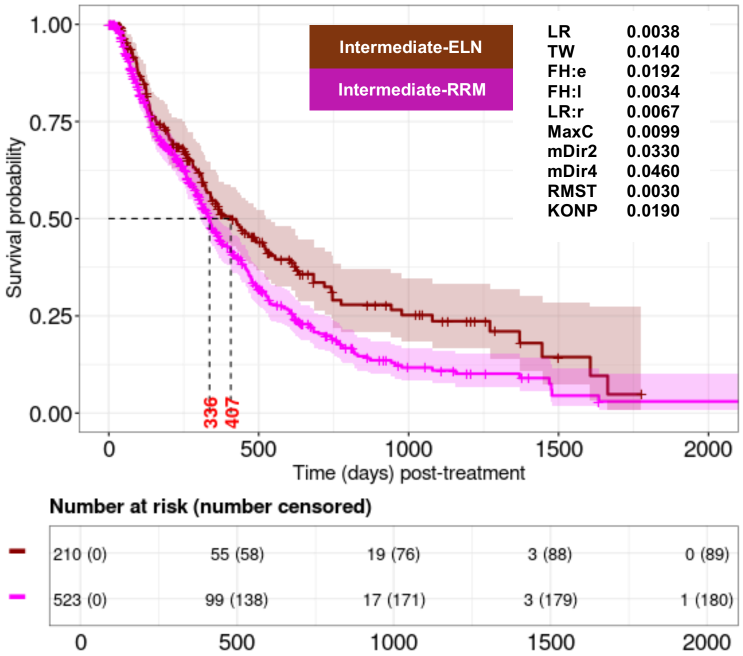


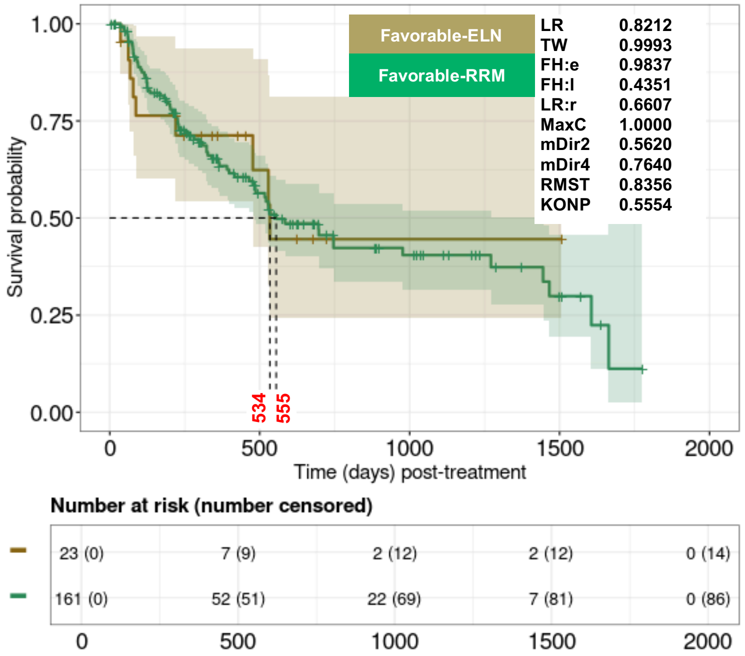

**Supplemental Figure 8, cont'd**

**D. Agreements between RRM and ELN22 risk models using the RWC dataset (FAS for RRM and CCAS for ELN).**

| Fleiss kappa (*P* values) | Favorable | Intermediate | Adverse |
|---|---|---|---|
| ***Within group*** | 0.13 (0.001) | 0.19 (<0.001) | 0.37 (<0.001) |
| ***Overall*** | | 0.25 (<0.001) | |

**Remarks:**

**The higher positive value (i.e., close to 1) means more agreement**
**The lower negative value (i.e., close to -1) means less agreement**
**Value close to 0 means agreement is no better than obtained by chance**

**Supplemental Figure 9. RRM, mPRS, and e-mPRS comparisons in the RWC dataset.**

A. **mPRS: RWC FAS (left panel) and RWC IAS (right panel), allo-HCT recipients excluded in both.**

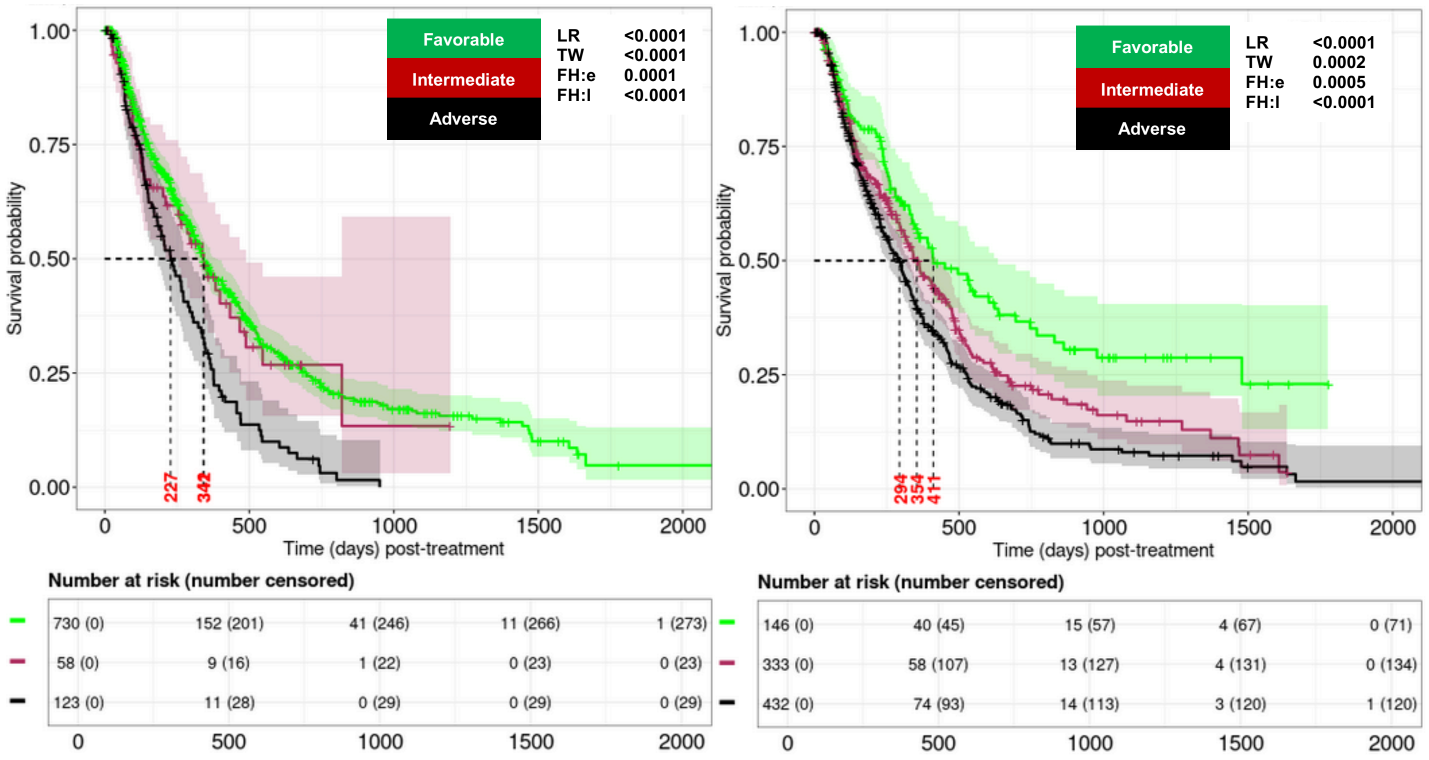

**Supplemental Figure 9, cont'd**

**B. e-mPRS: RWC CCAS (top-left panel), FAS (top-right panel), and IAS (bottom panel) with allo-HCT recipients excluded.**

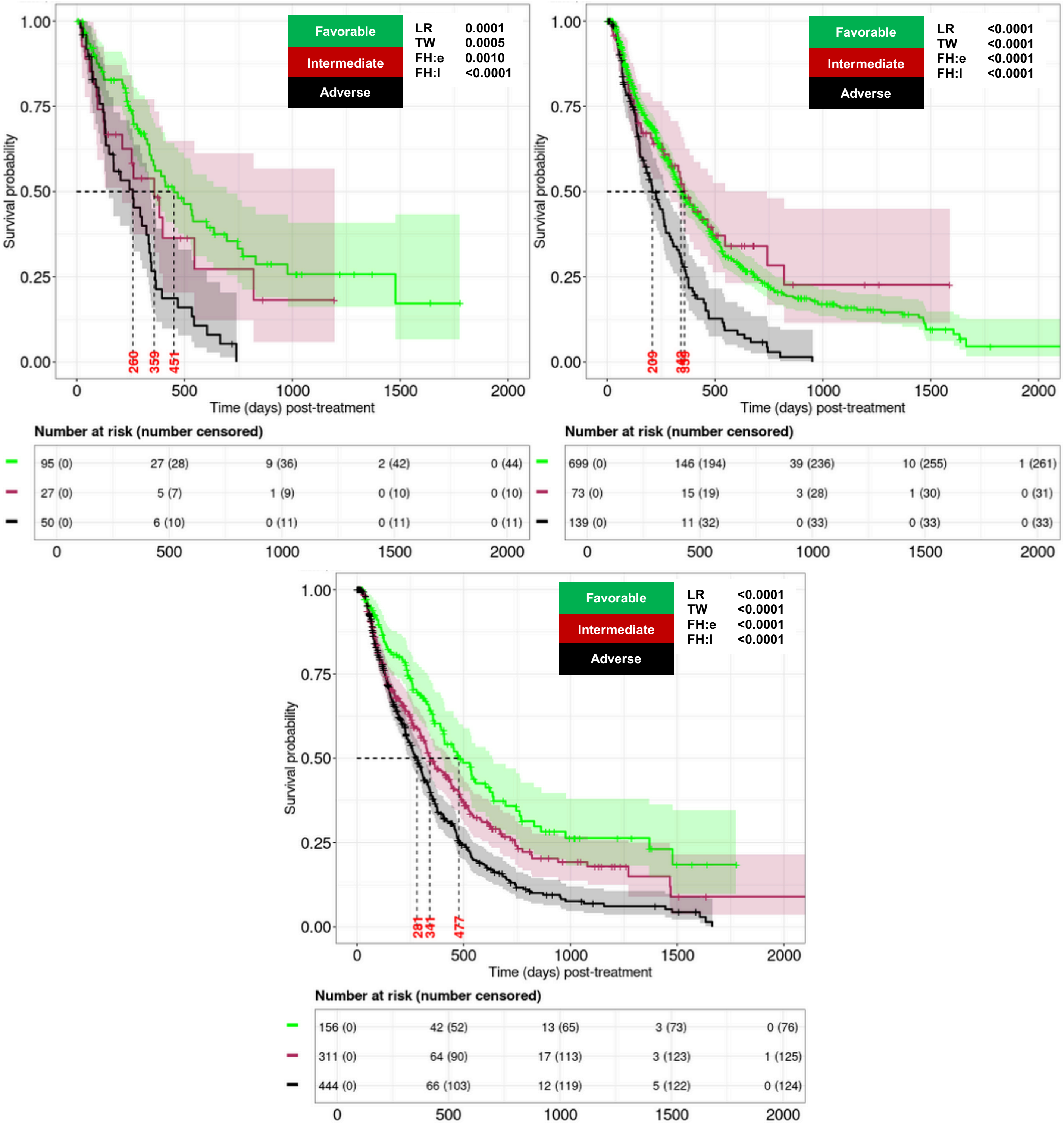

**Supplemental Figure 9, cont'd**

C. **Pairwise comparisons between the RRM and e-mPRS in the RWC dCCAS with allo-HCT patients excluded.**

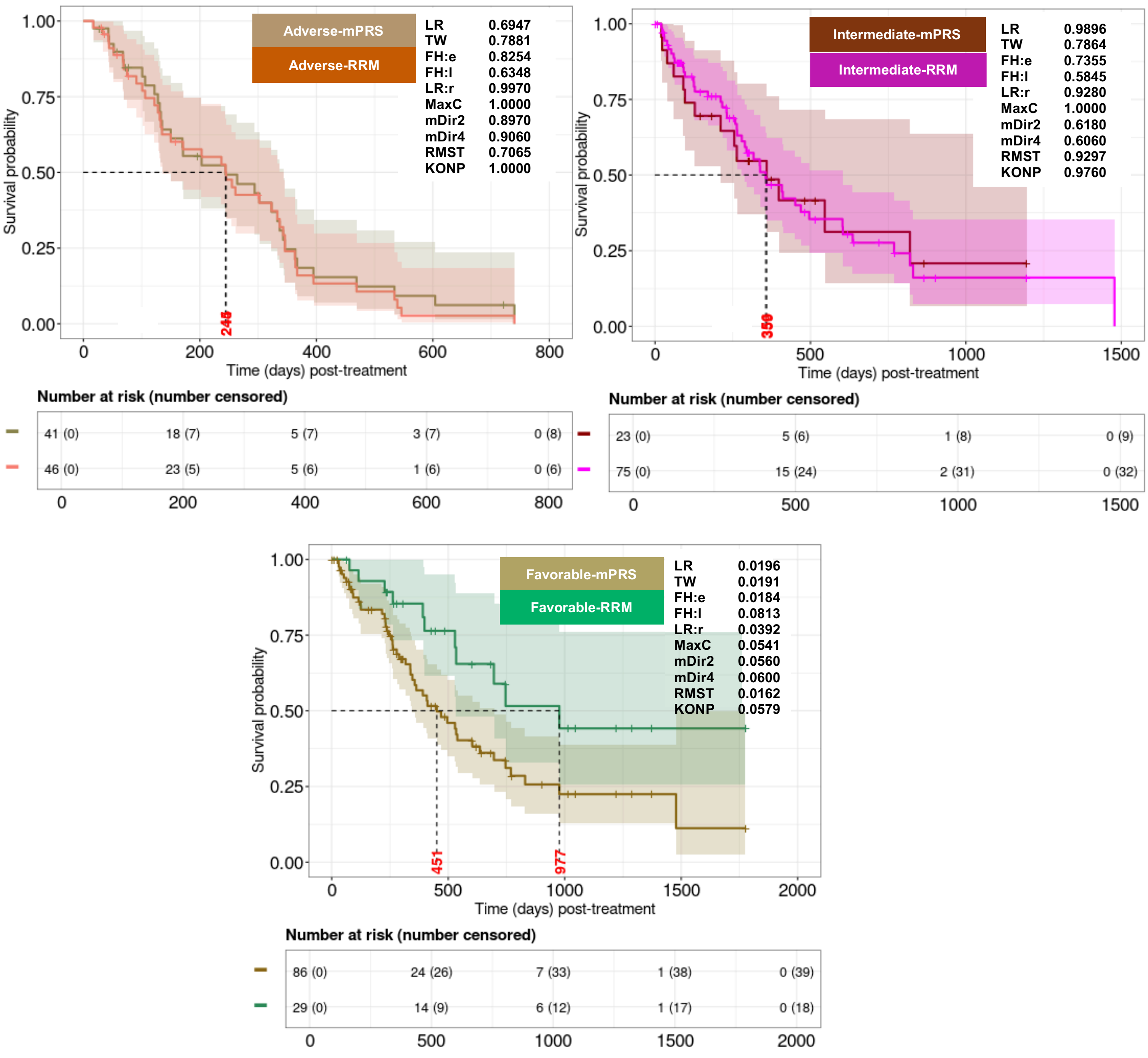

Supplemental Figure 9, cont'd

#### D. Agreements between the RRM and e-mPRS using the RWC dCCAS.

| Fleiss kappa (*P* values) | Favorable | Intermediate | Adverse |
|---|---|---|---|
| ***Within group*** | 0.15 (0.066) | 0.12 (0.130) | 0.54 (<0.001) |
| ***Overall*** | | 0.26 (<0.001) | |

**Remarks:**

**The higher positive value (i.e., close to 1) means more agreement**
**The lower negative value (i.e., close to -1) means less agreement**
**Value close to 0 means agreement is no better than obtained by chance**

**Supplemental Figure 10. Evaluation of predictive performance of RRM, mPRS, e-mPRS, and ELN22 risk models in the RWC for OS.**

#### A) FAS with mPRS

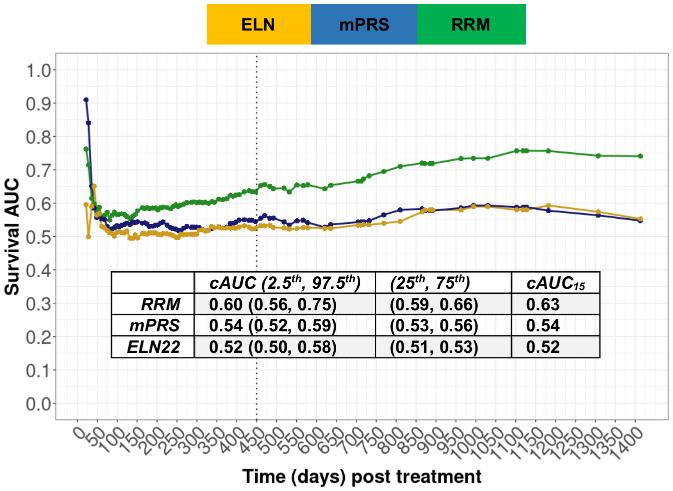


| | cAUC (2.5th, 97.5th) | (25th, 75th) | $cAUC_{15}$ |
|---|---|---|---|
| *RRM* | 0.60 (0.56, 0.75) | (0.59, 0.66) | 0.63 |
| *mPRS* | 0.54 (0.52, 0.59) | (0.53, 0.56) | 0.54 |
| *ELN22* | 0.52 (0.50, 0.58) | (0.51, 0.53) | 0.52 |

#### B) IAS with mPRS

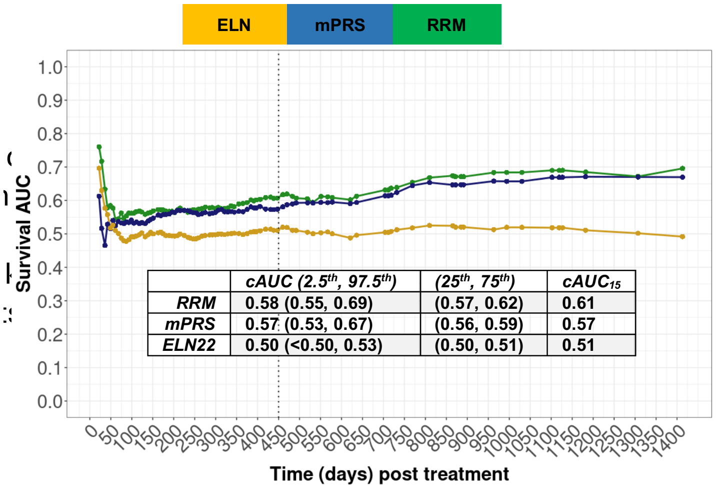


| | cAUC (2.5th, 97.5th) | (25th, 75th) | $cAUC_{15}$ |
|---|---|---|---|
| *RRM* | 0.58 (0.55, 0.69) | (0.57, 0.62) | 0.61 |
| *mPRS* | 0.57 (0.53, 0.67) | (0.56, 0.59) | 0.57 |
| *ELN22* | 0.50 (<0.50, 0.53) | (0.50, 0.51) | 0.51 |

#### C) FAS with e-mPRS

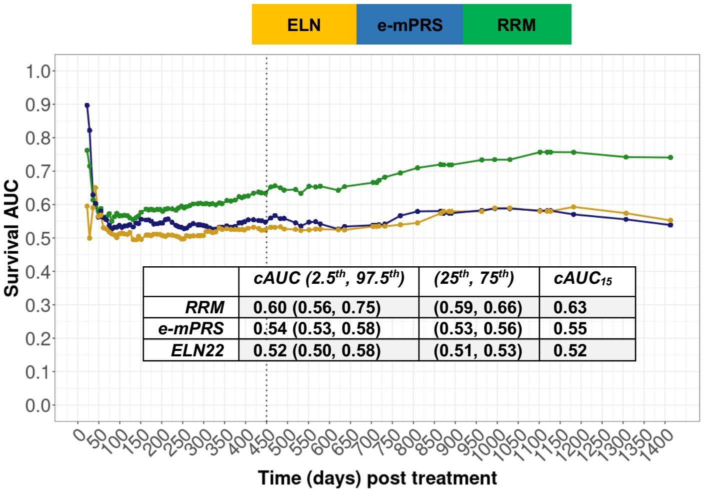


| | cAUC (2.5th, 97.5th) | (25th, 75th) | $cAUC_{15}$ |
|---|---|---|---|
| *RRM* | 0.60 (0.56, 0.75) | (0.59, 0.66) | 0.63 |
| *e-mPRS* | 0.54 (0.53, 0.58) | (0.53, 0.56) | 0.55 |
| *ELN22* | 0.52 (0.50, 0.58) | (0.51, 0.53) | 0.52 |

#### D) IAS with e-mPRS

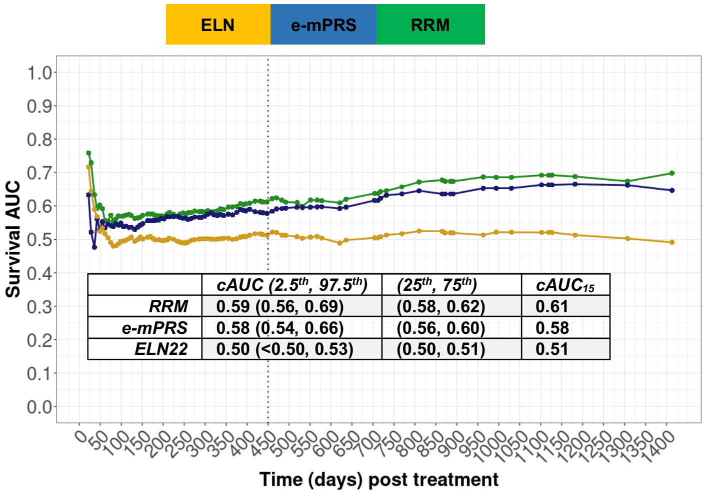


| | cAUC (2.5th, 97.5th) | (25th, 75th) | $cAUC_{15}$ |
|---|---|---|---|
| *RRM* | 0.59 (0.56, 0.69) | (0.58, 0.62) | 0.61 |
| *e-mPRS* | 0.58 (0.54, 0.66) | (0.56, 0.60) | 0.58 |
| *ELN22* | 0.50 (<0.50, 0.53) | (0.50, 0.51) | 0.51 |
